# Plasma proteomic analysis of intermuscular fat links muscle integrity with processing speed in older adults

**DOI:** 10.1101/2025.01.24.25320976

**Authors:** Toshiko Tanaka, Caterina Rosano, Xiaoning Huang, Qu Tian, Bennett A. Landman, Ann Z Moore, Iva Miljkovic, Andrew Perry, Sadiya Khan, Ravi Kalhan, John Jeffrey Carr, James G. Terry, Kristine Yaffe, Keenan Walker, Julián Candia, Luigi Ferrucci

## Abstract

**INTRODUCTION:** More intermuscular fat (IMF) has been associated with lower cognitive performance and faster age-associated decline in cognitive function however, the mechanisms driving this relationship have not been fully elucidated. We utilized proteomic analyses to identify the molecular mediators of the association between IMF and cognition to gain further insight into the mechanisms underlying this association.

**METHODS:** In this cross-sectional study, the plasma proteomic profile of IMF was assessed in the Baltimore Longitudinal Study on Aging (BLSA; n=941, age=66.7±15.2) and validated in the Coronary Artery Risk Development in Young Adults Study (CARDIA; n=2451, age=50.2±3.6). The 7628 plasma proteins were assessed using an aptamer-based assay and tested for association with IMF from the thigh (BLSA) and abdomen (CARDIA). Processing speed assessed by Digit Symbol Substitution Test (DSST). Associations between the main exposures, outcome and mediators were evaluated using linear regression, and mediating effects were assessed by causal mediation analysis adjusting for age, sex, muscle area or muscle volume, self-reported race, and years of education.

**RESULTS:** Higher IMF was associated with lower DSST performance both in the BLSA and CARDIA studies. There were 722 plasma proteins associated with IMF in both the discovery and replication cohorts (FDR-adjusted p≤0.05). Of the 722 IMF-associated proteins, 26 (24 unique proteins) mediated the relationship between IMF and processing speed with mediation effects ranging from 2.8 to 20.9% (p≤0.05). Overrepresentation analysis of the IMF-associated proteins showed enrichment of proteins in synaptic function and organization, and growth factor binding (FDR-adjusted p≤0.05).

**DISCUSSION:** There is a robust proteomic signature explaining, at least in part, the link of IMF with DSST. This signature reflected neurological function and growth factor regulation, which are both implicated in lower processing speed. Reducing IMF through behavioral or pharmacological intervention may improve cognition through reduction in growth factor activity and improvements in synaptic activity.

## Background

Aging is associated with decline in cognitive function across different domains [1].

Slowing rates of decline in cognitive function is an important public health goal, as cognitive decline is associated with unfavorable aging outcomes such as disability and dementia that could influence one’s quality of life [2]. There is growing evidence linking skeletal muscle health with cognitive decline, thus understanding the biology linking muscle with cognitive function may identify novel targets to slow the rates of cognitive decline with aging [3, 4].

Intermuscular fat (IMF) increases with older age, and it has been associated with worse cognition in older adults, particularly with poorer processing speed [3–9]. It has been hypothesized that neurodegenerative effects of IMF may be triggered by inflammatory and cardio-metabolic disorders. When adipocytes reach their fat storage capacity, greater adipose tissue accumulates ectopically, that is in organs that usually have little adiposity, such as the skeletal muscle. As adipose tissue accumulates in skeletal muscle, both adipocytes and myocytes may release pro-inflammatory cytokines and these in turn can increase risk of systemic metabolic abnormalities such as insulin resistance and type 2 diabetes [10, 11]. It has also been shown that myokines and adipokines (cytokines releases from myocytes and adipocytes, respectively) have a direct signaling effects on the central nervous system, influencing amyloid clearance and mitochondrial function in the brain [12–17]. However, evidence is primarily from animal models or human studies at the extremes of muscle health (e.g. athletes or patient populations); the molecules linking IMF with poorer cognitive function have not been directly measured in community-dwelling older adults without overt disease.

To address this gap in knowledge, we aimed to characterize the plasma proteomic profile of IMF and explore whether IMF-associated proteins mediate the relationship between IMF and cognitive function, focusing on processing speed. By taking an agnostic approach to explore the molecular pathways that are represented by the IMF-associated proteins, the results of this study provide mechanistic insights into the link between IMF and cognitive function.

## Methods

### Study Design and Cohort Description

The discovery cohort, the Baltimore Longitudinal Study of Aging (BLSA), is a rolling enrollment population-based study of participants residing predominantly in the Baltimore-Washington DC area [18]. Briefly, eligibility criteria include the absence of major chronic diseases (except for controlled hypertension), and cognitive or functional disabilities.

Participants are continually followed at varying time intervals based on their age, namely every 4 years for participants younger than 60 years, every 2 years between 60-79 years, and every year for participants older than 80 years. Demographic characteristics including age, sex, self-reported race, and years of education were obtained during a structured interview by BLSA staff. The study protocol (Protocol number 03-AG-0325) was approved by the National Institutes of Health Intramural Research Program Institutional Review Board and informed consent was obtained from participants at each visit.

The validation cohort, Coronary Artery Risk Development in Young Adults (CARDIA) study began in 1985 with the recruitment of 5,115 participants aged 18 to 30 years at field centers located in Birmingham, AL, Chicago, IL, Minneapolis, MN, and Oakland, CA [19].

Recruitment was balanced for equal inclusion of black and white as well as female and male participants, age (18–24, 25–30 years), and education (≤12 years, >12 years). The current study includes data from 3,172 participants who agreed and received abdominal CTs at the year 25 (Y25) examination, of these, a total of 2,390 had DSST, proteomics and other key factors such as age, sex, race, and education. All participants provided written informed consent, and institutional review boards from each field center and the coordinating center approved the study annually.

### Assessment of intermuscular fat

IMF was quantified using CT scan of the thigh (BLSA) or abdomen (CARDIA). In the BLSA, Imaging was performed by the Radiology Department of MedStar Harbor Hospital (Baltimore, MD) using a Somatom Sensation 10CT scanner (Siemens, Malvern, PA). A two-stage deep learning pipeline was implemented to estimate IMF from 2D thigh CT slices [20, 21]. From this algorithm, the total muscle area, and IMF was calculated as the average of the right and left thigh. In the CARDIA study, scans were performed using 64-channel multidetector GE CT scanners (GE Healthcare Milwaukee, WI) at the Birmingham, AL, and Oakland, CA, centers and Siemens CT scanners (Siemens, Erlangen, Germany) at the Chicago, IL, and Minneapolis, MN, centers. The protocol has been previously published [22, 23].

### Cognitive function assessment

In both cohorts, the Digit Symbol Substitution Test (DSST) was used to evaluate processing speed [24]. During the test, participants are initially shown nine pairs of digits and symbols. In the next sequence participants are provided a series of digits and asked to draw the matching symbols for each digit. The score is the number of correct digit-symbol pairs identified during a period of 90 seconds. The DSST values were analyzed as a z-score.

### Proteomic Assessment

Proteomic assessment in fasting plasma samples were conducted using the 7k SomaScan assay v4.1 (SomaLogic, Inc.; Boulder, CO, USA) in both BLSA and CARDIA, following a protocol that was detailed elsewhere [25, 26]. The final normalized dataset (hybridization control, median signal, plate-scale, inter-plate normalization) was used for the analysis. The final protein count consisted of 7,268 target annotated human SOMAmers that were assessed and passed quality control in both the BLSA and CARDIA study. Some proteins were targeted by multiple SOMAmers, thus 6583 unique human proteins are represented in the panel. From this point forward, each SOMAmer will be referred to proteins in the manuscript. Each protein relative abundance (RFU) was log2 transformed and scaled before analysis.

### Statistical Analysis

All analyses were conducted using R version 4.2.2. The associations between IMF and plasma proteins or DSST were assessed using linear regression models. For proteomic analysis of IMF, the winsorized protein value was the dependent variable, and IMF the independent variable in the regression model. The models were adjusted for age, sex, muscle area (or muscle volume for CARDIA), self-reported race, and years of education. For the proteomic analysis, the p-value was adjusted using false discovery rate (FDR), and an FDR adjusted p≤0.05 was considered statistically significant. Proteins that were significantly associated with IMF in the BLSA were tested for validation in CARDIA study. Significant validation was considered at FDR adjusted p≤0.05. To test whether the IMF-associated proteins mediate the association between IMF and DSST, a causal mediation analysis was conducted using R regmedint package (version 1.0.1)[27]. For each model, IMF was the main exposure variable, DSST the dependent variable, and individual protein was the mediator. Mediation was considered significant when a significant indirect association for IMF on DSST was observed (p≤0.05).

Over-representation analysis (ORA) was conducted to determine whether any pathways were enriched among the IMF-associated proteins. Using the proteins measured by the SomaScan platform as background, we used the 716 IMF-associated proteins to explore enrichment of GO biological processes (BP), cellular component (CC), and molecular function (MF) libraries. Enrichment was considered significant at FDR adjusted p≤0.05. The over-representation analysis was performed using R ClusterProfiler package (version 4.6.2).

## Results

*Association between intermuscular fat and DSST.* The average age of the discovery cohort was older than the validation cohort (**Table 1**; 66.7±15.2 vs 50.2±3.6). The age difference is reflected in processing speed, with higher DSST scores in the CARDIA study compared to BLSA. Despite these differences, IMF was found to be statistically significant in both studies. In the BLSA, there was significant negative relationship between IMF and DSST (ß[95%CI]=-0.11 [-0.15,- 0.05]) indicating that higher IMF was associated with slower DSST. Similarly, in the CARDIA study a negative relationship was also observed (ß[95%CI]=-0.09[-0.12,-0.05]).

*Proteomics of intermuscular fat –* In the discovery cohort, of 7268 SOMAmers (6375 proteins) assessed, 782 SOMAmers (710 proteins) were significantly associated with IMF (Supplemental Table 1; Figure 1A). There were 321 SOMAmers (297 proteins) found in higher abundance with greater IMF, and 461 SOMAmers (413 proteins) in lower abundance with higher IMF. These 782 SOMAmers were tested for association with IMF in the CARDIA study, and 722 (654 proteins; 92.3%) were found to be significantly associated. For all proteins, the direction of association was consistent, with high correlation (r=0.95, Figure 1B) between the beta estimates from the regression models from the two studies for the 782 SOMAmers that were tested for replication.

**Figure 1.**
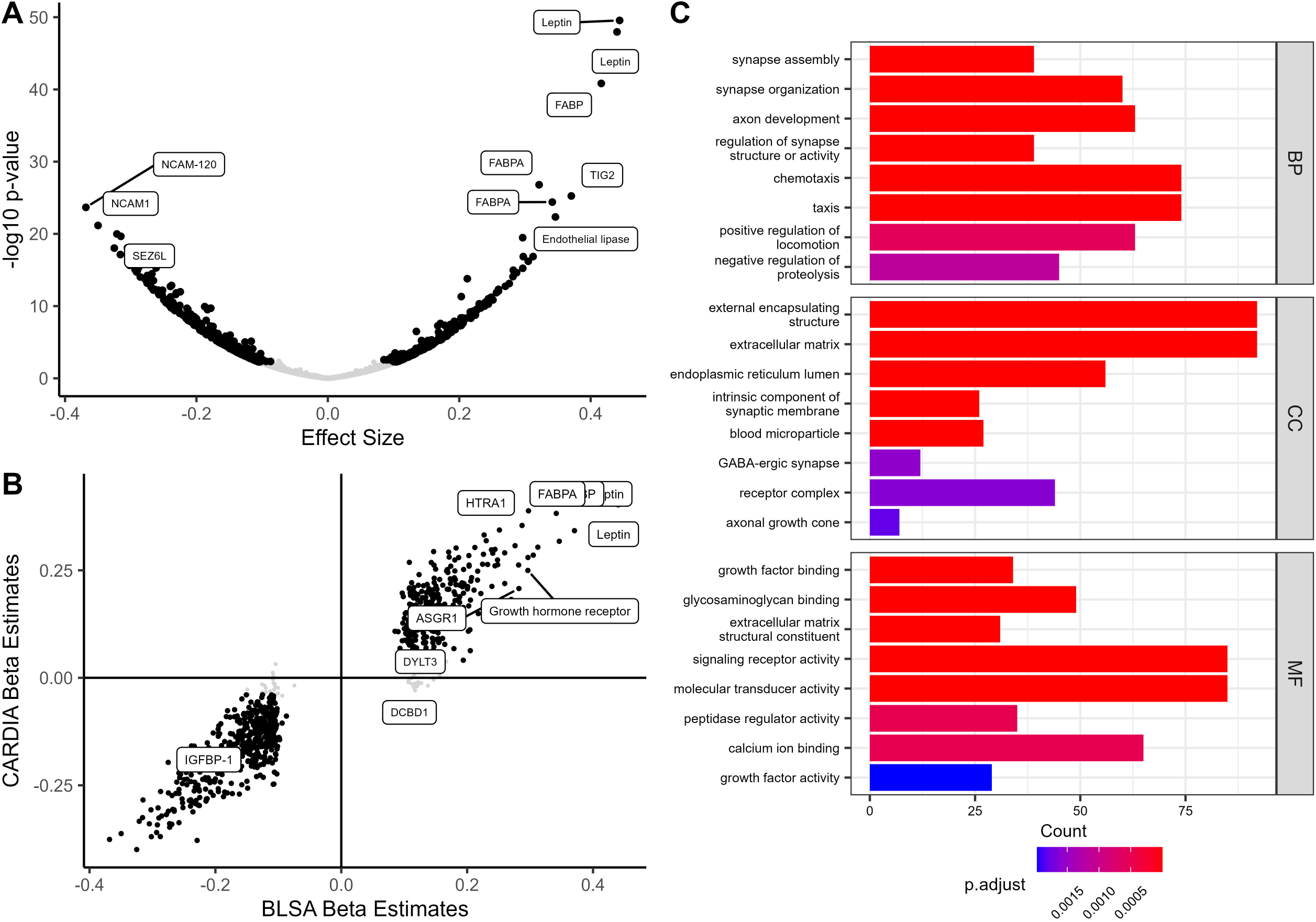
Plasma proteomic profile of intermuscular fat. Associations between intermuscular fat (IMF) and plasma proteins were tested in participants of the BLSA (n=941) and validated in the CARDIA study (n=2445). (A) In the discovery BLSA study, of the 7268 SOMAmers assessed, 782 were associated with IMF. (B) Of the 782 SOMAmers, 722 were validated in the CARDIA study. The associations between the beta estimates from the regression between IMF and SOMAmers in the BLSA and CARDIA study was high (r=0.96). (C) Enrichment analysis of GO biological process (BP), cellular component (CC), and molecular functions (MF) of the 722 SOMAmers (representing 654 unique proteins) show enrichment in neurological pathways and growth factor binding.

An ORA analysis based on GO terms for the 654 proteins represented by the 722 IMF-associated SOMAmers, identified multiple pathways (Figure 1C). The GO-biological process (BP) terms encompass two broad categories of pathways that represent synaptic biology ( “regulation of synapse assembly”, “regulation of synapse structure and activity”, “axon development”) and inflammation (“taxis” and “chemotaxis”). The GO-cellular component (CC) terms complement the results from synaptic function from the BP that include CC terms such as “intrinsic component of synaptic membrane, GABA-ergic synapse” and “axonal growth cone”, as well as indicator of extracellular matrix. Finally, the GO-molecular function (MF) identified included “glycosaminoglycan binding” and “growth factor binding”, and terms representing general signal transduction activities.

*Proteomic mediators of IMF and DSST.* Of the 722 IMF-associated SOMAmers, 26 SOMAmers representing 24 proteins significantly mediated the relationship between IMF and DSST in both BLSA and CARDIA studies (Table S2; Figure 2). The proportion mediation ranges were 7.2- 20.9% in BLSA and 2.8-8.9% in CARDIA. For all but one protein, the directions of indirect effects was negative in both the BLSA and CARDIA studies. Among the 24 mediating proteins, 14 were found in lower abundance with higher IMF, and associated with higher DSST. Conversely, 10 proteins were found in higher abundance with greater IMF and associated with lower DSST. The ORA analysis of the 24 proteins that mediate the relationship represent pathways showed no significantly enriched pathways.

**Figure 2.**
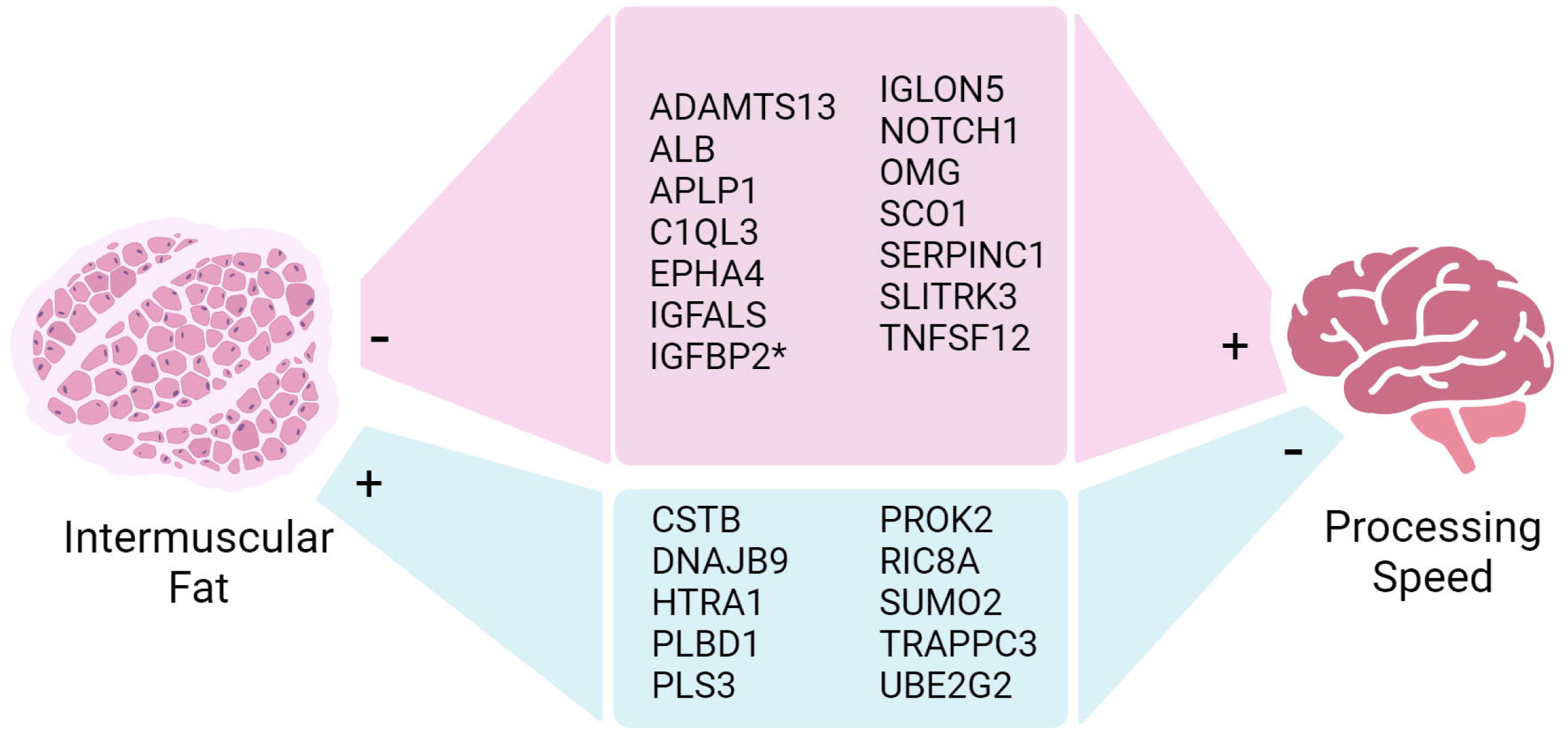
Association between intermuscular fat and processing speed mediation by plasma proteins. The association between degree of intermuscular fat (IMF) and processing speed assessed using DSST was significantly mediated by 24 proteins in both the BLSA and CARDIA studies. Except for IGFBP2, all proteins show negative indirect effects. Red: There were 14 proteins that had negative associations with IMF (lower levels for higher IMF) and positive association with DSST, indicating a potential neuroprotective role. * IGFBP2 was the only protein with a negative association with DSST in CARDIA. Blue: there were 10 proteins with positive associations with IMF (higher levels for higher IMF) and negative associations with DSST, indicating a potential neurodegenerative role. Created with BioRender.com

## Discussion

In this study, we explored the plasma proteomic profiles of IMF showed that there is a distinct proteomic signature of 654 proteins in two independent cohorts of older adults living in the community. The protein signature was highly consistent despite IMF being measured in skeletal muscle at different locations. A subset of 24 IMF-associated proteins explain, at least in part, the association between higher IMF and slower processing speed. Taking an agnostic approach, enrichment analyses of the proteins identified indicate that pathways such as synaptic function and growth factor activity may be the link between IMF and processing speed.

The study results confirm the growing evidence supporting the associations between higher IMF and lower cognitive function, particularly with slower processing speed. Since the initial reporting inverse associations of measures of IMF using peripheral quantitative computer tomography (pQCT) with measured of psychomotor function, visual learning, and overall cognitive function [28], similar associations have been reported in different populations particularly with processing speed [3, 4, 9] and with other indicators of brain health such as neuroimaging markers [29, 30]. Our study explored used proteomic to explore the mechanisms that may link higher IMF with slower processing speed.

One intriguing insight from our analysis was that the proteome associated with IMF was enriched with proteins involved in synaptic organization and function. The four proteins (SLITRK3, NOTCH1, C1QL3, EPHA4) that significantly mediated the association were found in lower abundance with higher IMF and lower processing speed. These proteins are expressed widely across the central nervous system, with some also found in muscle tissue [31–34]. They play crucial roles in brain function by influencing various processes: SLITRK3 regulates neurite outgrowth [32]; C1QL3 participate in neuronal organization as trans-synaptic cell adhesion complexes [33]; NOTCH is involved in neuronal stem cell maintenance and activation [31, 34]; and EPH4 is crucial for synaptic function and integrity through regulation of bidirectional communication between neurons and astrocytes [35]. SLITRK3 knockout mice have impaired postsynaptic neurotransmission resulting in multiple abnormal hypermobile movements[36].

C1QL3 knockout animals display fewer excitatory neurons compared to wildtype and exhibited stunted fear conditioning as well as a modest deficit in motor learning [37]. In humans, several lines of evidence support the link between these proteins with cognitive function. First, lower abundance of soluble NOTCH1 protein was reported in patients with AD, further protein abundances were positively correlated with MMSE [38]. Mendelian randomization study supported the causal association between SLITRK3 with general cognitive function [39]. Taken together, our data suggest lower levels of proteins involved in synaptic function and integrity simultaneously manifest as lower integrity in the skeletal muscle (higher IMF) and central nervous system (slower processing speed). Our results of causal modeling analyses indicate a potential direction of these associations, whereby higher IMF may affect processing speed by lowering the circulating levels of neuroprotective proteins.

Among the proteins that were found in higher abundance in relation to higher IMF and lower DSST (e.g. with potential neurodegenerative effects), the myokine High-Temperature Requirement A1 (HTRA1) is notable, for its proteolytic effects on extracellular matrix remodeling skeletal muscle, brain, and adipose tissue[40, 41]. HTRA1 has also inhibitory effects on the transforming growth factor-β superfamily. In a translational proteomic study of several thousand proteins[42]that HTRA1 was positively correlated with brain amyloid. Several HTRA1 mutations have been associated with cerebral small vessel disease in humans [43], but these studies did not test associations with cognitive function. We and others have shown that higher cerebral small vessel disease is strongly associated with lower processing speed; thus it is possible that higher IMF leads to higher levels of the myokine HTRA1, this in turn drives cerebral small vessel disease, manifesting as slower processing speed.

The finding that proteins related to IGF are related with IMF and cognition has been previously shown. Of note, IGF binding protein 2 (IGFBP-2) was positively associated with skeletal muscle mass and strength in prior proteomic studies in South Africa[44], and Germany[45]. Conversely, its role in relation with cognition is less clear. In a recent study of older adults[46], IGFBP2 was associated with smaller hippocampal volume among amyloid negative adults, but not amyloid positive, indicating the influence on IGFBP2 on hippocampal volume varies depending on severity of brain pathology; conversely, others found a positive association with tau[48]. The association of IGFBP2 with cognitive status also varies across studies, presumably because the role of IGFBP-2 varies depending on developmental stages[47, 48]. Differences in age, and/or in brain pathology may explain why in our study IGFPB-2 had an inverse association with DSST in CARDIA (younger) but not in BLSA (older).

Prior studies have linked IMF with factors that influence cardiometabolic health, in particular serum cytokine levels and adipokines [11]. Both muscle and the fat that infiltrates muscle have secretory capabilities including pro-and anti-inflammatory cytokines that have been associated with cognitive health [49]. Moreover, cytokines released by skeletal muscles and adipocytes with neuronal signaling properties can cross the blood brain barrier [50].

Consequently, some studies explored these proteins as possible mechanism linking IMF with cognitive function. In the Geelong osteoporosis study (GOS), TNFα explained the association between muscle density measured in the radius and psychomotor function, but the mediation was not established for the muscle density measured in the tibia [28]. Further, other inflammatory markers, IL-6, IL-10, and IL-13 did not explain the association in the GOS study. In other studies, while no formal mediation analysis was conducted, the association between IMF and cognitive function were robust to adjustment for cardiometabolic conditions, including hypertension, diabetes, obesity, measures of inflammation (leptin, adiponectin, IL-6) [4]. In the current report, IL-6 was moderately associated with IMF in the BLSA, and CRP was negatively associated with IMF in both BLSA and CARDIA studies but were not significant mediators.

Intriguingly, enrichment analysis did not show inflammation as a major mediating pathway linking IMF with processing speed.

While our findings provide strong biological evidence linking IMF with processing speed, there are several important limitations. First, this is a cross-sectional study, thus we cannot make causal inferences on the associations we observe. For example, it is possible that higher IMF may drive lower levels of proteins, which in turn compromise synaptic integrity in the brain, ultimately manifesting as slower processing speed. Alternatively, there is another factor that alters homeostasis of proteins engaged in synapsis remodeling among older adults with higher IMF. Further, there may be a bi-directional relationship – those with lower cognitive function and processing speed may have lowered synaptic activity leading to decreased muscle quality. While our study uses direct measurements of IMF using CT scans, the levels of IMF are correlated with other measures of adiposity thus we cannot exclude the possibility that the correlations we observed are not driven by fat infiltrations in the muscle per se, but through overall adiposity. The analysis was not adjusted for measures of adiposity due to the correlation between the two traits, and overall adiposity may be part of the casual pathways linking intermuscular fat with processing speed. Our study has several strengths. First, our results were consistent across two studies that have different demographics from the standpoint of age, self-reported race, and geographical distributions. In both studies, IMF was measured using CT-scan providing us with an objective measure. Finally, while the proteomic assessment did not measure all circulating proteins, the analysis targeted large number of proteins using the same assessment method. Future studies should explore validating these findings in longitudinal studies to determine the temporal relationship between proteins abundances, IMF, and processing speed.

By leveraging data on circulating proteins, we provide strong evidence suggesting that IMF affects processing speed through inflammation, regulating of growth factor activity, and synaptic organization, function and activity. These data support the interplay between the muscular quality and the brain and suggest that the muscle is an important target for prevention of cognitive decline associated with aging.

## Data Availability

Data for this study can be accessed at the BLSA data coordinating center (https://www.blsa.nih.gov), and CARDIA coordinating center (www.cardia.dopm.uab.edu).

https://www.cardia.dopm.uab.edu

## Acknowledgement

The BLSA was supported by the Intramural Research Program of the NIH, National Institute on Aging

## Conflict of interest

The authors have no conflicts to declare.

## Funding Sources

The Coronary Artery Risk Development in Young Adults Study (CARDIA) is conducted and supported by the National Heart, Lung, and Blood Institute (NHLBI) in collaboration with the University of Alabama at Birmingham (75N92023D00002 & 75N92023D00005), Northwestern University (75N92023D00004), University of Minnesota (75N92023D00006), and Kaiser Foundation Research Institute (75N92023D00003). This manuscript has been reviewed by CARDIA for scientific content. CARDIA proteomics data was funded through the CARDIA Lung Study (R01 HL122477).

## Consent Statement

All participants of the study in this report provided informed consent

## Notes

### Competing Interest Statement

The authors have declared no competing interest.

