## Supplemental Tables for "Plasma proteomic analysis of intermuscular fat links muscle integrity with processing speed in older adults"

**Supplemental Table 1**. Association between plasma proteins and thigh (BLSA) or abdominal (CARDIA) intermuscular fat**.**

| SeqId | SomaId | Target | Gene | uniprot | BLSA (Discovery) | | | | CARDIA (Validation) | | | |
| --- | --- | --- | --- | --- | --- | --- | --- | --- | --- | --- | --- | --- |
|  |  |  |  |  | beta | SE | p | FDR-p | beta | SE | p | FDR-p |
| 2575-5 | SL000498 | Leptin | LEP | P41159 | 0.444048 | 0.028016 | 2.8007566362793e-50 | 2.03558992324779e-46 | 0.40552 | 0.014795 | 2.1e-144 | 1.6422e-141 |
| 8484-24 | SL000498 | Leptin | LEP | P41159 | 0.4401 | 0.028292 | 1.09203442637521e-48 | 3.96845310544751e-45 | 0.402335 | 0.015057 | 3.5e-138 | 1.3685e-135 |
| 5437-63 | SL001774 | FABP | FABP3 | P05413 | 0.416143 | 0.029315 | 1.4840133167113e-41 | 3.59526959528591e-38 | 0.433022 | 0.016939 | 6.6e-128 | 1.7204e-125 |
| 15386-7 | SL005086 | FABPA | FABP4 | P15090 | 0.321509 | 0.028636 | 1.58518123713396e-27 | 2.8802743078724e-24 | 0.416449 | 0.017525 | 1.9e-112 | 3.7145e-110 |
| 3079-62 | SL005152 | TIG2 | RARRES2 | Q99969 | 0.370509 | 0.034101 | 5.63948070484618e-26 | 8.19754915256441e-23 | 0.342217 | 0.019613 | 2.45e-64 | 8.70863636363636e-63 |
| 9851-9 | SL005086 | FABPA | FABP4 | P15090 | 0.341582 | 0.032034 | 3.95647177956144e-25 | 4.79260614897542e-22 | 0.382411 | 0.017993 | 4.2e-92 | 6.5688e-90 |
| 4498-62 | SL003764 | NCAM-120 | NCAM1 | P13591 | -0.36828 | 0.035122 | 2.1434496938972e-24 | 2.22551319646355e-21 | -0.37589 | 0.019782 | 3.45e-75 | 2.24825e-73 |
| 23148-100 | SL005612 | Endothelial lipase | LIPG | Q9Y5X9 | 0.346377 | 0.034098 | 4.58458176825581e-23 | 4.16509253646041e-20 | 0.317553 | 0.019389 | 2.81e-57 | 6.65884848484848e-56 |
| 20161-41 | SL018262 | NCAM1 | NCAM1 | P13591 | -0.34978 | 0.035469 | 6.86828323926399e-22 | 5.54652028699674e-19 | -0.36234 | 0.019891 | 1.29e-69 | 6.304875e-68 |
| 19563-3 | SL008781 | SEZ6L | SEZ6L | Q9BYH1 | -0.32107 | 0.033592 | 1.02791728578828e-20 | 7.47090283310922e-18 | -0.33377 | 0.017122 | 8.96e-79 | 6.36974545454545e-77 |
| 2771-35 | SL000462 | IGFBP-1 | IGFBP1 | P08833 | -0.31505 | 0.033263 | 2.19173160053719e-20 | 1.44813684297312e-17 | -0.28406 | 0.019112 | 6.73e-48 | 1.01208846153846e-46 |
| 2948-58 | SL005168 | Growth hormone receptor | GHR | P10912 | 0.296504 | 0.03147 | 3.38327273133967e-20 | 2.04913551761473e-17 | 0.249721 | 0.019808 | 2.38e-35 | 1.7232962962963e-34 |
| 10521-10 | SL025939 | MXRA8:ECD | MXRA8 | Q9BRK3 | -0.30498 | 0.03372 | 8.54782390287919e-19 | 4.77889108662508e-16 | -0.33958 | 0.018483 | 1.04e-70 | 5.42186666666667e-69 |
| 5843-60 | SL003542 | NG36 | EHMT2 | Q96KQ7 | -0.32503 | 0.035983 | 9.40415490058565e-19 | 4.88209984410403e-16 | -0.39941 | 0.019708 | 1.6e-84 | 1.78742857142857e-82 |
| 7194-36 | SL008497 | NPTN | NPTN | Q9Y639 | -0.31578 | 0.035937 | 7.26599788232431e-18 | 3.52061817391554e-15 | -0.32521 | 0.020032 | 2.33e-56 | 5.359e-55 |
| 7638-30 | SL005403 | Lectin, mannose-binding 2 | LMAN2 | Q12907 | 0.312336 | 0.035867 | 1.38673334996934e-17 | 6.01225583078528e-15 | 0.303783 | 0.020311 | 1.85e-48 | 2.8934e-47 |
| 15594-47 | SL008268 | HTRA1 | HTRA1 | Q92743 | 0.29743 | 0.034162 | 1.40627888171918e-17 | 6.01225583078528e-15 | 0.388443 | 0.019603 | 3.78e-81 | 3.2844e-79 |
| 4983-6 | SL003522 | ERP29 | ERP29 | P30040 | 0.305044 | 0.03579 | 6.18158725755059e-17 | 2.49598756599321e-14 | 0.285279 | 0.020697 | 1.12e-41 | 1.0948e-40 |
| 8958-51 | SL008936 | CHL1 | CHL1 | O00533 | -0.30239 | 0.035919 | 1.42314659217779e-16 | 5.44391022734114e-14 | -0.30707 | 0.020896 | 6.6e-47 | 9.21642857142857e-46 |
| 15559-5 | SL011048 | ANTR2 | ANTXR2 | P58335 | -0.30181 | 0.036099 | 2.25540721506228e-16 | 8.19614981953632e-14 | -0.36953 | 0.020008 | 2.19e-71 | 1.22327142857143e-69 |
| 21548-20 | SL014786 | GGT5 | GGT5 | P36269 | -0.28711 | 0.034433 | 2.68648540522925e-16 | 9.29779805962199e-14 | -0.2645 | 0.019398 | 7.46e-41 | 6.78339534883721e-40 |
| 15573-110 | SL008782 | CSPG3 | NCAN | O14594 | -0.28753 | 0.034688 | 3.94948570066279e-16 | 1.30476645783714e-13 | -0.36929 | 0.018633 | 3.55e-81 | 3.2844e-79 |
| 13741-36 | SL000462 | IGFBP-1 | IGFBP1 | P08833 | -0.26138 | 0.03166 | 5.12452980434234e-16 | 1.61935141817218e-13 | -0.24258 | 0.019132 | 1.01e-35 | 7.52209523809524e-35 |
| 9199-6 | SL018946 | UB2G2 | UBE2G2 | P60604 | 0.296654 | 0.036004 | 5.81628222047544e-16 | 1.76136413243398e-13 | 0.279849 | 0.019455 | 4.29e-45 | 5.41093548387097e-44 |
| 13408-23 | SL010391 | WFKN2 | WFIKKN2 | Q8TEU8 | -0.2922 | 0.035499 | 6.19509072636663e-16 | 1.80103677596931e-13 | -0.34208 | 0.01993 | 1.98e-62 | 5.52985714285714e-61 |
| 2658-27 | SL004639 | TrkC | NTRK3 | Q16288 | -0.2934 | 0.035824 | 8.54756127195194e-16 | 2.38937212786718e-13 | -0.35984 | 0.020546 | 8.74e-65 | 3.25460952380952e-63 |
| 5452-71 | SL008835 | ASGR1 | ASGR1 | P07306 | 0.282256 | 0.034588 | 1.07275368755157e-15 | 2.88769400041661e-13 | 0.207652 | 0.01908 | 5.71e-27 | 2.68989156626506e-26 |
| 20187-10 | SL002726 | Integrin aVb3 | ITGAV\|ITGB3 | P06756\|P05106 | -0.28985 | 0.035566 | 1.16735142222251e-15 | 3.03011076311185e-13 | -0.32602 | 0.020119 | 3.61e-56 | 7.84172222222222e-55 |
| 7218-87 | SL018093 | AT1B2 | ATP1B2 | P14415 | -0.29112 | 0.035998 | 1.88773750323172e-15 | 4.73106074947868e-13 | -0.30247 | 0.020297 | 3.95e-48 | 6.05666666666667e-47 |
| 4337-49 | SL000051 | CRP | CRP | P02741 | 0.287233 | 0.035648 | 2.37230648156435e-15 | 5.74730783600324e-13 | 0.354325 | 0.019833 | 3.51e-67 | 1.5249e-65 |
| 6478-2 | SL017460 | IGLO5 | IGLON5 | A6NGN9 | -0.26773 | 0.033416 | 3.35208222894184e-15 | 7.85901085159655e-13 | -0.30605 | 0.01781 | 1.45e-62 | 4.19962962962963e-61 |
| 8890-9 | SL018697 | T132B | TMEM132B | Q14DG7 | -0.27476 | 0.034607 | 5.81092983764558e-15 | 1.31980743937525e-12 | -0.3103 | 0.018891 | 1.38e-57 | 3.372375e-56 |
| 10565-19 | SL017951 | SLIK3 | SLITRK3 | O94933 | -0.27466 | 0.034758 | 7.69384472214337e-15 | 1.69451101334964e-12 | -0.26772 | 0.02031 | 2.22e-38 | 1.84685106382979e-37 |
| 17706-4 | SL020934 | PPR1A | PPP1R1A | Q13522 | 0.28177 | 0.035698 | 8.23961078209569e-15 | 1.76133797541975e-12 | 0.26235 | 0.019628 | 2.25e-39 | 1.97696629213483e-38 |
| 18347-15 | SL021367 | Laminin-2 | LAMA2\|LAMB1\|LAMC1 | P24043\|P07942\|P11047 | -0.28546 | 0.036244 | 9.36910230044794e-15 | 1.9455610148473e-12 | -0.32141 | 0.020436 | 3.69e-53 | 7.038e-52 |
| 7211-2 | SL005355 | RNase 1 | RNASE1 | P07998 | 0.212236 | 0.027197 | 1.60942845491384e-14 | 3.24925722508717e-12 | 0.303086 | 0.01728 | 5.91e-65 | 2.31081e-63 |
| 3235-50 | SL010391 | WFKN2 | WFIKKN2 | Q8TEU8 | -0.27786 | 0.035757 | 2.0558749424735e-14 | 4.03840515726957e-12 | -0.31816 | 0.019725 | 1.09e-55 | 2.30372972972973e-54 |
| 3461-58 | SL009089 | PGCB | BCAN | Q96GW7 | -0.26546 | 0.034366 | 2.88903102066273e-14 | 5.52565196267808e-12 | -0.31032 | 0.019877 | 1.92e-52 | 3.41236363636364e-51 |
| 9296-15 | SL008499 | PTPRD | PTPRD | P23468 | -0.26925 | 0.035018 | 3.75939937411553e-14 | 6.83140843946026e-12 | -0.34069 | 0.017997 | 1.13e-74 | 6.79738461538461e-73 |
| 8885-6 | SL018710 | CA2D3 | CACNA2D3 | Q8IZS8 | -0.27282 | 0.035483 | 3.7597184587013e-14 | 6.83140843946026e-12 | -0.33923 | 0.018791 | 1.77e-68 | 8.142e-67 |
| 13690-26 | SL007804 | BGN | BGN | P21810 | -0.27265 | 0.035539 | 4.25868909426189e-14 | 7.54930544807205e-12 | -0.34757 | 0.019986 | 6e-64 | 1.955e-62 |
| 3298-52 | SL010454 | Contactin-4 | CNTN4 | Q8IWV2 | -0.27502 | 0.036138 | 6.67843823560434e-14 | 1.15568783562791e-11 | -0.19674 | 0.020111 | 3.4e-22 | 1.18168888888889e-21 |
| 5353-89 | SL001990 | IL-1Ra | IL1RN | P18510 | 0.276106 | 0.036361 | 7.54681856874177e-14 | 1.27558784552594e-11 | 0.307251 | 0.0182 | 1.47e-60 | 3.96393103448276e-59 |
| 10970-3 | SL018330 | NAR3 | ART3 | Q13508 | -0.23782 | 0.031664 | 1.37366266033064e-13 | 2.26904095801888e-11 | -0.28969 | 0.018265 | 5.44e-54 | 1.09078974358974e-52 |
| 10041-3 | SL007226 | HNF4A | HNF4A | P41235 | 0.270184 | 0.036154 | 1.80088347834405e-13 | 2.9086269156899e-11 | 0.182062 | 0.021003 | 7.85e-18 | 2.01268852459016e-17 |
| 7970-315 | SL018330 | NAR3 | ART3 | Q13508 | -0.23959 | 0.032088 | 1.88337563193086e-13 | 2.97573349845076e-11 | -0.29852 | 0.018758 | 2.49e-54 | 5.12415789473684e-53 |
| 3601-54 | SL008936 | CHL1 | CHL1 | O00533 | -0.26886 | 0.036171 | 2.39903068871356e-13 | 3.70982022246174e-11 | -0.27388 | 0.021057 | 1.93e-37 | 1.58869473684211e-36 |
| 8660-33 | SL009409 | OLFL3 | OLFML3 | Q9NRN5 | 0.259637 | 0.035401 | 4.84363028206949e-13 | 7.33406351876689e-11 | 0.290605 | 0.020213 | 4.73e-45 | 5.87120634920635e-44 |
| 2974-61 | SL004855 | contactin-1 | CNTN1 | Q12860 | -0.26293 | 0.035888 | 5.1109125263829e-13 | 7.58083923301039e-11 | -0.26894 | 0.020147 | 2.77e-39 | 2.40682222222222e-38 |
| 6507-16 | SL006401 | NCAM2 | NCAM2 | O15394 | -0.26563 | 0.036362 | 5.94349742348769e-13 | 8.6394678547817e-11 | -0.30719 | 0.019565 | 5.4e-53 | 1.00542857142857e-51 |
| 9769-48 | SL025836 | DNER:ECD | DNER | Q8NFT8 | -0.25786 | 0.035343 | 6.3374378107903e-13 | 8.94847662529797e-11 | -0.25974 | 0.019387 | 1.51e-39 | 1.34184090909091e-38 |
| 7957-2 | SL012750 | SCG3 | SCG3 | Q8WXD2 | -0.25157 | 0.034488 | 6.40232229658083e-13 | 8.94847662529797e-11 | -0.30197 | 0.018604 | 2.44e-56 | 5.45165714285714e-55 |
| 2950-57 | SL005171 | IGFBP-4 | IGFBP4 | P22692 | 0.259699 | 0.035785 | 8.30996299048372e-13 | 1.13956247197803e-10 | 0.219342 | 0.019036 | 5.97e-30 | 3.17587755102041e-29 |
| 13416-8 | SL020086 | T132D | TMEM132D | Q14C87 | -0.25298 | 0.034887 | 8.66903048139593e-13 | 1.16678728775529e-10 | -0.28175 | 0.018763 | 8.2e-49 | 1.33591666666667e-47 |
| 15539-15 | SL008760 | SLIK1 | SLITRK1 | Q96PX8 | -0.2524 | 0.034887 | 9.72522332847471e-13 | 1.28514405729735e-10 | -0.24602 | 0.017979 | 4.05e-41 | 3.91e-40 |
| 2765-4 | SL021043 | GDF-11/8 | GDF11\|MSTN | O95390\|O14793 | -0.22462 | 0.031093 | 1.04749899630893e-12 | 1.35950405449523e-10 | -0.21929 | 0.017613 | 1.53e-34 | 1.02261538461538e-33 |
| 6408-2 | SL007288 | INHBC | INHBC | P55103 | 0.256648 | 0.035724 | 1.3823293569792e-12 | 1.72696298634989e-10 | 0.401717 | 0.019597 | 2.77e-86 | 3.61023333333333e-84 |
| 14618-26 | SL012766 | ECOP | VOPP1 | Q96AW1 | -0.25793 | 0.035904 | 1.38347532108746e-12 | 1.72696298634989e-10 | -0.2691 | 0.019019 | 9.77e-44 | 1.09144857142857e-42 |
| 5358-3 | SL008574 | OMD | OMD | Q99983 | -0.25802 | 0.035926 | 1.40190996415305e-12 | 1.72696298634989e-10 | -0.22149 | 0.020699 | 3.81e-26 | 1.72220809248555e-25 |
| 6556-5 | SL012863 | ENPP5 | ENPP5 | Q9UJA9 | -0.23082 | 0.032178 | 1.49292880596854e-12 | 1.80843442696322e-10 | -0.2853 | 0.019646 | 6.88e-46 | 9.11891525423729e-45 |
| 10464-6 | SL008696 | ANTR1 | ANTXR1 | Q9H6X2 | -0.25787 | 0.036038 | 1.68792313854332e-12 | 2.01111891326768e-10 | -0.33695 | 0.01936 | 4.85e-64 | 1.649e-62 |
| 5532-53 | SL003060 | bFGF-R | FGFR1 | P11362 | -0.25544 | 0.035905 | 2.23734947263693e-12 | 2.62275096243955e-10 | -0.22445 | 0.020271 | 7.77e-28 | 3.8456582278481e-27 |
| 9793-145 | SL019123 | IGDC4 | IGDCC4 | Q8TDY8 | -0.23145 | 0.032594 | 2.45599028762678e-12 | 2.83335514451927e-10 | -0.29289 | 0.019896 | 4.64e-47 | 6.71940740740741e-46 |
| 21887-2 | SL012665 | CBLN2 | CBLN2 | Q8IUK8 | -0.25675 | 0.03621 | 2.6364862060857e-12 | 2.99405964778607e-10 | -0.23205 | 0.020795 | 3.1e-28 | 1.55397435897436e-27 |
| 10025-1 | SL008018 | DLDH | DLD | P09622 | -0.25218 | 0.035706 | 3.19103095747443e-12 | 3.56806353829603e-10 | -0.29082 | 0.020703 | 3.68e-43 | 3.99688888888889e-42 |
| 3438-10 | SL009324 | FSTL3 | FSTL3 | O95633 | 0.203056 | 0.029 | 4.82657046624895e-12 | 5.31507790131778e-10 | 0.262279 | 0.018271 | 6.38e-45 | 7.7955625e-44 |
| 16307-22 | SL005231 | UNC5H4 | UNC5D | Q6UXZ4 | -0.23935 | 0.034246 | 5.26047154186928e-12 | 5.70643390541879e-10 | -0.28369 | 0.018823 | 3.79e-49 | 6.443e-48 |
| 2925-9 | SL000006 | PAI-1 | SERPINE1 | P05121 | 0.248548 | 0.035651 | 5.92409825954032e-12 | 6.33181561034398e-10 | 0.263073 | 0.021098 | 1.22e-34 | 8.36877192982456e-34 |
| 8352-26 | SL025985 | SIG12:Ig-like C2-type 2 | SIGLEC12 | Q96PQ1 | 0.248232 | 0.035826 | 7.90331796805956e-12 | 8.3248282596894e-10 | 0.274063 | 0.02009 | 6.91e-41 | 6.3572e-40 |
| 25413-80 | SL004257 | GAGE-2 | GAGE2B | Q13066 | 0.24428 | 0.035415 | 9.74663884439572e-12 | 1.01197958744383e-09 | 0.26044 | 0.020977 | 2.31e-34 | 1.49290909090909e-33 |
| 5725-1 | SL008574 | OMD | OMD | Q99983 | -0.24647 | 0.035928 | 1.25131307622412e-11 | 1.28092161098547e-09 | -0.21734 | 0.020478 | 9.3e-26 | 4.08573033707865e-25 |
| 9277-16 | SL002648 | PAP1 | REG3A | Q06141 | -0.24965 | 0.036472 | 1.38505642530362e-11 | 1.39671719762144e-09 | -0.23678 | 0.020287 | 1.15e-30 | 6.37801418439716e-30 |
| 5307-12 | SL004400 | Coagulation Factor IXab | F9 | P00740 | 0.251261 | 0.036718 | 1.40286675050035e-11 | 1.39671719762144e-09 | 0.343694 | 0.019974 | 1.02e-62 | 3.06784615384615e-61 |
| 5483-1 | SL010467 | RGMA | RGMA | Q96B86 | -0.24512 | 0.035861 | 1.47732440620254e-11 | 1.45097213301082e-09 | -0.23598 | 0.019689 | 3.35e-32 | 2.01515384615385e-31 |
| 7210-25 | SL004470 | Amyloid-like protein 1 | APLP1 | P51693 | -0.24142 | 0.03547 | 1.78912804910362e-11 | 1.73378435478468e-09 | -0.28547 | 0.01982 | 3.34e-45 | 4.28177049180328e-44 |
| 21883-17 | SL003992 | BMP-5 | BMP5 | P22003 | -0.24476 | 0.036125 | 2.19670614508168e-11 | 2.10074477137548e-09 | -0.21417 | 0.021022 | 6.73e-24 | 2.60537623762376e-23 |
| 22533-95 | SL022816 | PP1RA | PPP1R10 | Q96QC0 | 0.240683 | 0.035646 | 2.56000195074529e-11 | 2.4163758672749e-09 | 0.289814 | 0.019335 | 1.19e-48 | 1.89914285714286e-47 |
| 18896-23 | SL021212 | H6ST3 | HS6ST3 | Q8IZP7 | -0.23573 | 0.035072 | 3.13575611720956e-11 | 2.91473546607927e-09 | -0.31166 | 0.018673 | 2.66e-59 | 6.71006451612903e-58 |
| 14337-1 | SL019699 | TPPC3 | TRAPPC3 | O43617 | 0.23929 | 0.035611 | 3.1681907239992e-11 | 2.91473546607927e-09 | 0.2126 | 0.017495 | 4.92e-33 | 3.05352380952381e-32 |
| 15667-39 | SL003996 | BMP-4 | BMP4 | P12644 | -0.24639 | 0.036694 | 3.26838019755194e-11 | 2.96932340947594e-09 | -0.22009 | 0.020722 | 8.6e-26 | 3.79954802259887e-25 |
| 13124-20 | SL018587 | ISLR2 | ISLR2 | Q6UXK2 | -0.24537 | 0.036668 | 3.80439735254915e-11 | 3.41362468621324e-09 | -0.24886 | 0.021068 | 2.37e-31 | 1.38308955223881e-30 |
| 2212-69 | SL000053 | tPA | PLAT | P00750 | 0.23032 | 0.0346 | 4.77363290823044e-11 | 4.23106877768523e-09 | 0.318839 | 0.020257 | 3.08e-53 | 6.0214e-52 |
| 4876-32 | SL000357 | Coagulation Factor IX | F9 | P00740 | 0.244256 | 0.03677 | 5.22225180495243e-11 | 4.57293085763786e-09 | 0.29875 | 0.016935 | 1.21e-65 | 4.98010526315789e-64 |
| 8900-28 | SL007582 | NEO1 | NEO1 | Q92859 | -0.23366 | 0.035512 | 7.85210885553264e-11 | 6.76267349625619e-09 | -0.15883 | 0.020922 | 4.47e-14 | 9.00912371134021e-14 |
| 5140-56 | SL005231 | UNC5H4 | UNC5D | Q6UXZ4 | -0.22548 | 0.034288 | 8.03267190672285e-11 | 6.76267349625619e-09 | -0.26322 | 0.018592 | 8.66e-44 | 9.81466666666667e-43 |
| 11278-4 | SL009488 | COL11A2 | COL11A2 | P13942 | -0.23804 | 0.0362 | 8.04752237192926e-11 | 6.76267349625619e-09 | -0.15847 | 0.021261 | 1.26e-13 | 2.50081218274112e-13 |
| 3198-4 | SL008639 | IDS | IDS | P22304 | -0.24113 | 0.036674 | 8.09510999139087e-11 | 6.76267349625619e-09 | -0.17326 | 0.019969 | 7.33e-18 | 1.88554605263158e-17 |
| 12488-9 | SL015331 | MCTS1 | MCTS1 | Q9ULC4 | 0.238572 | 0.036481 | 1.01459337591271e-10 | 8.37961892742451e-09 | 0.297772 | 0.020604 | 1.74e-45 | 2.2678e-44 |
| 8469-41 | SL000466 | IGFBP-2 | IGFBP2 | P18065 | -0.18744 | 0.028768 | 1.18321646045068e-10 | 9.66249127478149e-09 | -0.20183 | 0.019291 | 4.34e-25 | 1.81490909090909e-24 |
| 11208-15 | SL019369 | NAGPA | NAGPA | Q9UK23 | -0.22763 | 0.034948 | 1.19898153527145e-10 | 9.68244199816992e-09 | -0.24965 | 0.020101 | 2.18e-34 | 1.43257142857143e-33 |
| 7049-2 | SL015911 | ADAM 23 | ADAM23 | O75077 | -0.2391 | 0.036775 | 1.29077620930327e-10 | 1.03091884496881e-08 | -0.24133 | 0.020985 | 7.63e-30 | 4.03152702702703e-29 |
| 6914-15 | SL015046 | AMGO2 | AMIGO2 | Q86SJ2 | -0.23842 | 0.03694 | 1.74646295506033e-10 | 1.37970573449766e-08 | -0.22589 | 0.021359 | 1.37e-25 | 5.91900552486188e-25 |
| 5029-3 | SL006528 | SEPR | FAP | Q12884 | -0.22712 | 0.035203 | 1.7731978966628e-10 | 1.385763689564e-08 | -0.20297 | 0.020841 | 5.14e-22 | 1.7629298245614e-21 |
| 13697-51 | SL007151 | GPDA | GPD1 | P21695 | 0.232528 | 0.036086 | 1.8659862481047e-10 | 1.44276468630053e-08 | 0.263588 | 0.020587 | 2.27e-36 | 1.74033333333333e-35 |
| 22985-160 | SL000466 | IGFBP-2 | IGFBP2 | P18065 | -0.17813 | 0.027677 | 1.95790011334937e-10 | 1.49789663408665e-08 | -0.19499 | 0.019176 | 8.15e-24 | 3.13955665024631e-23 |
| 10746-24 | SL009412 | DKK3 | DKK3 | Q9UBP4 | -0.21444 | 0.033337 | 2.00362752853194e-10 | 1.51691300805939e-08 | -0.28195 | 0.020298 | 2.82e-42 | 2.86394805194805e-41 |
| 8819-3 | SL000466 | IGFBP-2 | IGFBP2 | P18065 | -0.18371 | 0.02858 | 2.05799563385013e-10 | 1.54201157389925e-08 | -0.18764 | 0.019378 | 8.72e-22 | 2.86514285714286e-21 |
| 20579-50 | SL008643 | MAG | MAG | P20916 | -0.22742 | 0.035478 | 2.30389411508145e-10 | 1.70864310494e-08 | -0.29603 | 0.018905 | 9.8e-53 | 1.78223255813953e-51 |
| 8262-20 | SL005361 | Apo D | APOD | P05090 | -0.22903 | 0.035792 | 2.47515281050106e-10 | 1.81711218451734e-08 | -0.37825 | 0.019211 | 3.27e-80 | 2.55714e-78 |
| 17366-6 | SL010950 | DCNL1 | DCUN1D1 | Q96GG9 | 0.226942 | 0.035512 | 2.60748583856051e-10 | 1.89512070746578e-08 | 0.33229 | 0.019807 | 7.31e-60 | 1.90547333333333e-58 |
| 18917-53 | SL005382 | Pancreatic alpha-amylase | AMY2A | P04746 | -0.23203 | 0.036368 | 2.78375292015249e-10 | 2.00319962610577e-08 | -0.25432 | 0.020739 | 1.37e-33 | 8.63983870967742e-33 |
| 3607-71 | SL009412 | DKK3 | DKK3 | Q9UBP4 | -0.21315 | 0.03345 | 2.92213297755962e-10 | 2.08216298832386e-08 | -0.28105 | 0.020121 | 1.04e-42 | 1.11408219178082e-41 |
| 2570-72 | SL000466 | IGFBP-2 | IGFBP2 | P18065 | -0.18321 | 0.028785 | 3.06192686485497e-10 | 2.16059072366659e-08 | -0.18012 | 0.019412 | 3.67e-20 | 1.06294074074074e-19 |
| 21698-11 | SL022706 | Integrin a11b1 | ITGA11\|ITGB1 | Q9UKX5\|P05556 | -0.23377 | 0.036825 | 3.39888738340285e-10 | 2.3752993752473e-08 | -0.26311 | 0.020399 | 7.19e-37 | 5.79647422680412e-36 |
| 8528-74 | SL018587 | ISLR2 | ISLR2 | Q6UXK2 | -0.23273 | 0.036769 | 3.8136858710149e-10 | 2.6397970390987e-08 | -0.24296 | 0.021078 | 5.71e-30 | 3.0583698630137e-29 |
| 3736-60 | SL003993 | BMP-6 | BMP6 | P22004 | -0.23281 | 0.036852 | 4.11136462555873e-10 | 2.81899982061895e-08 | -0.17437 | 0.020781 | 8.05e-17 | 1.96109034267913e-16 |
| 13460-4 | SL012363 | CHAD | CHAD | O15335 | -0.22908 | 0.036281 | 4.19511484353201e-10 | 2.84954155913931e-08 | -0.19454 | 0.021182 | 8.6e-20 | 2.42787003610108e-19 |
| 11214-40 | SL019363 | DNJB9 | DNAJB9 | Q9UBS3 | 0.225935 | 0.035859 | 4.56396224803e-10 | 3.07137755728538e-08 | 0.287272 | 0.020606 | 1.46e-42 | 1.50226315789474e-41 |
| 9416-77 | SL008646 | CBPM | CPM | P14384 | 0.221784 | 0.035243 | 4.78395178397912e-10 | 3.18988638219819e-08 | 0.196045 | 0.020289 | 1.06e-21 | 3.4395020746888e-21 |
| 24671-15 | SL007810 | ASC | PYCARD | Q9ULZ3 | 0.216743 | 0.034709 | 6.44648512304809e-10 | 4.2593685340285e-08 | 0.24796 | 0.019694 | 2.89e-35 | 2.05452727272727e-34 |
| 9360-33 | SL012367 | EDIL3 | EDIL3 | O43854 | -0.22617 | 0.036381 | 7.6422499984657e-10 | 5.0039525215179e-08 | -0.25919 | 0.020576 | 2.7e-35 | 1.93706422018349e-34 |
| 10439-57 | SL005393 | Alpha-amylase 2B | AMY2B | P19961 | -0.22496 | 0.036288 | 8.50867455268439e-10 | 5.52152202222412e-08 | -0.27828 | 0.020671 | 6.69e-40 | 6.01331034482759e-39 |
| 6388-21 | SL017451 | CC126 | CCDC126 | Q96EE4 | -0.22546 | 0.036547 | 1.02179285129362e-09 | 6.57202694088675e-08 | -0.29704 | 0.020026 | 1.01e-47 | 1.49022641509434e-46 |
| 2999-6 | SL005196 | LSAMP | LSAMP | Q13449 | -0.20866 | 0.03405 | 1.31107196375623e-09 | 8.35865880050902e-08 | -0.24394 | 0.018776 | 2.28e-37 | 1.85725e-36 |
| 14125-5 | SL004747 | ApoM | APOM | O95445 | -0.21493 | 0.035294 | 1.65025777217425e-09 | 1.04296291201413e-07 | -0.28677 | 0.019676 | 3.38e-46 | 4.5571724137931e-45 |
| 8356-88 | SL009326 | NEU1 | OXT | P01178 | 0.224529 | 0.036923 | 1.74030785793683e-09 | 1.09039288892111e-07 | 0.264177 | 0.020497 | 8.1e-37 | 6.39818181818182e-36 |
| 21796-43 | SL001886 | Glutathione peroxidase 3 | GPX3 | P22352 | -0.22022 | 0.03626 | 1.82135427149675e-09 | 1.13141904660157e-07 | -0.2362 | 0.02013 | 5.73e-31 | 3.29475e-30 |
| 7228-2 | SL018113 | SIA7F | ST6GALNAC6 | Q969X2 | -0.21585 | 0.035677 | 2.0941656846473e-09 | 1.28986408440818e-07 | -0.23229 | 0.018705 | 2.21e-34 | 1.44018333333333e-33 |
| 15556-49 | SL005393 | Alpha-amylase 2B | AMY2B | P19961 | -0.21562 | 0.035657 | 2.13029746529827e-09 | 1.30109260317545e-07 | -0.2589 | 0.020087 | 8.05e-37 | 6.39818181818182e-36 |
| 13524-25 | SL020161 | H6ST2 | HS6ST2 | Q96MM7 | -0.21632 | 0.035898 | 2.41582789783199e-09 | 1.46318643012024e-07 | -0.17348 | 0.020925 | 1.84e-16 | 4.32096096096096e-16 |
| 19553-14 | SL004304 | STX1a | STX1A | Q16623 | -0.20383 | 0.033968 | 2.81094045328658e-09 | 1.68842274499891e-07 | -0.2141 | 0.018995 | 9.22e-29 | 4.68184415584416e-28 |
| 13117-232 | SL018256 | CHKB | CHKB | Q9Y259 | 0.216493 | 0.036385 | 3.78865213532867e-09 | 2.25704292783351e-07 | 0.227281 | 0.021245 | 3.9e-26 | 1.75275862068966e-25 |
| 11266-8 | SL025980 | SELPL:ECD | SELPLG | Q14242 | -0.21062 | 0.035472 | 4.08059853324987e-09 | 2.41120245037887e-07 | -0.16243 | 0.019936 | 5.86e-16 | 1.32442774566474e-15 |
| 4929-55 | SL005102 | SHBG | SHBG | P04278 | -0.20141 | 0.033976 | 4.30769492540667e-09 | 2.50985234859297e-07 | -0.24222 | 0.019349 | 6.81e-35 | 4.75483928571429e-34 |
| 7768-10 | SL008646 | CBPM | CPM | P14384 | 0.202527 | 0.034165 | 4.3166145235845e-09 | 2.50985234859297e-07 | 0.167194 | 0.01727 | 8.91e-22 | 2.91532217573222e-21 |
| 8264-43 | SL006970 | DLL1 | DLL1 | O00548 | 0.205253 | 0.034635 | 4.35904447010655e-09 | 2.51440755624876e-07 | 0.180802 | 0.019413 | 2.67e-20 | 7.90886363636364e-20 |
| 4328-2 | SL013490 | BOC | BOC | Q9BWV1 | -0.216 | 0.036497 | 4.56482088499577e-09 | 2.61237151119285e-07 | -0.16948 | 0.020751 | 5.01e-16 | 1.14556140350877e-15 |
| 3316-58 | SL004466 | Heparin cofactor II | SERPIND1 | P05546 | 0.210942 | 0.035655 | 4.62156832911967e-09 | 2.62418426687826e-07 | 0.204027 | 0.020971 | 5.66e-22 | 1.91455172413793e-21 |
| 8312-139 | SL007871 | Cytidylate kinase | CMPK1 | P30085 | 0.217712 | 0.036839 | 4.79716640659732e-09 | 2.70037711293895e-07 | 0.149297 | 0.017865 | 1.07e-16 | 2.5588379204893e-16 |
| 11192-168 | SL008302 | TINAL | TINAGL1 | Q9GZM7 | -0.2144 | 0.036286 | 4.83006363073836e-09 | 2.70037711293895e-07 | -0.23768 | 0.020928 | 3.64e-29 | 1.86044444444444e-28 |
| 13747-9 | SL010288 | Carbonic anhydrase 6 | CA6 | P23280 | -0.2 | 0.034006 | 5.66645130495982e-09 | 3.14379909041587e-07 | -0.23521 | 0.019752 | 8.03e-32 | 4.75716666666667e-31 |
| 8997-4 | SL008970 | NPTXR | NPTXR | O95502 | -0.20769 | 0.035589 | 7.38064346503797e-09 | 4.06382702302242e-07 | -0.26578 | 0.018746 | 6.66e-44 | 7.659e-43 |
| 8280-238 | SL000616 | Vitronectin | VTN | P04004 | 0.207098 | 0.035678 | 8.83568666255022e-09 | 4.82840380927932e-07 | 0.233066 | 0.020455 | 2.44e-29 | 1.27205333333333e-28 |
| 15384-15 | SL011369 | KLOTHO | KL | Q9UEF7 | -0.21287 | 0.036718 | 9.20328203551787e-09 | 4.99175028613014e-07 | -0.15128 | 0.021745 | 4.45e-12 | 8.05532407407407e-12 |
| 2677-1 | SL002644 | ERBB1 | EGFR | P00533 | -0.18522 | 0.031973 | 9.44246017385584e-09 | 5.08354078100624e-07 | -0.26563 | 0.020793 | 3.18e-36 | 2.41433009708738e-35 |
| 3331-8 | SL010468 | RGMB | RGMB | Q6NW40 | -0.20153 | 0.034846 | 9.97486064256314e-09 | 5.33068287868742e-07 | -0.19887 | 0.019587 | 9.44e-24 | 3.60101463414634e-23 |
| 8364-74 | SL006797 | UST | UST | Q9Y2C2 | -0.2114 | 0.036637 | 1.07643680787048e-08 | 5.71061512379755e-07 | -0.2327 | 0.019852 | 6.57e-31 | 3.75017518248175e-30 |
| 11196-31 | SL025822 | Collagen alpha-3(VI):BPTI/Kunitz inhibitor | COL6A3 | P12111 | 0.192516 | 0.033546 | 1.2879586072006e-08 | 6.78324866458981e-07 | 0.211801 | 0.015918 | 4.78e-39 | 4.063e-38 |
| 7853-19 | SL018291 | SCO1 | SCO1 | O75880 | -0.20911 | 0.03649 | 1.3514393797388e-08 | 7.06637511650474e-07 | -0.17891 | 0.017084 | 3.92e-25 | 1.64808602150538e-24 |
| 12727-7 | SL019877 | FPRP | PTGFRN | Q9P2B2 | -0.20158 | 0.035289 | 1.4984478451614e-08 | 7.77908495616644e-07 | -0.20968 | 0.019931 | 2.41e-25 | 1.03550549450549e-24 |
| 10036-201 | SL019252 | ZHX3 | ZHX3 | Q9H4I2 | 0.204425 | 0.035855 | 1.59354568076288e-08 | 8.21410638849975e-07 | 0.11169 | 0.018067 | 7.4e-10 | 1.1690505050505e-09 |
| 3344-60 | SL000272 | Antithrombin III | SERPINC1 | P01008 | -0.20638 | 0.036323 | 1.78037207336619e-08 | 9.11249593607426e-07 | -0.2407 | 0.020882 | 5.69e-30 | 3.0583698630137e-29 |
| 21581-87 | SL021164 | CDK15 | CDK15 | Q96Q40 | 0.204997 | 0.036272 | 2.11083531711156e-08 | 1.07283574019348e-06 | 0.063124 | 0.021263 | 0.00302 | 0.003378039 |
| 15511-37 | SL008970 | NPTXR | NPTXR | O95502 | -0.20351 | 0.036031 | 2.15329383130617e-08 | 1.08681524763425e-06 | -0.27345 | 0.019261 | 5.13e-44 | 5.98755223880597e-43 |
| 16907-3 | SL004806 | Nectin-like protein 3 | CADM2 | Q8N3J6 | -0.1954 | 0.034632 | 2.22585537886176e-08 | 1.10999156445795e-06 | -0.25789 | 0.018636 | 5.55e-42 | 5.49379746835443e-41 |
| 5982-50 | SL002690 | FHR1 | CFHR1 | Q03591 | 0.205861 | 0.036488 | 2.22975740796451e-08 | 1.10999156445795e-06 | 0.224808 | 0.021233 | 1.22e-25 | 5.30022222222222e-25 |
| 6462-12 | SL004789 | TIMP-4 | TIMP4 | Q99727 | 0.17127 | 0.030487 | 2.55152699601372e-08 | 1.261530490274e-06 | 0.15266 | 0.019688 | 1.3e-14 | 2.69655172413793e-14 |
| 10440-26 | SL025782 | ACAM:ECD | CLMP | Q9H6B4 | 0.193297 | 0.034441 | 2.63069309314447e-08 | 1.29188360817392e-06 | 0.156792 | 0.016531 | 5.55e-21 | 1.75004032258065e-20 |
| 4324-33 | SL008516 | CYTT | CST2 | P09228 | -0.19462 | 0.034685 | 2.64881517574236e-08 | 1.29205293270439e-06 | -0.11181 | 0.020867 | 9.19e-08 | 1.30903096539162e-07 |
| 3805-16 | SL010458 | Endocan | ESM1 | Q9NQ30 | -0.19314 | 0.034562 | 3.01022361147571e-08 | 1.45505963991901e-06 | -0.21612 | 0.019853 | 5.55e-27 | 2.63036363636364e-26 |
| 9185-15 | SL008591 | TFF1 | TFF1 | P04155 | -0.20259 | 0.036261 | 3.03064871949414e-08 | 1.45505963991901e-06 | -0.18413 | 0.021464 | 1.68e-17 | 4.25165048543689e-17 |
| 19768-13 | SL000395 | Cystatin B | CSTB | P04080 | 0.191931 | 0.034357 | 3.0430526316413e-08 | 1.45505963991901e-06 | 0.220889 | 0.017962 | 9.12e-34 | 5.84577049180328e-33 |
| 2474-54 | SL000573 | SAP | APCS | P02743 | 0.186431 | 0.033535 | 3.53700613508655e-08 | 1.6801935026019e-06 | 0.279664 | 0.01901 | 5.32e-47 | 7.56407272727273e-46 |
| 4129-72 | SL000414 | Factor B | CFB | P00751 | 0.194332 | 0.035022 | 3.74679983085397e-08 | 1.76829488121082e-06 | 0.251984 | 0.018451 | 5.69e-41 | 5.29711904761905e-40 |
| 15491-20 | SL008437 | CD248 | CD248 | Q9HCU0 | -0.19562 | 0.035312 | 3.93982177002884e-08 | 1.84181928212127e-06 | -0.11926 | 0.021193 | 2.04e-08 | 3.02136363636364e-08 |
| 7909-37 | SL005102 | SHBG | SHBG | P04278 | -0.19224 | 0.034706 | 3.95327198694165e-08 | 1.84181928212127e-06 | -0.24252 | 0.019174 | 1.45e-35 | 1.05971962616822e-34 |
| 4159-130 | SL000415 | Factor H | CFH | P08603 | 0.20271 | 0.036632 | 4.07525129536026e-08 | 1.88655582259098e-06 | 0.284269 | 0.01996 | 2.86e-44 | 3.38866666666667e-43 |
| 2687-2 | SL001947 | MIA | MIA | Q16674 | -0.19625 | 0.035492 | 4.16912781234891e-08 | 1.9177987936805e-06 | -0.22226 | 0.020469 | 7.41e-27 | 3.46983233532934e-26 |
| 6525-17 | SL012566 | DUSP13 | DUSP13 | Q6B8I1 | -0.2035 | 0.036832 | 4.27291252344154e-08 | 1.95317787549516e-06 | -0.12842 | 0.018867 | 1.26e-11 | 2.199375e-11 |
| 8888-33 | SL018720 | CX068 | SMIM9 | A6NGZ8 | 0.20193 | 0.036616 | 4.52032206305368e-08 | 2.05335629714213e-06 | 0.108391 | 0.019793 | 4.79e-08 | 6.96241635687732e-08 |
| 4979-34 | SL008178 | DERM | DPT | Q07507 | 0.177877 | 0.032264 | 4.56125145379596e-08 | 2.05907922771361e-06 | 0.169039 | 0.020628 | 4.01e-16 | 9.25020648967552e-16 |
| 2567-5 | SL003328 | Factor I | CFI | P05156 | 0.203093 | 0.036906 | 4.82491278885142e-08 | 2.16465840428223e-06 | 0.259724 | 0.020854 | 1.46e-34 | 9.84241379310345e-34 |
| 4883-56 | SL000021 | Insulin | INS | P01308 | 0.200473 | 0.036541 | 5.29096403082292e-08 | 2.34680057449904e-06 | 0.206725 | 0.020114 | 2.76e-24 | 1.10517346938776e-23 |
| 9830-109 | SL019151 | VAV3 | VAV3 | Q9UKW4 | 0.200887 | 0.036618 | 5.29547735577659e-08 | 2.34680057449904e-06 | 0.212404 | 0.020279 | 3.85e-25 | 1.62740540540541e-24 |
| 17678-28 | SL006796 | VSIG4 | VSIG4 | Q9Y279 | 0.167356 | 0.030529 | 5.42039302254721e-08 | 2.38760099926504e-06 | 0.248453 | 0.019598 | 1.03e-35 | 7.59867924528302e-35 |
| 13133-73 | SL007033 | LTBP4 | LTBP4 | Q8N2S1 | -0.17888 | 0.032698 | 5.75983140413621e-08 | 2.52183461718446e-06 | -0.1947 | 0.020008 | 5.55e-22 | 1.887e-21 |
| 7871-16 | SL008988 | T132A | TMEM132A | Q24JP5 | -0.1992 | 0.036422 | 5.80612363855934e-08 | 2.52688063503289e-06 | -0.18889 | 0.020329 | 3.25e-20 | 9.44795539033457e-20 |
| 12987-12 | SL019472 | SRSF7 | SRSF7 | Q16629 | 0.194958 | 0.035705 | 6.09716441957884e-08 | 2.63774946437494e-06 | 0.219071 | 0.018026 | 4.89e-33 | 3.05352380952381e-32 |
| 10702-1 | SL012521 | COSA1 | COL28A1 | Q2UY09 | 0.181231 | 0.033225 | 6.28769320839244e-08 | 2.70408013246132e-06 | 0.228314 | 0.017131 | 3.62e-39 | 3.11081318681319e-38 |
| 9018-38 | SL025955 | PCD10:ECD | PCDH10 | Q9P2E7 | -0.17477 | 0.032103 | 6.66020741951373e-08 | 2.84743456029563e-06 | -0.24551 | 0.017972 | 5.41e-41 | 5.09713253012048e-40 |
| 15453-3 | SL003022 | a1-Microglobulin | AMBP | P02760 | 0.186964 | 0.034365 | 6.79099823390257e-08 | 2.88637281660842e-06 | 0.184295 | 0.020686 | 9.84e-19 | 2.66258823529412e-18 |
| 22966-20 | SL020978 | GBRAP | GABARAP | O95166 | 0.180396 | 0.033224 | 7.20393115758962e-08 | 3.04407974728845e-06 | 0.27406 | 0.019636 | 1.2e-42 | 1.2512e-41 |
| 8275-31 | SL025956 | PEAR1:ECD | PEAR1 | Q5VY43 | -0.19543 | 0.036008 | 7.29356097548602e-08 | 3.06413879594407e-06 | -0.13597 | 0.02053 | 4.31e-11 | 7.24821505376344e-11 |
| 16908-5 | SL010377 | OMGP | OMG | P23515 | -0.19515 | 0.035992 | 7.5010176188628e-08 | 3.13318368125832e-06 | -0.26462 | 0.019951 | 8.06e-39 | 6.77733333333333e-38 |
| 7859-21 | SL008367 | PCDGK | PCDHGC3 | Q9UN70 | 0.194143 | 0.036288 | 1.10620156213368e-07 | 4.59421311633575e-06 | 0.196916 | 0.021388 | 7.02e-20 | 1.99623272727273e-19 |
| 4232-19 | SL003304 | IGF-I sR | IGF1R | P08069 | -0.19414 | 0.036485 | 1.29193851918158e-07 | 5.33511883943846e-06 | -0.18474 | 0.020893 | 1.76e-18 | 4.64972972972973e-18 |
| 9005-16 | SL011810 | PLXA1 | PLXNA1 | Q9UIW2 | -0.19143 | 0.036084 | 1.40714709663278e-07 | 5.77804807815087e-06 | -0.10677 | 0.019869 | 8.45e-08 | 1.20802559414991e-07 |
| 7090-17 | SL018090 | GALT1 | GALNT1 | Q10472 | 0.193858 | 0.03657 | 1.43805941367526e-07 | 5.87180663965832e-06 | 0.040814 | 0.015669 | 0.00925 | 0.010216697 |
| 13463-1 | SL007573 | PXDN | PXDN | Q92626 | 0.182424 | 0.034643 | 1.73275093591859e-07 | 7.03554961019907e-06 | 0.301973 | 0.01737 | 6.55e-64 | 2.04884e-62 |
| 15483-377 | SL005368 | Agrin | AGRN | O00468 | 0.193809 | 0.036826 | 1.76087156890312e-07 | 7.1100080904377e-06 | 0.163624 | 0.021106 | 1.31e-14 | 2.70295514511873e-14 |
| 16070-7 | SL004652 | WIF-1 | WIF1 | Q9Y5W5 | -0.1949 | 0.037195 | 1.98723445527218e-07 | 7.97967956956808e-06 | -0.12815 | 0.019016 | 1.98e-11 | 3.4104845814978e-11 |
| 13933-276 | SL020257 | NAB1 | NAB1 | Q13506 | 0.188266 | 0.035944 | 2.00896353683011e-07 | 8.02260823389077e-06 | 0.188941 | 0.02001 | 8.21e-21 | 2.55785657370518e-20 |
| 5837-49 | SL004714 | LIF sR | LIFR | P42702 | -0.19134 | 0.036541 | 2.02537849852298e-07 | 8.04396225533608e-06 | -0.19125 | 0.020848 | 9.47e-20 | 2.65431541218638e-19 |
| 6904-14 | SL018013 | LRRT2 | LRRTM2 | O43300 | -0.18663 | 0.035771 | 2.23760811866456e-07 | 8.838552068725e-06 | -0.18758 | 0.018985 | 1.33e-22 | 4.74913242009132e-22 |
| 21770-18 | SL022629 | B3GA1 | B3GAT1 | Q9P2W7 | -0.18505 | 0.035496 | 2.2857368745968e-07 | 8.97985708355112e-06 | -0.20806 | 0.020666 | 2.18e-23 | 8.19596153846154e-23 |
| 5107-7 | SL005703 | Notch 1 | NOTCH1 | P46531 | -0.19222 | 0.036902 | 2.33693802026691e-07 | 9.13164813510747e-06 | -0.20246 | 0.02081 | 5.68e-22 | 1.91455172413793e-21 |
| 3323-37 | SL004610 | LRP8 | LRP8 | Q14114 | -0.19329 | 0.037151 | 2.41271316261889e-07 | 9.31553334199829e-06 | -0.13009 | 0.01848 | 2.5e-12 | 4.61084905660377e-12 |
| 9242-11 | SL000137 | BTC | BTC | P35070 | -0.18952 | 0.036431 | 2.42094270076136e-07 | 9.31553334199829e-06 | -0.22515 | 0.019052 | 2.23e-31 | 1.31117293233083e-30 |
| 3057-55 | SL004689 | WISP-1 | CCN4 | O95388 | 0.187954 | 0.03613 | 2.42244881898414e-07 | 9.31553334199829e-06 | 0.175321 | 0.021484 | 5.28e-16 | 1.20377842565598e-15 |
| 3074-6 | SL003309 | LBP | LBP | P18428 | 0.189412 | 0.03665 | 2.89210999525732e-07 | 1.1063081813437e-05 | 0.161799 | 0.020141 | 1.46e-15 | 3.22519774011299e-15 |
| 4568-17 | SL014070 | SLIK5 | SLITRK5 | O94991 | -0.18668 | 0.036151 | 2.9584802049536e-07 | 1.12577142039805e-05 | -0.19527 | 0.019914 | 2.74e-22 | 9.56553571428571e-22 |
| 11814-29 | SL000134 | Met | MET | P08581 | -0.1908 | 0.037049 | 3.17974354715322e-07 | 1.20366542191196e-05 | -0.19612 | 0.019754 | 8.48e-23 | 3.07007407407407e-22 |
| 6580-29 | SL004740 | Pregnancy zone protein | PZP | P20742 | 0.134708 | 0.026169 | 3.21662044147674e-07 | 1.20665519976643e-05 | 0.069914 | 0.014853 | 2.65e-06 | 3.48285714285714e-06 |
| 15626-223 | SL005400 | Perlecan | HSPG2 | P98160 | -0.19028 | 0.036966 | 3.220846295469e-07 | 1.20665519976643e-05 | -0.15779 | 0.019115 | 2.46e-16 | 5.74244776119403e-16 |
| 9595-11 | SL019028 | B4GT2 | B4GALT2 | O60909 | -0.1872 | 0.036513 | 3.58245100757128e-07 | 1.33524379092451e-05 | -0.27635 | 0.018393 | 7.27e-49 | 1.20960425531915e-47 |
| 16612-28 | SL020764 | SDC3 | SDC3 | O75056 | 0.18224 | 0.035617 | 3.77446778588535e-07 | 1.39963427897014e-05 | 0.216545 | 0.020435 | 1.11e-25 | 4.84927374301676e-25 |
| 19233-75 | SL006960 | ATOX1 | ATOX1 | O00244 | 0.168453 | 0.033015 | 4.06353308956448e-07 | 1.49917555811952e-05 | 0.206093 | 0.018907 | 4.79e-27 | 2.28401219512195e-26 |
| 18380-78 | SL000254 | Albumin | ALB | P02768 | -0.1731 | 0.033944 | 4.11959472776158e-07 | 1.5121825495642e-05 | -0.26933 | 0.019704 | 4.87e-41 | 4.64431707317073e-40 |
| 8305-18 | SL008928 | SULF2 | SULF2 | Q8IWU5 | 0.187827 | 0.03687 | 4.23702986477501e-07 | 1.54747402297411e-05 | 0.215462 | 0.018647 | 4.19e-30 | 2.29131468531469e-29 |
| 10833-64 | SL012788 | HHIP | HHIP | Q96QV1 | -0.18246 | 0.03584 | 4.30860568949681e-07 | 1.5626650947736e-05 | -0.19139 | 0.020425 | 1.59e-20 | 4.83805447470817e-20 |
| 20512-2 | SL002727 | MUC18 | MCAM | P43121 | -0.17277 | 0.033941 | 4.32162471174317e-07 | 1.5626650947736e-05 | -0.13071 | 0.021229 | 8.64e-10 | 1.35945271629779e-09 |
| 15620-4 | SL018410 | NLGN1 | NLGN1 | Q8N2Q7 | -0.1785 | 0.035087 | 4.39099188868887e-07 | 1.5798875765837e-05 | -0.1565 | 0.01967 | 2.69e-15 | 5.82709141274238e-15 |
| 16607-78 | SL005572 | Gelsolin | GSN | P06396 | -0.18654 | 0.03677 | 4.71992240466642e-07 | 1.68987172596628e-05 | -0.21291 | 0.02084 | 5.06e-24 | 1.97846e-23 |
| 7948-129 | SL010876 | GLTD2 | GLTPD2 | A6NH11 | 0.18043 | 0.03572 | 5.27905620685258e-07 | 1.88079316232375e-05 | 0.248549 | 0.019952 | 1.4e-34 | 9.52e-34 |
| 7991-54 | SL018364 | IQCF1 | IQCF1 | Q8N6M8 | 0.182283 | 0.036099 | 5.32450204076185e-07 | 1.88773077230522e-05 | 0.154525 | 0.021084 | 3.14e-13 | 6.06291358024691e-13 |
| 5105-2 | SL005208 | Nogo Receptor | RTN4R | Q9BZR6 | 0.180638 | 0.036 | 6.26213838095056e-07 | 2.19955034104121e-05 | 0.179577 | 0.021251 | 4.92e-17 | 1.20988679245283e-16 |
| 5695-5 | SL010290 | SPLC2 | BPIFA2 | Q96DR5 | -0.16752 | 0.033386 | 6.26454211056041e-07 | 2.19955034104121e-05 | -0.18319 | 0.019359 | 6.83e-21 | 2.14500401606426e-20 |
| 5015-15 | SL003440 | PAFAH | PLA2G7 | Q13093 | -0.17455 | 0.034836 | 6.48984043609316e-07 | 2.26770001391947e-05 | -0.19061 | 0.019886 | 2.18e-21 | 6.90186234817814e-21 |
| 5036-50 | SL004782 | TSG-6 | TNFAIP6 | P98066 | -0.16913 | 0.033957 | 7.54768508226333e-07 | 2.62471651568851e-05 | -0.20814 | 0.019594 | 8.52e-26 | 3.78559090909091e-25 |
| 15562-24 | SL008667 | BGLR | GUSB | P08236 | 0.179658 | 0.03609 | 7.64952826406279e-07 | 2.64746530586706e-05 | 0.232628 | 0.020165 | 5.14e-30 | 2.79130555555556e-29 |
| 13731-14 | SL000323 | C7 | C7 | P10643 | -0.17603 | 0.035418 | 7.96442356665097e-07 | 2.73057000523723e-05 | -0.12983 | 0.02011 | 1.29e-10 | 2.11928571428571e-10 |
| 3049-61 | SL000603 | Trypsin | PRSS1 | P07477 | -0.18532 | 0.037287 | 7.96478867790716e-07 | 2.73057000523723e-05 | -0.14826 | 0.021265 | 4.01e-12 | 7.29260465116279e-12 |
| 21707-15 | SL018449 | C1QL3 | C1QL3 | Q5VWW1 | -0.1811 | 0.036581 | 8.77299539057534e-07 | 2.98983309429789e-05 | -0.26792 | 0.017603 | 4.78e-50 | 8.30657777777778e-49 |
| 20530-2 | SL021501 | SEM6D | SEMA6D | Q8NFY4 | -0.18268 | 0.036906 | 8.80330602889033e-07 | 2.98983309429789e-05 | -0.0902 | 0.021788 | 3.59e-05 | 4.42107086614173e-05 |
| 8986-2 | SL018791 | SERA | PHGDH | O43175 | 0.182923 | 0.03698 | 8.95872330205548e-07 | 3.02373427611771e-05 | 0.127222 | 0.020456 | 5.86e-10 | 9.31406504065041e-10 |
| 16323-8 | SL020462 | NRX3A | NRXN3 | Q9Y4C0 | -0.17175 | 0.034725 | 8.98633191581487e-07 | 3.02373427611771e-05 | -0.21436 | 0.019392 | 9.44e-28 | 4.6138e-27 |
| 5657-28 | SL009528 | SIA4A | ST3GAL1 | Q11201 | -0.18432 | 0.037287 | 9.10069878047197e-07 | 3.04049682137401e-05 | -0.19245 | 0.018726 | 2.77e-24 | 1.10517346938776e-23 |
| 7735-17 | SL003066 | PEDF | SERPINF1 | P36955 | 0.173179 | 0.035035 | 9.11981710318565e-07 | 3.04049682137401e-05 | 0.230219 | 0.019715 | 1.07e-30 | 5.97671428571428e-30 |
| 15387-44 | SL005203 | Neuropilin-2 | NRP2 | O60462 | -0.17681 | 0.035828 | 9.49764766587384e-07 | 3.15200471395302e-05 | -0.09157 | 0.02056 | 8.81e-06 | 1.12756464811784e-05 |
| 16609-106 | SL012592 | KIRR2 | KIRREL2 | Q6UWL6 | -0.1807 | 0.036646 | 9.67865525454711e-07 | 3.1974757450022e-05 | -0.07652 | 0.017896 | 1.98e-05 | 2.4853290529695e-05 |
| 6605-17 | SL005608 | IGFALS | IGFALS | P35858 | -0.14869 | 0.030237 | 1.03578610716548e-06 | 3.40637711623471e-05 | -0.16518 | 0.019732 | 9.48e-17 | 2.28807407407407e-16 |
| 13501-10 | SL020163 | S35G2 | SLC35G2 | Q8TBE7 | 0.179329 | 0.036569 | 1.10834127054541e-06 | 3.62856952897478e-05 | 0.199902 | 0.021159 | 7.82e-21 | 2.446096e-20 |
| 6289-78 | SL017400 | RHG36 | ARHGAP36 | Q6ZRI8 | 0.179855 | 0.036703 | 1.12830492665202e-06 | 3.67736332148291e-05 | 0.227152 | 0.019805 | 1.07e-29 | 5.61570469798658e-29 |
| 15515-2 | SL000572 | SAA | SAA1 | P0DJI8 | 0.169382 | 0.034878 | 1.40133077843202e-06 | 4.54681790073391e-05 | 0.245006 | 0.019971 | 1.29e-33 | 8.20146341463415e-33 |
| 3352-80 | SL010288 | Carbonic anhydrase 6 | CA6 | P23280 | -0.17046 | 0.035112 | 1.41233762312836e-06 | 4.56216437550973e-05 | -0.20783 | 0.020347 | 5.17e-24 | 2.01141293532338e-23 |
| 25245-22 | SL023554 | RIC8A | RIC8A | Q9NPQ8 | 0.180933 | 0.037285 | 1.42673573403636e-06 | 4.58828111282137e-05 | 0.193869 | 0.018125 | 3.99e-26 | 1.78296e-25 |
| 2571-12 | SL000045 | IGFBP-3 | IGFBP3 | P17936 | -0.15548 | 0.03207 | 1.45938106787231e-06 | 4.67259101378676e-05 | -0.12244 | 0.019266 | 2.47e-10 | 3.96620123203285e-10 |
| 17735-130 | SL020978 | GBRAP | GABARAP | O95166 | 0.171436 | 0.03541 | 1.50814378767054e-06 | 4.8075390564866e-05 | 0.280787 | 0.020114 | 1.15e-42 | 1.21527027027027e-41 |
| 8337-65 | SL018463 | PTPRU | PTPRU | Q92729 | 0.173982 | 0.035961 | 1.53311056399993e-06 | 4.86578496906178e-05 | 0.25959 | 0.02089 | 2.03e-34 | 1.34530508474576e-33 |
| 13053-6 | SL015342 | S6A14 | SLC6A14 | Q9UN76 | 0.178958 | 0.037086 | 1.63053202611299e-06 | 5.15248120251704e-05 | 0.067811 | 0.017255 | 8.73e-05 | 0.000104706 |
| 5698-60 | SL005005 | Tenascin-X | TNXB | P22105 | -0.16858 | 0.035416 | 2.24202279803186e-06 | 7.05412194636172e-05 | -0.18008 | 0.020616 | 4.43e-18 | 1.15861538461538e-17 |
| 13109-82 | SL008810 | NEGR1 | NEGR1 | Q7Z3B1 | -0.16979 | 0.035683 | 2.26141412293822e-06 | 7.08446458858406e-05 | -0.18755 | 0.020147 | 2.79e-20 | 8.20218045112782e-20 |
| 3175-51 | SL006610 | ATS13 | ADAMTS13 | Q76LX8 | -0.16337 | 0.034347 | 2.28300609876104e-06 | 7.12141129862455e-05 | -0.13318 | 0.020465 | 9.22e-11 | 1.52110548523207e-10 |
| 10908-2 | SL017992 | GLT13 | GALNT13 | Q8IUC8 | -0.17695 | 0.037252 | 2.35292546970714e-06 | 7.30814628796218e-05 | -0.1501 | 0.01886 | 2.64e-15 | 5.73466666666667e-15 |
| 8620-56 | SL003919 | kallikrein 14 | KLK14 | Q9P0G3 | -0.16876 | 0.035555 | 2.3931552382797e-06 | 7.37236976055379e-05 | -0.19413 | 0.020214 | 1.84e-21 | 5.87297959183673e-21 |
| 10832-24 | SL018597 | B4GT6 | B4GALT6 | Q9UBX8 | -0.17159 | 0.036153 | 2.39829447356152e-06 | 7.37236976055379e-05 | -0.19576 | 0.01989 | 1.93e-22 | 6.76798206278027e-22 |
| 18893-26 | SL009016 | GPR56 | ADGRG1 | Q9Y653 | 0.166886 | 0.035166 | 2.40403361757189e-06 | 7.37236976055379e-05 | 0.037715 | 0.020047 | 0.060044 | 0.064675493 |
| 9805-51 | SL009047 | SEM4B | SEMA4B | Q9NPR2 | -0.17354 | 0.036604 | 2.4582924934868e-06 | 7.48362496412301e-05 | -0.15829 | 0.018994 | 1.29e-16 | 3.06620060790274e-16 |
| 3035-80 | SL004354 | IL-19 | IL19 | Q9UHD0 | -0.16985 | 0.035828 | 2.46090584263263e-06 | 7.48362496412301e-05 | -0.09071 | 0.021985 | 3.81e-05 | 4.67726844583987e-05 |
| 10980-11 | SL007155 | ACES | ACHE | P22303 | -0.1669 | 0.035223 | 2.49002975520108e-06 | 7.54064010866729e-05 | -0.1855 | 0.020081 | 5.32e-20 | 1.52389743589744e-19 |
| 5939-42 | SL004365 | TWEAK | TNFSF12 | O43508 | -0.17129 | 0.036347 | 2.81786286304187e-06 | 8.49801962182088e-05 | -0.15492 | 0.016784 | 5.69e-20 | 1.62393430656934e-19 |
| 2991-9 | SL001997 | IL-1 sRI | IL1R1 | P14778 | -0.16577 | 0.035261 | 2.97765561802363e-06 | 8.94281034371726e-05 | -0.20972 | 0.021012 | 5.09e-23 | 1.87753773584906e-22 |
| 23020-18 | SL005549 | RGS4 | RGS4 | P49798 | 0.168623 | 0.035889 | 3.0154805020798e-06 | 9.0191408597185e-05 | 0.251896 | 0.020109 | 6.22e-35 | 4.38201801801802e-34 |
| 5701-81 | SL001871 | Tetranectin | CLEC3B | P05452 | -0.16272 | 0.03464 | 3.03023363757926e-06 | 9.02612216308445e-05 | -0.13077 | 0.021311 | 9.85e-10 | 1.54672690763052e-09 |
| 8039-41 | SL018368 | F177A | FAM177A1 | Q8N128 | -0.16287 | 0.034896 | 3.49607213503062e-06 | 0.000103712 | -0.17178 | 0.020332 | 4.97e-17 | 1.21835109717868e-16 |
| 8866-53 | SL018693 | QPCTL | QPCTL | Q9NXS2 | -0.15809 | 0.033931 | 3.63697758391188e-06 | 0.000107453 | -0.24874 | 0.019921 | 1e-34 | 6.92035398230088e-34 |
| 3803-10 | SL008382 | CYTD | CST5 | P28325 | -0.15288 | 0.032896 | 3.84889381013714e-06 | 0.000113254 | -0.10967 | 0.020102 | 5.37e-08 | 7.77655555555556e-08 |
| 5000-52 | SL006522 | LG3BP | LGALS3BP | Q08380 | 0.173506 | 0.037368 | 3.92174117367106e-06 | 0.000114932 | 0.219151 | 0.020832 | 2.43e-25 | 1.03839344262295e-24 |
| 7215-18 | SL003669 | NADH-cytochrome b5 reductase | CYB5R3 | P00387 | -0.16759 | 0.036212 | 4.2141804714603e-06 | 0.000123007 | -0.15396 | 0.020109 | 2.74e-14 | 5.59446475195822e-14 |
| 9256-78 | SL012506 | NPTX1 | NPTX1 | Q15818 | -0.16767 | 0.036302 | 4.40091239372125e-06 | 0.000127943 | -0.13752 | 0.018699 | 2.61e-13 | 5.10255e-13 |
| 18411-83 | SL021354 | ARL15 | ARL15 | Q9NXU5 | 0.170617 | 0.036963 | 4.46169038735533e-06 | 0.000128722 | 0.292443 | 0.020477 | 1.69e-44 | 2.0332e-43 |
| 12817-1 | SL019954 | GEM | GEM | P55040 | 0.169515 | 0.036725 | 4.46310407925159e-06 | 0.000128722 | 0.119782 | 0.020884 | 1.09e-08 | 1.63919230769231e-08 |
| 9599-6 | SL019019 | PIANP | PIANP | Q8IYJ0 | -0.15732 | 0.034129 | 4.59951985714085e-06 | 0.000132132 | -0.17452 | 0.017278 | 1.58e-23 | 5.96888888888889e-23 |
| 5111-15 | SL008728 | NRX3B | NRXN3 | Q9HDB5 | -0.16055 | 0.034903 | 4.81021132201855e-06 | 0.000137403 | -0.21429 | 0.019975 | 2.87e-26 | 1.31247953216374e-25 |
| 12759-47 | SL004792 | DRBP76 | ILF3 | Q12906 | 0.157869 | 0.034324 | 4.82082800842189e-06 | 0.000137403 | 0.156352 | 0.018193 | 1.47e-17 | 3.73227272727273e-17 |
| 5092-51 | SL025896 | JAG1:ECD | JAG1 | P78504 | -0.16474 | 0.035835 | 4.8692148613185e-06 | 0.00013824 | -0.13862 | 0.02074 | 2.87e-11 | 4.9110284463895e-11 |
| 13666-222 | SL010449 | Carbonic Anhydrase X | CA10 | Q9NS85 | -0.16508 | 0.036008 | 5.16715150151878e-06 | 0.000145672 | -0.19544 | 0.020339 | 1.74e-21 | 5.57655737704918e-21 |
| 4125-52 | SL003680 | sRAGE | AGER | Q15109 | -0.15408 | 0.033609 | 5.17108520963653e-06 | 0.000145672 | -0.11962 | 0.018746 | 2.1e-10 | 3.4e-10 |
| 14072-9 | SL005679 | TCTP | TPT1 | P13693 | 0.170827 | 0.0373 | 5.28573283685189e-06 | 0.000147916 | 0.205885 | 0.020907 | 1.83e-22 | 6.44621621621622e-22 |
| 15544-25 | SL003919 | kallikrein 14 | KLK14 | Q9P0G3 | -0.16223 | 0.035425 | 5.29144509935893e-06 | 0.000147916 | -0.19386 | 0.020014 | 8.48e-22 | 2.79804219409283e-21 |
| 5644-60 | SL007198 | RNAS4 | RNASE4 | P34096 | 0.152288 | 0.033392 | 5.78109863344364e-06 | 0.000160985 | 0.194567 | 0.02086 | 2.36e-20 | 7.01718631178707e-20 |
| 8773-172 | SL025844 | EMIL3:region 1 | EMILIN3 | Q9NT22 | -0.16487 | 0.036159 | 5.810736344877e-06 | 0.000161192 | -0.10064 | 0.019256 | 1.88e-07 | 2.6441726618705e-07 |
| 8368-102 | SL001800 | TNF sR-II | TNFRSF1B | P20333 | 0.142506 | 0.031264 | 5.84815229029879e-06 | 0.000161614 | 0.163834 | 0.017433 | 1.24e-20 | 3.81763779527559e-20 |
| 7008-13 | SL025852 | F176C:CD | EVA1C | P58658 | -0.16524 | 0.036316 | 6.06960301138196e-06 | 0.000167098 | -0.15093 | 0.021202 | 1.43e-12 | 2.66887828162291e-12 |
| 4910-21 | SL004064 | GIB | PLA2G1B | P04054 | -0.16953 | 0.037298 | 6.20660815106159e-06 | 0.000170225 | -0.07421 | 0.020087 | 0.000225 | 0.00026522 |
| 15468-14 | SL002690 | FHR1 | CFHR1 | Q03591 | 0.155849 | 0.034331 | 6.37192015116331e-06 | 0.000174102 | 0.185257 | 0.019915 | 2.95e-20 | 8.60783582089552e-20 |
| 19555-1 | SL013060 | SUMO2 | SUMO2 | P61956 | 0.160066 | 0.03537 | 6.80348782944125e-06 | 0.000185198 | 0.167378 | 0.019569 | 2.08e-17 | 5.23009646302251e-17 |
| 9278-9 | SL004712 | SDF-1 | CXCL12 | P48061 | -0.16485 | 0.036468 | 6.96415164203818e-06 | 0.000188864 | -0.16592 | 0.021239 | 8.29e-15 | 1.73801072386059e-14 |
| 6414-8 | SL008961 | OAF | OAF | Q86UD1 | 0.161039 | 0.035638 | 7.0172004302608e-06 | 0.000189595 | 0.127595 | 0.019281 | 4.47e-11 | 7.48509635974304e-11 |
| 6590-54 | SL005203 | Neuropilin-2 | NRP2 | O60462 | -0.16379 | 0.036295 | 7.2120681583659e-06 | 0.000194138 | -0.14355 | 0.021295 | 1.96e-11 | 3.38348785871965e-11 |
| 12370-30 | SL006348 | Apo F | APOF | Q13790 | -0.14285 | 0.031688 | 7.37749574335665e-06 | 0.000197534 | -0.20537 | 0.020478 | 3.17e-23 | 1.18044761904762e-22 |
| 3362-61 | SL009400 | CRDL1 | CHRDL1 | Q9BU40 | -0.1166 | 0.025868 | 7.39256311399622e-06 | 0.000197534 | -0.15912 | 0.020146 | 4.24e-15 | 9.03455040871935e-15 |
| 17396-23 | SL008013 | ADH1A | ADH1A | P07327 | 0.161337 | 0.0358 | 7.42099772920754e-06 | 0.000197567 | 0.215454 | 0.020916 | 2.19e-24 | 8.82773195876289e-24 |
| 20173-39 | SL021143 | CAN9 | CAPN9 | O14815 | 0.164414 | 0.036533 | 7.63725680875121e-06 | 0.000202582 | 0.077823 | 0.01878 | 3.53e-05 | 4.35403785488959e-05 |
| 7190-50 | SL013797 | EPHA4 | EPHA4 | P54764 | -0.16419 | 0.036497 | 7.69186376198713e-06 | 0.000203289 | -0.21241 | 0.020681 | 2.96e-24 | 1.1749847715736e-23 |
| 16927-9 | SL003003 | Coagulation factor XIII | F13A1\|F13B | P00488\|P05160 | 0.164348 | 0.03656 | 7.8212180342459e-06 | 0.000205959 | 0.171195 | 0.021318 | 1.5e-15 | 3.30422535211268e-15 |
| 6423-66 | SL017430 | C1QL2 | C1QL2 | Q7Z5L3 | -0.1611 | 0.035851 | 7.88008761349547e-06 | 0.00020676 | -0.14309 | 0.021401 | 2.83e-11 | 4.85320175438596e-11 |
| 2620-4 | SL003872 | gp130, soluble | IL6ST | P40189 | -0.16287 | 0.036305 | 8.15190561181217e-06 | 0.000213122 | -0.10161 | 0.020959 | 1.32e-06 | 1.76451282051282e-06 |
| 19241-31 | SL005353 | RBP-III | RBP5 | P82980 | 0.15631 | 0.034867 | 8.2713904080821e-06 | 0.000215471 | 0.22875 | 0.0197 | 2.24e-30 | 1.23357746478873e-29 |
| 2643-57 | SL004183 | P-Cadherin | CDH3 | P22223 | 0.155214 | 0.034678 | 8.54849321078098e-06 | 0.000221894 | 0.083286 | 0.021096 | 8.1e-05 | 9.75993836671803e-05 |
| 12934-1 | SL015312 | HERC5 | HERC5 | Q9UII4 | 0.164483 | 0.036758 | 8.59652178003952e-06 | 0.000222347 | 0.147488 | 0.017254 | 2.17e-17 | 5.43891025641026e-17 |
| 6315-58 | SL017411 | PLBL1 | PLBD1 | Q6P4A8 | 0.163156 | 0.036469 | 8.63286373207708e-06 | 0.000222495 | 0.146405 | 0.016825 | 5.91e-18 | 1.52786798679868e-17 |
| 10743-13 | SL008760 | SLIK1 | SLITRK1 | Q96PX8 | -0.14357 | 0.032132 | 8.86277791470865e-06 | 0.000227614 | -0.05653 | 0.019664 | 0.004076 | 0.004553139 |
| 7139-14 | SL018085 | SLIK4 | SLITRK4 | Q8IW52 | -0.12612 | 0.02832 | 9.47640395623909e-06 | 0.000242516 | -0.11997 | 0.018087 | 4.04e-11 | 6.80879310344828e-11 |
| 2837-3 | SL000134 | Met | MET | P08581 | -0.16226 | 0.036502 | 9.83358770747808e-06 | 0.000250774 | -0.18415 | 0.021132 | 5.31e-18 | 1.3795415282392e-17 |
| 20584-4 | SL025941 | nectin-1 gamma:ECD | NECTIN1 | Q15223 | -0.16159 | 0.036357 | 9.87029773607684e-06 | 0.00025083 | -0.09766 | 0.020566 | 2.17e-06 | 2.8664527027027e-06 |
| 8908-14 | SL025907 | KCE1L:CD | KCNE5 | Q9UJ90 | -0.16194 | 0.036551 | 1.0517184173579e-05 | 0.000266338 | -0.2704 | 0.019513 | 4.31e-42 | 4.32105128205128e-41 |
| 18220-141 | SL013277 | SRA1 | SRA1 | Q9HD15 | 0.16102 | 0.036382 | 1.07428084584368e-05 | 0.000270979 | 0.091676 | 0.021902 | 2.94e-05 | 3.64933333333333e-05 |
| 18339-207 | SL008674 | PSB3 | PSMB3 | P49720 | 0.163072 | 0.036851 | 1.07750469691833e-05 | 0.000270979 | 0.143417 | 0.018137 | 3.95e-15 | 8.4627397260274e-15 |
| 17828-3 | SL005106 | S100A14 | S100A14 | Q9HCY8 | -0.16536 | 0.037376 | 1.08206556700964e-05 | 0.000271188 | -0.08864 | 0.019674 | 6.94e-06 | 8.92611842105263e-06 |
| 10916-44 | SL013523 | PLA2R | PLA2R1 | Q13018 | -0.15886 | 0.035936 | 1.10041267760355e-05 | 0.000274838 | -0.13857 | 0.021706 | 2.06e-10 | 3.34215767634855e-10 |
| 2828-82 | SL004645 | HAI-1 | SPINT1 | O43278 | -0.15833 | 0.035842 | 1.11612070544147e-05 | 0.000277807 | -0.12824 | 0.02117 | 1.59e-09 | 2.47685258964143e-09 |
| 15513-108 | SL003260 | Prostasin | PRSS8 | Q16651 | 0.154659 | 0.035117 | 1.18490268383903e-05 | 0.000293921 | 0.081915 | 0.018931 | 1.57e-05 | 1.98022580645161e-05 |
| 4842-62 | SL000070 | Glypican 3 | GPC3 | P51654 | -0.15839 | 0.03603 | 1.22969311818373e-05 | 0.000303994 | -0.20592 | 0.018618 | 8.83e-28 | 4.34280503144654e-27 |
| 4908-6 | SL004482 | Endoglin | ENG | P17813 | -0.16083 | 0.036647 | 1.27103984407814e-05 | 0.00031315 | -0.10533 | 0.020912 | 5.07e-07 | 6.94350262697023e-07 |
| 5542-22 | SL006397 | NRP1 | NRP1 | O14786 | -0.15535 | 0.03542 | 1.28656843994411e-05 | 0.000315905 | -0.08546 | 0.02117 | 5.59e-05 | 6.77733333333333e-05 |
| 11232-46 | SL003164 | Holo-TC I | TCN1 | P20061 | -0.15876 | 0.036318 | 1.37266062150181e-05 | 0.000335909 | -0.1249 | 0.020916 | 2.7e-09 | 4.16449704142012e-09 |
| 5076-53 | SL014294 | EPHAA | EPHA10 | Q5JZY3 | -0.1629 | 0.037295 | 1.39565780728793e-05 | 0.000340391 | -0.13469 | 0.018592 | 5.78e-13 | 1.10242926829268e-12 |
| 25285-14 | SL023668 | KBTBB | KBTBD11 | O94819 | -0.16273 | 0.037269 | 1.40513837412747e-05 | 0.000341557 | -0.12496 | 0.019922 | 4.19e-10 | 6.68689795918367e-10 |
| 15503-20 | SL004647 | Lefty-A | LEFTY2 | O00292 | -0.15773 | 0.036295 | 1.54037975647639e-05 | 0.000372499 | -0.14392 | 0.021037 | 9.87e-12 | 1.74228893905192e-11 |
| 12960-9 | SL007208 | HXK4 | GCK | P35557 | 0.160915 | 0.037031 | 1.54268372325938e-05 | 0.000372499 | 0.124368 | 0.020584 | 1.76e-09 | 2.7362226640159e-09 |
| 8080-24 | SL011622 | PSMP | MSMP | Q1L6U9 | -0.15395 | 0.035563 | 1.66026389656601e-05 | 0.000399563 | -0.09618 | 0.021514 | 8.17e-06 | 1.04736721311475e-05 |
| 4145-58 | SL004359 | Neurotrophin-3 | NTF3 | P20783 | -0.16058 | 0.037126 | 1.68779538315736e-05 | 0.000404848 | -0.12398 | 0.019256 | 1.45e-10 | 2.36722338204593e-10 |
| 7173-141 | SL025999 | T132C:ECD | TMEM132C | Q8N3T6 | -0.15631 | 0.036152 | 1.6988851366913e-05 | 0.000406168 | -0.15207 | 0.019315 | 5.16e-15 | 1.09352845528455e-14 |
| 18942-11 | SL016413 | PSB9 | PSMB9 | P28065 | -0.15896 | 0.036924 | 1.84657521351287e-05 | 0.00044003 | -0.17776 | 0.02001 | 1.23e-18 | 3.30536082474227e-18 |
| 21397-21 | SL022545 | PRTG | PRTG | Q2VWP7 | -0.15834 | 0.036846 | 1.90985347414524e-05 | 0.000453621 | -0.17539 | 0.020854 | 6.87e-17 | 1.67885625e-16 |
| 6042-52 | SL003198 | Tenascin | TNC | P24821 | -0.15773 | 0.036731 | 1.9373531438885e-05 | 0.000458654 | -0.15712 | 0.021422 | 3.02e-13 | 5.86014888337469e-13 |
| 16785-45 | SL004530 | HD-5 | DEFA5 | Q01523 | -0.15368 | 0.035851 | 2.00324917004216e-05 | 0.000472715 | -0.13487 | 0.021203 | 2.39e-10 | 3.85356701030928e-10 |
| 2982-82 | SL005165 | Galectin-4 | LGALS4 | P56470 | 0.159189 | 0.03738 | 2.2642506501363e-05 | 0.00053094 | 0.134954 | 0.019788 | 1.14e-11 | 1.99883408071749e-11 |
| 9294-45 | SL012443 | MFAP2 | MFAP2 | P55001 | 0.143459 | 0.033686 | 2.26460381551369e-05 | 0.00053094 | 0.139472 | 0.020226 | 6.8e-12 | 1.21963302752294e-11 |
| 13678-169 | SL003327 | Factor D | CFD | P00746 | 0.147169 | 0.034564 | 2.27268664556629e-05 | 0.000531122 | 0.072231 | 0.020882 | 0.000552 | 0.000636096 |
| 16288-17 | SL013797 | EPHA4 | EPHA4 | P54764 | -0.15717 | 0.036961 | 2.32753804180949e-05 | 0.000542197 | -0.18232 | 0.020953 | 5.92e-18 | 1.52786798679868e-17 |
| 7923-41 | SL018333 | SEM4C | SEMA4C | Q9C0C4 | -0.15797 | 0.037178 | 2.36222113202015e-05 | 0.000548518 | -0.04914 | 0.018468 | 0.007844 | 0.008676439 |
| 8052-115 | SL018410 | NLGN1 | NLGN1 | Q8N2Q7 | -0.15343 | 0.03625 | 2.54024044524242e-05 | 0.000587977 | -0.10525 | 0.020224 | 2.11e-07 | 2.95702508960573e-07 |
| 23640-10 | SL008316 | EHD2 | EHD2 | Q9NZN4 | 0.145345 | 0.034414 | 2.64225306790067e-05 | 0.000609647 | 0.185064 | 0.020111 | 7.31e-20 | 2.07116666666667e-19 |
| 6252-62 | SL006092 | Secretoglobin family 3A member 1 | SCGB3A1 | Q96QR1 | -0.14992 | 0.035514 | 2.66421172001192e-05 | 0.000612454 | -0.24066 | 0.020464 | 4.26e-31 | 2.46764444444444e-30 |
| 12758-47 | SL019871 | GRID2 | GRID2 | O43424 | -0.15598 | 0.036955 | 2.67127232669831e-05 | 0.000612454 | -0.16781 | 0.020534 | 4.81e-16 | 1.10305571847507e-15 |
| 13614-6 | SL003777 | CREB-binding protein | CREBBP | Q92793 | 0.155571 | 0.036868 | 2.68580646255509e-05 | 0.00061385 | 0.181281 | 0.020994 | 1.04e-17 | 2.64912052117264e-17 |
| 18196-8 | SL004742 | Afamin | AFM | P43652 | 0.153693 | 0.036445 | 2.71496885853039e-05 | 0.00061857 | 0.104574 | 0.021201 | 8.67e-07 | 1.17097409326425e-06 |
| 9357-4 | SL008587 | CREG1 | CREG1 | O75629 | 0.155237 | 0.036824 | 2.73294544043516e-05 | 0.00062072 | 0.084888 | 0.019867 | 2e-05 | 2.50641025641026e-05 |
| 7787-25 | SL018231 | LIRA5 | LILRA5 | A6NI73 | 0.147594 | 0.035071 | 2.81916011363926e-05 | 0.000638307 | 0.221546 | 0.019994 | 7.13e-28 | 3.55137579617834e-27 |
| 4158-54 | SL000613 | uPA | PLAU | P00749 | -0.1548 | 0.036837 | 2.89460375303233e-05 | 0.000653353 | -0.08297 | 0.020347 | 4.7e-05 | 5.71601866251944e-05 |
| 2889-37 | SL002783 | Cardiotrophin-1 | CTF1 | Q16619 | 0.15608 | 0.03719 | 2.96580472214928e-05 | 0.000667352 | 0.005594 | 0.018328 | 0.76024 | 0.767106886 |
| 2942-50 | SL000396 | Cytochrome c | CYCS | P99999 | -0.14917 | 0.035594 | 3.04328201582758e-05 | 0.000682672 | -0.11623 | 0.017313 | 2.36e-11 | 4.05608791208791e-11 |
| 2748-3 | SL001938 | Activin A | INHBA | P08476 | 0.142446 | 0.034011 | 3.07691373812075e-05 | 0.000687571 | 0.193786 | 0.02082 | 2.82e-20 | 8.25932584269663e-20 |
| 11081-1 | SL007151 | GPDA | GPD1 | P21695 | 0.154984 | 0.037009 | 3.08404044314907e-05 | 0.000687571 | 0.171714 | 0.020537 | 1.03e-16 | 2.47073619631902e-16 |
| 5108-72 | SL005209 | Notch-3 | NOTCH3 | Q9UM47 | -0.14726 | 0.035232 | 3.19192231762879e-05 | 0.000709446 | -0.1764 | 0.020842 | 4.4e-17 | 1.08542586750789e-16 |
| 2475-1 | SL004010 | SCF sR | KIT | P10721 | -0.14082 | 0.033768 | 3.32448586943017e-05 | 0.00073605 | -0.19011 | 0.021021 | 3e-19 | 8.28975265017668e-19 |
| 11424-4 | SL008049 | FAAA | FAH | P16930 | 0.15102 | 0.036217 | 3.33187057591461e-05 | 0.00073605 | 0.167611 | 0.020777 | 1.12e-15 | 2.49527065527066e-15 |
| 25960-15 | SL006111 | T-plastin | PLS3 | P13797 | 0.15239 | 0.036595 | 3.41331023101187e-05 | 0.000751756 | 0.187098 | 0.021127 | 1.56e-18 | 4.14938775510204e-18 |
| 9484-75 | SL004857 | Desmoglein-2 | DSG2 | Q14126 | -0.14971 | 0.035965 | 3.43434705945282e-05 | 0.000754104 | -0.16369 | 0.019858 | 2.7e-16 | 6.26528189910979e-16 |
| 4246-40 | SL004154 | NCAM-L1 | L1CAM | P32004 | -0.15332 | 0.036878 | 3.51627709685971e-05 | 0.000769768 | -0.15205 | 0.021653 | 2.82e-12 | 5.1888e-12 |
| 13738-8 | SL001932 | Inhibin bA chain | INHBA | P08476 | 0.149901 | 0.036085 | 3.56527264963006e-05 | 0.000775964 | 0.252811 | 0.021015 | 2.01e-32 | 1.227984375e-31 |
| 15476-6 | SL007493 | REG3G | REG3G | Q6UW15 | -0.14927 | 0.035934 | 3.56593421164014e-05 | 0.000775964 | -0.02727 | 0.021278 | 0.200033 | 0.210249319 |
| 14133-93 | SL000145 | IL-1 sRII | IL1R2 | P27930 | -0.14755 | 0.035553 | 3.62798064660938e-05 | 0.000787109 | -0.03798 | 0.02147 | 0.077047 | 0.082310094 |
| 13665-35 | SL004990 | PP2A, subunit B | PPP2R3A | Q06190 | 0.152448 | 0.036825 | 3.79116145520203e-05 | 0.000818904 | 0.14959 | 0.01975 | 5.1e-14 | 1.0252442159383e-13 |
| 3516-60 | SL004712 | SDF-1 | CXCL12 | P48061 | -0.14796 | 0.035744 | 3.79706596465371e-05 | 0.000818904 | -0.19317 | 0.020845 | 4.09e-20 | 1.18021402214022e-19 |
| 23259-23 | SL023427 | EFMT1 | EEF1AKMT1 | Q8WVE0 | 0.153095 | 0.037025 | 3.87077612822831e-05 | 0.000832331 | 0.142937 | 0.021416 | 3.06e-11 | 5.21333333333333e-11 |
| 19584-33 | SL003849 | FGF9 | FGF9 | P31371 | -0.15334 | 0.037094 | 3.88840482894127e-05 | 0.000833656 | -0.16221 | 0.020505 | 3.84e-15 | 8.24967032967033e-15 |
| 18930-28 | SL008427 | SLIT2 | SLIT2 | O94813 | -0.13627 | 0.032991 | 3.94625591426001e-05 | 0.00084357 | -0.20317 | 0.018979 | 3.64e-26 | 1.65493023255814e-25 |
| 11547-84 | SL019518 | MUSK | MUSK | O15146 | 0.154525 | 0.037457 | 4.031099167378e-05 | 0.00085918 | 0.091033 | 0.02012 | 6.34e-06 | 8.16784184514003e-06 |
| 11219-95 | SL012727 | FGFP3 | FGFBP3 | Q8TAT2 | -0.14419 | 0.034969 | 4.06644528980942e-05 | 0.000862109 | -0.0907 | 0.021137 | 1.85e-05 | 2.32962962962963e-05 |
| 6260-14 | SL003198 | Tenascin | TNC | P24821 | -0.15071 | 0.036552 | 4.06856588509582e-05 | 0.000862109 | -0.10889 | 0.021547 | 4.65e-07 | 6.37947368421053e-07 |
| 8397-147 | SL018482 | QSOX2 | QSOX2 | Q6ZRP7 | -0.14393 | 0.034963 | 4.18722838000851e-05 | 0.000884674 | -0.09897 | 0.021109 | 2.9e-06 | 3.79230769230769e-06 |
| 16763-11 | SL017261 | LECT2 | LECT2 | O14960 | 0.147652 | 0.035896 | 4.243753927239e-05 | 0.000894017 | 0.231574 | 0.019783 | 7.86e-31 | 4.454e-30 |
| 14294-61 | SL013880 | MBD1 | MBD1 | Q9UIS9 | 0.153577 | 0.037372 | 4.31650678462329e-05 | 0.000906716 | 0.194373 | 0.021377 | 1.95e-19 | 5.44607142857143e-19 |
| 5738-25 | SL003198 | Tenascin | TNC | P24821 | -0.15037 | 0.036598 | 4.33061657074749e-05 | 0.000907058 | -0.1061 | 0.021692 | 1.07e-06 | 1.43769759450172e-06 |
| 7198-197 | SL012378 | FA20B | FAM20B | O75063 | -0.14385 | 0.03507 | 4.46023277012086e-05 | 0.000931522 | -0.19637 | 0.019049 | 2.02e-24 | 8.18466321243523e-24 |
| 23359-25 | SL023708 | FL2D | WTAP | Q15007 | -0.15241 | 0.037177 | 4.49893417151249e-05 | 0.000936913 | -0.09301 | 0.020906 | 9.01e-06 | 1.15127777777778e-05 |
| 11709-29 | SL015112 | CPT1B | CPT1B | Q92523 | -0.15182 | 0.037101 | 4.64279140863802e-05 | 0.000964109 | -0.17272 | 0.019493 | 1.51e-18 | 4.0301023890785e-18 |
| 24957-6 | SL014551 | ESPN | None | B1AK53 | 0.147382 | 0.036086 | 4.80298810648249e-05 | 0.000994533 | 0.26197 | 0.020353 | 9.93e-37 | 7.76526e-36 |
| 3520-58 | SL000089 | TGF-b3 | TGFB3 | P10600 | 0.151653 | 0.037142 | 4.82637583391637e-05 | 0.000996537 | 0.089419 | 0.018515 | 1.45e-06 | 1.93498293515358e-06 |
| 21104-37 | SL022285 | UBL5 | UBL5 | Q9BZL1 | 0.152155 | 0.037295 | 4.89321402547267e-05 | 0.001007475 | 0.206081 | 0.021384 | 1.35e-21 | 4.34444444444444e-21 |
| 20516-11 | SL015287 | SCN3B | SCN3B | Q9NY72 | -0.1509 | 0.037007 | 4.93518201722512e-05 | 0.001013246 | -0.03965 | 0.019579 | 0.042977 | 0.046613057 |
| 20442-12 | SL020201 | RCAN3 | RCAN3 | Q9UKA8 | 0.150411 | 0.036967 | 5.12632913169919e-05 | 0.001049526 | 0.213312 | 0.021079 | 1.31e-23 | 4.97291262135922e-23 |
| 24926-9 | SL013144 | FUBP2 | KHSRP | Q92945 | 0.146213 | 0.035974 | 5.22029229364508e-05 | 0.001065761 | 0.194246 | 0.019111 | 8.47e-24 | 3.24683333333333e-23 |
| 9511-61 | SL012390 | NXPH2 | NXPH2 | O95156 | -0.14881 | 0.036807 | 5.71174554459165e-05 | 0.001162828 | -0.25673 | 0.020188 | 6.41e-36 | 4.81982692307692e-35 |
| 4917-62 | SL003182 | Integrin aVb5 | ITGAV\|ITGB5 | P06756\|P18084 | -0.14679 | 0.036326 | 5.76559782229626e-05 | 0.001170513 | -0.0687 | 0.019337 | 0.000389 | 0.000453001 |
| 6561-77 | SL017496 | Ig K chain V-I region HK102-like | IGKV1-5 | P01602 | 0.146222 | 0.036197 | 5.79307500262274e-05 | 0.001172815 | 0.06417 | 0.020427 | 0.001702 | 0.001925915 |
| 12988-49 | SL007342 | EWS | EWSR1 | Q01844 | 0.138175 | 0.034341 | 6.194675561865e-05 | 0.001250636 | 0.163411 | 0.019675 | 1.62e-16 | 3.81578313253012e-16 |
| 4480-59 | SL000314 | C3b | C3 | P01024 | 0.146719 | 0.03649 | 6.26882008742165e-05 | 0.001262099 | 0.293588 | 0.020029 | 1.1e-46 | 1.50912280701754e-45 |
| 5109-24 | SL005210 | Nr-CAM | NRCAM | Q92823 | -0.1488 | 0.037087 | 6.4937045708422e-05 | 0.001303764 | -0.14995 | 0.021221 | 2.07e-12 | 3.83587677725118e-12 |
| 3336-50 | SL001998 | TFPI | TFPI | P10646 | 0.139035 | 0.034669 | 6.54680334471006e-05 | 0.001310803 | 0.207697 | 0.020815 | 5.2e-23 | 1.90910798122066e-22 |
| 12661-44 | SL019788 | GBRL1 | GABARAPL1 | Q9H0R8 | 0.143005 | 0.035728 | 6.76195810476406e-05 | 0.001350162 | 0.268155 | 0.018992 | 1.44e-43 | 1.58602816901408e-42 |
| 8447-11 | SL004269 | ghrelin | GHRL | Q9UBU3 | -0.14549 | 0.036425 | 6.99651192597042e-05 | 0.001392216 | -0.14824 | 0.018132 | 4.68e-16 | 1.0764e-15 |
| 6997-32 | SL017971 | RAB26 | RAB26 | Q9ULW5 | -0.14819 | 0.037104 | 7.0108853086361e-05 | 0.001392216 | -0.13454 | 0.020652 | 8.82e-11 | 1.45819027484144e-10 |
| 15527-90 | SL008018 | DLDH | DLD | P09622 | -0.14833 | 0.037179 | 7.13340377868864e-05 | 0.001412686 | -0.19463 | 0.019728 | 1.54e-22 | 5.44923076923077e-22 |
| 3396-54 | SL000565 | Renin | REN | P00797 | 0.134146 | 0.03364 | 7.19412480247461e-05 | 0.00142084 | 0.064494 | 0.02074 | 0.001894 | 0.002140782 |
| 5699-19 | SL014977 | CI061 | FAM189A2 | Q15884 | -0.14626 | 0.036692 | 7.2396322403401e-05 | 0.001425952 | -0.14489 | 0.020824 | 4.43e-12 | 8.03772621809745e-12 |
| 22587-37 | SL022633 | EPHA6 | EPHA6 | Q9UF33 | -0.1218 | 0.030579 | 7.3294860900143e-05 | 0.001439749 | -0.13805 | 0.01898 | 4.71e-13 | 9.04968058968059e-13 |
| 6520-87 | SL007091 | MGP | MGP | P08493 | 0.146131 | 0.036695 | 7.35655548353337e-05 | 0.001441171 | 0.138287 | 0.020217 | 9.97e-12 | 1.75597747747748e-11 |
| 15569-15 | SL002527 | Collagen II | COL2A1 | P02458 | -0.14494 | 0.036535 | 7.83268430468829e-05 | 0.001530321 | -0.09401 | 0.021495 | 1.27e-05 | 1.60962722852512e-05 |
| 3647-49 | SL018625 | TLR4:MD-2 complex | TLR4\|LY96 | O00206\|Q9Y6Y9 | -0.14895 | 0.037565 | 7.90007608299175e-05 | 0.00153935 | -0.08194 | 0.021125 | 0.000108 | 0.000128702 |
| 22858-3 | SL007462 | MK06 | MAPK6 | Q16659 | -0.14696 | 0.037103 | 8.03692741514363e-05 | 0.001561829 | -0.15396 | 0.021648 | 1.5e-12 | 2.79285714285714e-12 |
| 9526-3 | SL019000 | COLL1 | CLPSL1 | A2RUU4 | 0.145584 | 0.036763 | 8.06190043105738e-05 | 0.001562504 | 0.204581 | 0.020562 | 6.87e-23 | 2.51043925233645e-22 |
| 8956-96 | SL025991 | SREC-II:ECD | SCARF2 | Q96GP6 | -0.11435 | 0.028886 | 8.11187087442168e-05 | 0.001568007 | -0.12462 | 0.018753 | 3.72e-11 | 6.31028199566161e-11 |
| 19361-78 | SL006992 | MATN3 | MATN3 | O15232 | -0.13087 | 0.033079 | 8.19256104041191e-05 | 0.001578463 | -0.13228 | 0.020706 | 2e-10 | 3.25155925155925e-10 |
| 3214-3 | SL006397 | NRP1 | NRP1 | O14786 | -0.14295 | 0.036137 | 8.20939776636221e-05 | 0.001578463 | -0.08012 | 0.021435 | 0.00019 | 0.000224426 |
| 4763-31 | SL004742 | Afamin | AFM | P43652 | 0.144083 | 0.036433 | 8.24024419017969e-05 | 0.001580214 | 0.109682 | 0.021213 | 2.52e-07 | 3.5064768683274e-07 |
| 2952-75 | SL000047 | IGF-I | IGF1 | P05019 | -0.13161 | 0.033306 | 8.34903217070105e-05 | 0.001596862 | -0.17813 | 0.020123 | 1.62e-18 | 4.29437288135593e-18 |
| 8042-88 | SL018399 | ISK9 | SPINK9 | Q5DT21 | -0.12835 | 0.032498 | 8.41919918263363e-05 | 0.001606056 | -0.12883 | 0.019421 | 4.02e-11 | 6.7897192224622e-11 |
| 5756-66 | SL008518 | PAPP2 | PAPPA2 | Q9BXP8 | 0.147094 | 0.037254 | 8.45797507908405e-05 | 0.001609229 | 0.107946 | 0.017811 | 1.57e-09 | 2.45057884231537e-09 |
| 13495-48 | SL020147 | HCAR2 | HCAR2 | Q8TDS4 | 0.145682 | 0.036916 | 8.53267261662252e-05 | 0.001616571 | 0.128059 | 0.018488 | 5.5e-12 | 9.91013824884793e-12 |
| 19617-5 | SL005407 | LTB4DH | PTGR1 | Q14914 | 0.145525 | 0.036883 | 8.55926202196505e-05 | 0.001616571 | 0.182258 | 0.020892 | 4.88e-18 | 1.27205333333333e-17 |
| 14048-7 | SL004588 | IL-1 R AcP | IL1RAP | Q9NPH3 | -0.14171 | 0.035918 | 8.56633590559518e-05 | 0.001616571 | -0.20025 | 0.020161 | 8.12e-23 | 2.95341395348837e-22 |
| 9370-69 | SL006682 | GGH | GGH | Q92820 | 0.139283 | 0.035308 | 8.58553252010877e-05 | 0.001616571 | 0.20083 | 0.019375 | 1.15e-24 | 4.73315789473684e-24 |
| 8235-48 | SL007079 | SCG1 | CHGB | P05060 | -0.13498 | 0.03424 | 8.68061145513081e-05 | 0.00163025 | -0.18671 | 0.019204 | 6.04e-22 | 2.00801694915254e-21 |
| 5363-51 | SL010470 | Semaphorin 3E | SEMA3E | O15041 | -0.14177 | 0.036 | 8.82654806516718e-05 | 0.001653385 | -0.10253 | 0.021353 | 1.67e-06 | 2.2172156196944e-06 |
| 20545-17 | SL021686 | MFAP3 | MFAP3 | P55082 | 0.146684 | 0.037324 | 9.1215385327898e-05 | 0.00170425 | -0.00464 | 0.020865 | 0.823969 | 0.829271447 |
| 5658-64 | SL003991 | coagulation factor XIII B | F13B | P05160 | 0.145445 | 0.037018 | 9.15847098397397e-05 | 0.001706763 | 0.14573 | 0.021444 | 1.35e-11 | 2.35122494432071e-11 |
| 13113-7 | SL002688 | Osteopontin | SPP1 | P10451 | -0.14224 | 0.036227 | 9.25987764443911e-05 | 0.001721248 | -0.08774 | 0.018761 | 3.07e-06 | 4.00791318864775e-06 |
| 7182-1 | SL006149 | carboxylesterase, liver | CES1 | P23141 | 0.143509 | 0.036569 | 9.33751622848371e-05 | 0.00172664 | 0.118554 | 0.018366 | 1.3e-10 | 2.13123689727463e-10 |
| 11556-19 | SL015077 | ARFG1 | ARFGAP1 | Q8N6T3 | 0.144549 | 0.03684 | 9.35992291435917e-05 | 0.00172664 | 0.116715 | 0.020467 | 1.32e-08 | 1.96992366412214e-08 |
| 10754-113 | SL005237 | Prokineticin-2 | PROK2 | Q9HC23 | 0.144222 | 0.036757 | 9.36015649754817e-05 | 0.00172664 | 0.149363 | 0.020671 | 6.63e-13 | 1.25841262135922e-12 |
| 5457-5 | SL007471 | COLEC12 | COLEC12 | Q5KU26 | -0.1368 | 0.034895 | 9.49101415034931e-05 | 0.001746347 | -0.1793 | 0.020058 | 7.58e-19 | 2.06535191637631e-18 |
| 5735-54 | SL017110 | C1GLC | C1GALT1C1 | Q96EU7 | -0.14524 | 0.037091 | 9.66792408435193e-05 | 0.001770918 | -0.13394 | 0.019144 | 3.37e-12 | 6.17175644028103e-12 |
| 9211-19 | SL003066 | PEDF | SERPINF1 | P36955 | 0.136483 | 0.034856 | 9.67328582236528e-05 | 0.001770918 | 0.21728 | 0.019913 | 4.25e-27 | 2.03895705521472e-26 |
| 24664-3 | SL016466 | INM02 | EMC10 | Q5UCC4 | -0.14427 | 0.036917 | 9.98057249107792e-05 | 0.001822583 | -0.15465 | 0.019947 | 1.31e-14 | 2.70295514511873e-14 |
| 17138-8 | SL005253 | GST A1-1 | GSTA1 | P08263 | 0.139943 | 0.03584 | 0.000101159 | 0.001842674 | 0.175361 | 0.020918 | 8.59e-17 | 2.08614285714286e-16 |
| 25918-60 | SL014713 | HLAE | HLA-E | P13747 | 0.145425 | 0.037255 | 0.000101624 | 0.001846508 | 0.081786 | 0.021013 | 0.000102 | 0.00012215 |
| 6387-61 | SL004530 | HD-5 | DEFA5 | Q01523 | -0.13552 | 0.034746 | 0.000102953 | 0.001865983 | -0.09916 | 0.021063 | 2.65e-06 | 3.48285714285714e-06 |
| 21217-20 | SL008287 | PHB2 | PHB2 | Q99623 | 0.144814 | 0.037183 | 0.000105352 | 0.001904723 | 0.091872 | 0.01739 | 1.38e-07 | 1.955e-07 |
| 18909-11 | SL019371 | EXOS8 | EXOSC8 | Q96B26 | 0.146031 | 0.037534 | 0.000107112 | 0.001931743 | -0.00117 | 0.020307 | 0.954218 | 0.95543979 |
| 10612-18 | SL008694 | PLOD3 | PLOD3 | O60568 | 0.143785 | 0.036978 | 0.000108081 | 0.001944379 | 0.173974 | 0.021345 | 5.72e-16 | 1.29653333333333e-15 |
| 6543-182 | SL012472 | PRRP | PRLH | P81277 | 0.143481 | 0.03692 | 0.000108987 | 0.001955841 | 0.147502 | 0.018557 | 2.85e-15 | 6.15662983425414e-15 |
| 3299-29 | SL010455 | Contactin-5 | CNTN5 | O94779 | -0.14379 | 0.037068 | 0.000112274 | 0.002009873 | -0.12609 | 0.01839 | 8.92e-12 | 1.58173242630385e-11 |
| 9231-23 | SL017105 | IMPA3 | BPNT2 | Q9NX62 | -0.14475 | 0.037338 | 0.000113338 | 0.00202393 | -0.16778 | 0.020816 | 1.18e-15 | 2.62147727272727e-15 |
| 12721-4 | SL019891 | TAXB1 | TAX1BP1 | Q86VP1 | 0.144477 | 0.037312 | 0.000115411 | 0.002055896 | 0.016842 | 0.016647 | 0.311783 | 0.32293327 |
| 20517-1 | SL004857 | Desmoglein-2 | DSG2 | Q14126 | -0.14242 | 0.036807 | 0.000116627 | 0.002072488 | -0.1276 | 0.02052 | 5.9e-10 | 9.35862068965517e-10 |
| 21492-19 | SL012507 | CLUL1 | CLUL1 | Q15846 | -0.14347 | 0.037105 | 0.000118011 | 0.002091961 | -0.16557 | 0.019397 | 2.41e-17 | 6.00197452229299e-17 |
| 9191-8 | SL002602 | Trefoil factor 2 | TFF2 | Q03403 | -0.1341 | 0.034713 | 0.00011973 | 0.002117261 | -0.13319 | 0.021233 | 4.18e-10 | 6.68458077709611e-10 |
| 12587-65 | SL019779 | ARL2 | ARL2 | P36404 | -0.14413 | 0.037365 | 0.000122474 | 0.002160545 | -0.1785 | 0.021566 | 2.05e-16 | 4.7997005988024e-16 |
| 13942-140 | SL020269 | SPSB1 | SPSB1 | Q96BD6 | 0.14206 | 0.036939 | 0.000128335 | 0.002256196 | 0.133912 | 0.018442 | 5.14e-13 | 9.85166666666667e-13 |
| 3457-57 | SL005084 | Periostin | POSTN | Q15063 | -0.13622 | 0.035423 | 0.000128518 | 0.002256196 | -0.19565 | 0.02131 | 8.84e-20 | 2.48664748201439e-19 |
| 8794-13 | SL008734 | DPEP1 | DPEP1 | P16444 | -0.14054 | 0.036575 | 0.000130006 | 0.002276837 | -0.12924 | 0.019525 | 4.43e-11 | 7.43403433476395e-11 |
| 2630-12 | SL004588 | IL-1 R AcP | IL1RAP | Q9NPH3 | -0.13788 | 0.036046 | 0.000139423 | 0.002435888 | -0.20719 | 0.01988 | 6.52e-25 | 2.69769312169312e-24 |
| 5350-14 | SL012881 | GPC6 | GPC6 | Q9Y625 | 0.141789 | 0.037116 | 0.000142211 | 0.002477286 | 0.11418 | 0.020678 | 3.71e-08 | 5.42284112149533e-08 |
| 21491-7 | SL004426 | VAP-1 | AOC3 | Q16853 | -0.13658 | 0.035756 | 0.000142475 | 0.002477286 | -0.14269 | 0.021373 | 3.02e-11 | 5.1564192139738e-11 |
| 8474-6 | SL006114 | ROR1 | ROR1 | Q01973 | -0.13054 | 0.034196 | 0.000143688 | 0.002492424 | -0.15023 | 0.020258 | 1.66e-13 | 3.26160804020101e-13 |
| 5084-154 | SL005174 | IL-17B R | IL17RB | Q9NRM6 | -0.13963 | 0.036607 | 0.000145576 | 0.002519166 | -0.11027 | 0.020923 | 1.48e-07 | 2.09287522603978e-07 |
| 3773-15 | SL003200 | sTie-2 | TEK | Q02763 | -0.14159 | 0.037127 | 0.000145957 | 0.002519752 | -0.07685 | 0.020923 | 0.000245 | 0.000287555 |
| 2590-69 | SL006114 | ROR1 | ROR1 | Q01973 | -0.12999 | 0.034159 | 0.000150777 | 0.002596801 | -0.16046 | 0.02013 | 2.39e-15 | 5.20607242339833e-15 |
| 5451-1 | SL003166 | ALCAM | ALCAM | Q13740 | -0.13608 | 0.035783 | 0.000152329 | 0.002617322 | -0.06351 | 0.019971 | 0.001492 | 0.001695281 |
| 6291-55 | SL011368 | Alcadein-beta | CLSTN3 | Q9BQT9 | 0.132946 | 0.034982 | 0.000153827 | 0.00263683 | 0.132572 | 0.017391 | 3.53e-14 | 7.15145077720207e-14 |
| 19787-14 | SL004873 | MER | MERTK | Q12866 | -0.1368 | 0.036062 | 0.000158159 | 0.002704702 | -0.11636 | 0.020369 | 1.25e-08 | 1.87260536398467e-08 |
| 8233-2 | SL012651 | ITIH5 | ITIH5 | Q86UX2 | -0.13915 | 0.036715 | 0.000160332 | 0.002735425 | -0.15361 | 0.021741 | 2.08e-12 | 3.84529550827423e-12 |
| 11318-20 | SL004840 | Apo A-V | APOA5 | Q6Q788 | -0.13547 | 0.035761 | 0.000161425 | 0.002747624 | -0.19063 | 0.02029 | 1.27e-20 | 3.89466666666667e-20 |
| 19622-7 | SL001938 | Activin A | INHBA | P08476 | 0.134271 | 0.035475 | 0.000163581 | 0.00277782 | 0.187119 | 0.021178 | 1.86e-18 | 4.89737373737374e-18 |
| 16292-288 | SL011908 | GIP | GIP | P09681 | 0.139769 | 0.03694 | 0.000164374 | 0.002784774 | 0.114057 | 0.019762 | 8.85e-09 | 1.33862669245648e-08 |
| 9313-27 | SL017250 | CBLN1 | CBLN1 | P23435 | -0.13361 | 0.035428 | 0.000172628 | 0.002917814 | -0.16302 | 0.0204 | 2.04e-15 | 4.48112359550562e-15 |
| 8906-60 | SL018698 | LRTM2 | LRTM2 | Q8N967 | -0.14072 | 0.0374 | 0.000178636 | 0.003012355 | -0.07583 | 0.016442 | 4.19e-06 | 5.43379767827529e-06 |
| 8325-37 | SL018474 | ADH4 | ADH4 | P08319 | 0.130001 | 0.034638 | 0.000185419 | 0.003119503 | 0.185798 | 0.020537 | 2.92e-19 | 8.09730496453901e-19 |
| 23605-3 | SL023496 | T11L1 | TCP11L1 | Q9NUJ3 | -0.13993 | 0.037317 | 0.000187982 | 0.003153444 | -0.19678 | 0.019221 | 4.08e-24 | 1.61139393939394e-23 |
| 7223-60 | SL005105 | S100A13 | S100A13 | Q99584 | -0.13688 | 0.03651 | 0.000188304 | 0.003153444 | -0.12289 | 0.021405 | 1.06e-08 | 1.59714836223507e-08 |
| 5879-51 | SL009207 | Dynactin subunit 2 | DCTN2 | Q13561 | 0.134765 | 0.036298 | 0.000217169 | 0.003628475 | 0.140409 | 0.019826 | 1.85e-12 | 3.43634204275534e-12 |
| 20423-40 | SL010770 | CIB1 | CIB1 | Q99828 | -0.13685 | 0.036894 | 0.000220059 | 0.003658797 | -0.19094 | 0.020395 | 1.71e-20 | 5.18302325581395e-20 |
| 8269-327 | SL012602 | ARSK | ARSK | Q6UWY0 | -0.13826 | 0.037277 | 0.00022043 | 0.003658797 | -0.15918 | 0.017745 | 5.75e-19 | 1.5722027972028e-18 |
| 9282-12 | SL009427 | CRIS2 | CRISP2 | P16562 | -0.12043 | 0.032472 | 0.000220494 | 0.003658797 | -0.17275 | 0.019135 | 3.47e-19 | 9.55471830985916e-19 |
| 5400-52 | SL003184 | sLeptin R | LEPR | P48357 | -0.13641 | 0.036797 | 0.000222058 | 0.003676356 | -0.09959 | 0.021191 | 2.75e-06 | 3.60822147651007e-06 |
| 5459-33 | SL010456 | CYTN | CST1 | P01037 | -0.13403 | 0.036177 | 0.00022395 | 0.003699245 | -0.09342 | 0.019186 | 1.19e-06 | 1.59345890410959e-06 |
| 3415-61 | SL004660 | BSP | IBSP | P21815 | -0.1344 | 0.03629 | 0.000225049 | 0.003708971 | -0.129 | 0.020288 | 2.43e-10 | 3.91e-10 |
| 9377-25 | SL002787 | SCF | KITLG | P21583 | -0.13593 | 0.036728 | 0.000227311 | 0.003731056 | -0.16348 | 0.020737 | 4.75e-15 | 1.009375e-14 |
| 21685-29 | SL003344 | DCC | DCC | P43146 | -0.13802 | 0.037295 | 0.000227416 | 0.003731056 | -0.15798 | 0.020771 | 4.02e-14 | 8.1231007751938e-14 |
| 4962-52 | SL012538 | ARMEL | CDNF | Q49AH0 | -0.13072 | 0.035333 | 0.000228514 | 0.003740622 | -0.16069 | 0.017702 | 2.23e-19 | 6.20590747330961e-19 |
| 6439-59 | SL017431 | CN093 | C14orf93 | Q9H972 | 0.13512 | 0.03654 | 0.000230115 | 0.003758378 | 0.171815 | 0.021337 | 1.26e-15 | 2.79127478753541e-15 |
| 4880-21 | SL007756 | GDF2 | GDF2 | Q9UK05 | -0.1336 | 0.036145 | 0.000231658 | 0.003775095 | -0.20853 | 0.019838 | 2.64e-25 | 1.122e-24 |
| 2888-49 | SL000323 | C7 | C7 | P10643 | -0.13717 | 0.037189 | 0.000238646 | 0.003878434 | -0.11478 | 0.021251 | 7.26e-08 | 1.043625e-07 |
| 7784-1 | SL017189 | Kininogen, HMW | KNG1 | P01042 | 0.135341 | 0.036697 | 0.000239067 | 0.003878434 | 0.198337 | 0.021251 | 2.23e-20 | 6.68145593869732e-20 |
| 11836-144 | SL004119 | discoidin domain receptor 1 | DDR1 | Q08345 | -0.13441 | 0.03646 | 0.000240433 | 0.003891903 | -0.12524 | 0.018325 | 1.04e-11 | 1.82759550561798e-11 |
| 18241-18 | SL020919 | HEM6 | CPOX | P36551 | -0.13078 | 0.035494 | 0.000242265 | 0.003912849 | -0.10901 | 0.019636 | 3.14e-08 | 4.61556390977444e-08 |
| 14144-3 | SL019979 | H2A3 | H2AW | Q7L7L0 | 0.134047 | 0.036409 | 0.000245014 | 0.003948478 | 0.166013 | 0.020118 | 2.52e-16 | 5.865e-16 |
| 12373-73 | SL014875 | TRA2B | TRA2B | P62995 | 0.1306 | 0.035499 | 0.000247536 | 0.00398029 | 0.214202 | 0.019934 | 2.39e-26 | 1.0994e-25 |
| 23631-1 | SL023276 | GALK2 | GALK2 | Q01415 | -0.1365 | 0.037158 | 0.000252761 | 0.004055331 | -0.10058 | 0.01913 | 1.59e-07 | 2.24436823104693e-07 |
| 20590-13 | SL004417 | NPY | NPY | P01303 | -0.13665 | 0.037207 | 0.000253604 | 0.004059898 | -0.18255 | 0.02053 | 1.14e-18 | 3.07406896551724e-18 |
| 10451-11 | SL008914 | NUCB1 | NUCB1 | Q02818 | 0.135421 | 0.036908 | 0.000257164 | 0.004107842 | 0.095082 | 0.019898 | 1.87e-06 | 2.47854237288136e-06 |
| 9391-60 | SL008370 | PCSK1 | PCSK1N | Q9UHG2 | -0.13687 | 0.037313 | 0.000258252 | 0.00411085 | -0.07542 | 0.018404 | 4.3e-05 | 5.24586583463339e-05 |
| 5744-12 | SL017113 | CA056 | MENT | Q9BUN1 | -0.13461 | 0.0367 | 0.000258484 | 0.00411085 | -0.2297 | 0.020996 | 3.16e-27 | 1.53485714285714e-26 |
| 5496-49 | SL005115 | Spondin-1 | SPON1 | Q9HCB6 | -0.1178 | 0.032176 | 0.000265336 | 0.004199466 | -0.12965 | 0.020345 | 2.21e-10 | 3.57070247933884e-10 |
| 9177-6 | SL007306 | FAM3B | FAM3B | P58499 | -0.13135 | 0.03588 | 0.000265413 | 0.004199466 | -0.07674 | 0.020646 | 0.000206 | 0.000243578 |
| 4971-1 | SL008380 | CATZ | CTSZ | Q9UBR2 | 0.125928 | 0.034401 | 0.000265789 | 0.004199466 | 0.175569 | 0.020952 | 8.85e-17 | 2.14263157894737e-16 |
| 12888-18 | SL012382 | SAP30 | SAP30 | O75446 | 0.137586 | 0.0376 | 0.000267233 | 0.004213125 | -0.01843 | 0.020675 | 0.372919 | 0.384219061 |
| 6895-1 | SL026013 | TR:CD | TFRC | P02786 | 0.130215 | 0.035624 | 0.00027127 | 0.004263841 | 0.106265 | 0.019923 | 1.05e-07 | 1.49290909090909e-07 |
| 8459-10 | SL003993 | BMP-6 | BMP6 | P22004 | -0.13743 | 0.0376 | 0.000271623 | 0.004263841 | -0.14899 | 0.021623 | 7.06e-12 | 1.26048401826484e-11 |
| 2879-9 | SL000248 | a1-Antichymotrypsin | SERPINA3 | P01011 | -0.13576 | 0.037182 | 0.000275554 | 0.004316228 | -0.1175 | 0.021556 | 5.52e-08 | 7.97900184842884e-08 |
| 19175-18 | SL015568 | MARCKSL1 | MARCKSL1 | P49006 | -0.13122 | 0.036 | 0.000282153 | 0.004410083 | -0.16018 | 0.020078 | 2.26e-15 | 4.93664804469274e-15 |
| 14178-18 | SL007463 | CDKN3 | CDKN3 | Q16667 | -0.13455 | 0.03699 | 0.000290343 | 0.004528359 | -0.08265 | 0.018937 | 1.33e-05 | 1.68294498381877e-05 |
| 11140-56 | SL025819 | CO1A1:C-term propeptide | COL1A1 | P02452 | -0.13246 | 0.036501 | 0.00030022 | 0.00467237 | -0.10796 | 0.020955 | 2.78e-07 | 3.8545390070922e-07 |
| 13961-18 | SL020291 | KIF3A | KIF3A | Q9Y496 | 0.134745 | 0.037142 | 0.000301269 | 0.004678688 | 0.177535 | 0.021326 | 1.39e-16 | 3.29387878787879e-16 |
| 5635-66 | SL017099 | TREM2 | TREM2 | Q9NZC2 | 0.102315 | 0.02824 | 0.000306888 | 0.004755785 | 0.103351 | 0.021037 | 9.57e-07 | 1.2903e-06 |
| 5988-49 | SL004648 | LIGHT | TNFSF14 | O43557 | 0.132558 | 0.036609 | 0.000309414 | 0.004784727 | 0.051646 | 0.019163 | 0.007084 | 0.007868653 |
| 6451-64 | SL008299 | ASPN | ASPN | Q9BXN1 | -0.12783 | 0.035338 | 0.000313512 | 0.004837797 | -0.15406 | 0.020438 | 6.68e-14 | 1.336e-13 |
| 22578-17 | SL007680 | ROBO2 | ROBO2 | Q9HCK4 | -0.13052 | 0.036115 | 0.000317528 | 0.004889391 | -0.14962 | 0.020061 | 1.21e-13 | 2.4076844783715e-13 |
| 12446-49 | SL005253 | GST A1-1 | GSTA1 | P08263 | 0.128931 | 0.035825 | 0.000336357 | 0.005161287 | 0.186429 | 0.021025 | 1.42e-18 | 3.80287671232877e-18 |
| 5677-15 | SL017102 | CB066 | C2orf66 | Q6UXQ4 | 0.13406 | 0.037253 | 0.000336672 | 0.005161287 | 0.08207 | 0.018665 | 1.15e-05 | 1.46227642276423e-05 |
| 15321-8 | SL019971 | CPLX2 | CPLX2 | Q6PUV4 | -0.12339 | 0.034292 | 0.000337316 | 0.005161287 | -0.2012 | 0.017676 | 2.77e-29 | 1.4345298013245e-28 |
| 19581-15 | SL004676 | IGFBP-5 | IGFBP5 | P24593 | -0.12892 | 0.035861 | 0.000341403 | 0.005212844 | -0.22648 | 0.020917 | 1.02e-26 | 4.71976331360947e-26 |
| 14114-18 | SL019019 | PIANP | PIANP | Q8IYJ0 | -0.1275 | 0.035504 | 0.000346454 | 0.005278877 | -0.18446 | 0.018409 | 3.48e-23 | 1.28974407582938e-22 |
| 9197-4 | SL008486 | LEG9 | LGALS9 | O00182 | 0.127721 | 0.03572 | 0.000367291 | 0.005577616 | 0.141355 | 0.019317 | 3.4e-13 | 6.5487684729064e-13 |
| 2890-59 | SL003303 | CCL28 | CCL28 | Q9NRJ3 | -0.13316 | 0.037243 | 0.000367595 | 0.005577616 | -0.17365 | 0.020439 | 3.36e-17 | 8.34133333333333e-17 |
| 21768-9 | SL007374 | MGT5A | MGAT5 | Q09328 | -0.12566 | 0.035176 | 0.000372062 | 0.005619534 | -0.11145 | 0.020721 | 8.23e-08 | 1.17872893772894e-07 |
| 20120-101 | SL025790 | AMGO1:ECD | AMIGO1 | Q86WK6 | -0.13224 | 0.03703 | 0.000373568 | 0.005619534 | -0.11283 | 0.018081 | 5.14e-10 | 8.18631364562118e-10 |
| 3044-3 | SL003323 | PARC | CCL18 | P55774 | 0.123632 | 0.034624 | 0.000374213 | 0.005619534 | 0.239972 | 0.020506 | 8.13e-31 | 4.57385611510791e-30 |
| 16890-37 | SL012697 | ATL1 | ADAMTSL1 | Q8N6G6 | -0.10235 | 0.028665 | 0.000374525 | 0.005619534 | -0.2236 | 0.019664 | 3.13e-29 | 1.61030263157895e-28 |
| 21182-8 | SL022269 | SEN15 | TSEN15 | Q8WW01 | -0.12677 | 0.035505 | 0.000374614 | 0.005619534 | -0.09405 | 0.018772 | 5.82e-07 | 7.92898954703833e-07 |
| 12807-89 | SL019940 | RHG30 | ARHGAP30 | Q7Z6I6 | 0.132969 | 0.037251 | 0.000375759 | 0.005619534 | 0.147828 | 0.017303 | 2.26e-17 | 5.64638977635783e-17 |
| 22010-36 | SL022185 | PRKRA | PRKRA | O75569 | 0.130656 | 0.036603 | 0.00037577 | 0.005619534 | 0.104054 | 0.021654 | 1.64e-06 | 2.18108843537415e-06 |
| 6544-33 | SL012542 | NELL1 | NELL1 | Q92832 | -0.13104 | 0.03675 | 0.000381302 | 0.005690568 | -0.14461 | 0.020946 | 6.43e-12 | 1.15592183908046e-11 |
| 13561-5 | SL020176 | 5HT6R | HTR6 | P50406 | 0.133157 | 0.037368 | 0.00038463 | 0.005728469 | 0.153701 | 0.019645 | 7.56e-15 | 1.59350943396226e-14 |
| 8289-8 | SL025876 | GPNMB:ECD | GPNMB | Q14956 | -0.12886 | 0.036183 | 0.000387654 | 0.005761697 | -0.06515 | 0.020507 | 0.001507 | 0.001710185 |
| 21999-61 | SL013068 | NUCB2 | NUCB2 | P80303 | 0.130572 | 0.03668 | 0.000389946 | 0.005783936 | 0.081179 | 0.018642 | 1.39e-05 | 1.75602584814216e-05 |
| 23173-3 | SL000591 | TIMP-1 | TIMP1 | P01033 | 0.115554 | 0.032485 | 0.000393832 | 0.005829675 | 0.130707 | 0.019341 | 1.74e-11 | 3.0170288248337e-11 |
| 6617-12 | SL017506 | FCRL6 | FCRL6 | Q6DN72 | 0.130669 | 0.03676 | 0.000397427 | 0.005870931 | 0.156498 | 0.020452 | 2.83e-14 | 5.76317708333333e-14 |
| 20928-39 | SL022251 | TCAL7 | TCEAL7 | Q9BRU2 | -0.13343 | 0.037543 | 0.000398455 | 0.005874178 | -0.05607 | 0.018749 | 0.002811 | 0.003157774 |
| 15441-6 | SL008548 | SAP3 | GM2A | P17900 | 0.111667 | 0.031441 | 0.000402087 | 0.005915725 | 0.169602 | 0.017161 | 1.3e-22 | 4.66330275229358e-22 |
| 13268-45 | SL012434 | WNT5A | WNT5A | P41221 | 0.128049 | 0.036079 | 0.000405763 | 0.005957744 | 0.040626 | 0.021009 | 0.05326 | 0.057447338 |
| 5671-1 | SL000353 | Chymotrypsin | CTRB1 | P17538 | -0.1327 | 0.037467 | 0.000417075 | 0.006111488 | -0.06371 | 0.021317 | 0.002828 | 0.003172878 |
| 4915-64 | SL000836 | Hemoglobin | HBA1\|HBB | P69905\|P68871 | -0.12901 | 0.036467 | 0.000423263 | 0.006189692 | -0.11898 | 0.019711 | 1.82e-09 | 2.82388888888889e-09 |
| 2762-30 | SL004337 | FGF-19 | FGF19 | O95750 | -0.13059 | 0.036991 | 0.00043515 | 0.006350739 | -0.0954 | 0.021513 | 9.63e-06 | 1.22649185667752e-05 |
| 24931-9 | SL006672 | ES8L3 | EPS8L3 | Q8TE67 | -0.13236 | 0.03751 | 0.000438075 | 0.006380624 | -0.0559 | 0.018014 | 0.001938 | 0.00218723 |
| 17760-128 | SL020928 | WDR5 | WDR5 | P61964 | 0.129805 | 0.036955 | 0.000465159 | 0.006761555 | 0.111209 | 0.01688 | 5.43e-11 | 9.05385927505331e-11 |
| 10445-20 | SL004747 | ApoM | APOM | O95445 | -0.12569 | 0.035846 | 0.000475789 | 0.006902268 | -0.15981 | 0.020783 | 2.13e-14 | 4.3603664921466e-14 |
| 6404-20 | SL012386 | C1QRF | C1QL1 | O75973 | -0.12067 | 0.03445 | 0.000482133 | 0.006980362 | -0.10618 | 0.020634 | 2.87e-07 | 3.96526501766784e-07 |
| 6227-1 | SL003923 | kallikrein 10 | KLK10 | O43240 | -0.12845 | 0.036795 | 0.000503968 | 0.007276632 | -0.20233 | 0.020405 | 9.55e-23 | 3.44152073732719e-22 |
| 6454-38 | SL005264 | Rho-GDIa | ARHGDIA | P52565 | 0.130787 | 0.037469 | 0.000504599 | 0.007276632 | 0.092337 | 0.019676 | 2.84e-06 | 3.72006700167504e-06 |
| 9754-33 | SL006251 | Quinone reductase 2 | NQO2 | P16083 | -0.12838 | 0.036858 | 0.000518694 | 0.007465089 | -0.0982 | 0.020081 | 1.07e-06 | 1.43769759450172e-06 |
| 2972-57 | SL004078 | BMP-7 | BMP7 | P18075 | -0.13065 | 0.037545 | 0.000524856 | 0.00753884 | -0.13629 | 0.01952 | 3.73e-12 | 6.79920745920746e-12 |
| 4162-54 | SL000601 | Transferrin | TF | P02787 | -0.12911 | 0.037188 | 0.000540859 | 0.007753382 | -0.12258 | 0.021486 | 1.3e-08 | 1.94378585086042e-08 |
| 19639-53 | SL011501 | IAPP | IAPP | P10997 | -0.1217 | 0.035135 | 0.000556644 | 0.00796396 | -0.19102 | 0.020272 | 9.86e-21 | 3.05973015873016e-20 |
| 15426-5 | SL008692 | NEUR1 | NEU1 | Q99519 | 0.126322 | 0.036488 | 0.000560561 | 0.008004235 | 0.160813 | 0.018592 | 9.2e-18 | 2.35111111111111e-17 |
| 16914-104 | SL001743 | sCD14 | CD14 | P08571 | -0.11848 | 0.034258 | 0.000568007 | 0.008079282 | -0.15373 | 0.019037 | 1.05e-15 | 2.346e-15 |
| 21813-171 | SL007374 | MGT5A | MGAT5 | Q09328 | -0.12308 | 0.035589 | 0.00056804 | 0.008079282 | -0.10335 | 0.02083 | 7.46e-07 | 1.01104332755633e-06 |
| 3041-55 | SL008416 | MRC2 | MRC2 | Q9UBG0 | -0.1263 | 0.036535 | 0.000570879 | 0.008103808 | -0.10861 | 0.021049 | 2.67e-07 | 3.70859680284192e-07 |
| 6416-8 | SL017435 | GKN2 | GKN2 | Q86XP6 | -0.12383 | 0.035924 | 0.000591976 | 0.008386908 | -0.0814 | 0.020955 | 0.000105 | 0.00012579 |
| 16919-1 | SL008480 | ACBP | DBI | P07108 | 0.128928 | 0.037443 | 0.000600086 | 0.008485264 | 0.160496 | 0.02105 | 3.48e-14 | 7.06846753246753e-14 |
| 2696-87 | SL004685 | Persephin | PSPN | O60542 | 0.110762 | 0.0323 | 0.000631987 | 0.008918999 | 0.098818 | 0.018572 | 1.13e-07 | 1.6037386569873e-07 |
| 20549-1 | SL021683 | IZUM4 | IZUMO4 | Q1ZYL8 | 0.120424 | 0.035128 | 0.000634209 | 0.008933009 | 0.045191 | 0.021079 | 0.032136 | 0.035000164 |
| 15364-101 | SL000275 | Apo C-I | APOC1 | P02654 | -0.1249 | 0.036454 | 0.000638467 | 0.008959775 | -0.20061 | 0.020627 | 5.84e-22 | 1.96003433476395e-21 |
| 16770-3 | SL012438 | REG1B | REG1B | P48304 | -0.12051 | 0.035172 | 0.000638575 | 0.008959775 | -0.08037 | 0.020858 | 0.00012 | 0.000142139 |
| 5803-24 | SL003362 | C3d | C3 | P01024 | 0.127945 | 0.037354 | 0.000641106 | 0.00896996 | 0.201007 | 0.020961 | 2.1e-21 | 6.67560975609756e-21 |
| 5491-12 | SL010471 | Testican-2 | SPOCK2 | Q92563 | -0.128 | 0.037375 | 0.000641769 | 0.00896996 | -0.1454 | 0.021158 | 8.01e-12 | 1.42359545454545e-11 |
| 10907-116 | SL009074 | NTRI | NTM | Q9P121 | -0.12139 | 0.035475 | 0.000649101 | 0.009055016 | -0.18966 | 0.019405 | 3.7e-22 | 1.28026548672566e-21 |
| 6965-19 | SL017948 | CNTP2 | CNTNAP2 | Q9UHC6 | -0.12251 | 0.035831 | 0.000655408 | 0.009125496 | -0.12514 | 0.019163 | 7.96e-11 | 1.3215966029724e-10 |
| 13095-51 | SL005357 | PSP | REG1A | P05451 | -0.12057 | 0.035314 | 0.000666877 | 0.009267417 | -0.08541 | 0.020071 | 2.17e-05 | 2.71076677316294e-05 |
| 7131-207 | SL003198 | Tenascin | TNC | P24821 | -0.12739 | 0.037316 | 0.000668392 | 0.009270752 | -0.07618 | 0.021731 | 0.000464 | 0.000536179 |
| 2900-53 | SL003329 | HCC-1 | CCL14 | Q16627 | 0.118462 | 0.034727 | 0.000674441 | 0.00933683 | 0.131227 | 0.01905 | 7.14e-12 | 1.27186332574032e-11 |
| 20091-138 | SL004138 | Ephrin-A1 | EFNA1 | P20827 | 0.115383 | 0.033875 | 0.000686939 | 0.009491776 | 0.102896 | 0.01759 | 5.58e-09 | 8.522578125e-09 |
| 3314-74 | SL004858 | GFRa-1 | GFRA1 | P56159 | 0.122439 | 0.036052 | 0.000712251 | 0.009822852 | 0.215291 | 0.019268 | 2.65e-28 | 1.33696774193548e-27 |
| 3624-3 | SL011069 | Marapsin | PRSS27 | Q9BQR3 | 0.12289 | 0.036197 | 0.000715117 | 0.009843699 | 0.057827 | 0.018232 | 0.001534 | 0.00173842 |
| 21328-2 | SL020091 | SYT1 | SYT1 | P21579 | -0.12574 | 0.037043 | 0.000716565 | 0.009844981 | -0.03329 | 0.01891 | 0.078484 | 0.083730223 |
| 14208-3 | SL019932 | RET7 | RBP7 | Q96R05 | 0.126149 | 0.037177 | 0.00071979 | 0.009870628 | 0.117021 | 0.019919 | 4.81e-09 | 7.36090019569472e-09 |
| 9506-10 | SL005699 | Apo L1 | APOL1 | O14791 | -0.12168 | 0.035865 | 0.000721172 | 0.009870956 | -0.10208 | 0.020533 | 7.11e-07 | 9.6528125e-07 |
| 22972-26 | SL003174 | Gro-b | CXCL2 | P19875 | 0.127279 | 0.03754 | 0.000727023 | 0.009932337 | -0.01225 | 0.021984 | 0.577515 | 0.587277424 |
| 25902-27 | SL007997 | DHE3 | GLUD1 | P00367 | 0.125311 | 0.037022 | 0.000742026 | 0.010118279 | 0.11331 | 0.019275 | 4.7e-09 | 7.20666666666667e-09 |
| 12501-10 | SL008849 | TBCA | TBCA | O75347 | -0.12415 | 0.036701 | 0.000747777 | 0.010177611 | -0.07754 | 0.021733 | 0.000367 | 0.000427999 |
| 10851-77 | SL007417 | IL27B | EBI3 | Q14213 | -0.1187 | 0.035132 | 0.0007587 | 0.010295772 | -0.0713 | 0.020218 | 0.000429 | 0.000497278 |
| 10974-20 | SL012454 | ISK7 | SPINK7 | P58062 | -0.11559 | 0.034214 | 0.000759292 | 0.010295772 | -0.14738 | 0.019086 | 1.66e-14 | 3.41610526315789e-14 |
| 5124-69 | SL005169 | sICAM-5 | ICAM5 | Q9UMF0 | -0.11794 | 0.035031 | 0.000791517 | 0.010712744 | -0.13946 | 0.019397 | 8.59e-13 | 1.61475480769231e-12 |
| 21690-31 | SL022651 | LRRC4 | LRRC4 | Q9HBW1 | -0.12634 | 0.037573 | 0.000804002 | 0.010861498 | -0.04568 | 0.021662 | 0.03505 | 0.038121032 |
| 15613-16 | SL012420 | LIPP | PNLIP | P16233 | -0.1254 | 0.037344 | 0.000816502 | 0.011009907 | -0.07887 | 0.021236 | 0.000209 | 0.000246041 |
| 17404-5 | SL019712 | ARL5B | ARL5B | Q96KC2 | 0.125185 | 0.037291 | 0.000819683 | 0.01103233 | 0.155644 | 0.020982 | 1.63e-13 | 3.21073047858942e-13 |
| 20564-53 | SL007075 | THY1 | THY1 | P04216 | 0.122062 | 0.036407 | 0.000832478 | 0.011176956 | 0.144548 | 0.017056 | 4.01e-17 | 9.92348101265823e-17 |
| 14711-27 | SL004438 | Cystatin M | CST6 | Q15828 | -0.11275 | 0.033632 | 0.000833504 | 0.011176956 | -0.18629 | 0.019874 | 1.54e-20 | 4.70421875e-20 |
| 6597-24 | SL008903 | TM157 | FAM174A | Q8TBP5 | -0.12514 | 0.037335 | 0.000835168 | 0.011178637 | -0.03959 | 0.019242 | 0.039754 | 0.04317737 |
| 14041-13 | SL004068 | Granzyme B | GZMB | P10144 | -0.12591 | 0.037603 | 0.00084504 | 0.011289641 | -0.01517 | 0.018321 | 0.407759 | 0.419562962 |
| 6645-53 | SL005084 | Periostin | POSTN | Q15063 | -0.11938 | 0.035659 | 0.000847233 | 0.011289641 | -0.19315 | 0.020856 | 4.31e-20 | 1.239125e-19 |
| 23326-10 | SL009143 | GSTA2 | GSTA2 | P09210 | 0.121728 | 0.036364 | 0.000848121 | 0.011289641 | 0.194029 | 0.020838 | 2.73e-20 | 8.05607547169811e-20 |
| 22500-9 | SL022881 | MED11 | MED11 | Q9P086 | -0.12397 | 0.037131 | 0.000874529 | 0.011619888 | -0.23144 | 0.019276 | 2.62e-32 | 1.5882480620155e-31 |
| 6557-50 | SL008774 | LRC15 | LRRC15 | Q8TF66 | -0.1215 | 0.036403 | 0.000878532 | 0.011651769 | -0.10305 | 0.020608 | 6.12e-07 | 8.3232e-07 |
| 8028-22 | SL009201 | SPINK5 | SPINK5 | Q9NQ38 | -0.11995 | 0.035977 | 0.000889133 | 0.011770895 | -0.15083 | 0.02058 | 3.13e-13 | 6.05856435643564e-13 |
| 5480-49 | SL000563 | RANTES | CCL5 | P13501 | 0.122966 | 0.036911 | 0.000897954 | 0.011866058 | -0.01401 | 0.02204 | 0.525153 | 0.53682276 |
| 3806-55 | SL004844 | EphA5 | EPHA5 | P54756 | -0.12172 | 0.036601 | 0.000916684 | 0.012091574 | -0.1009 | 0.019329 | 1.94e-07 | 2.72366247755835e-07 |
| 10462-14 | SL012880 | INSL5 | INSL5 | Q9Y5Q6 | 0.123704 | 0.037238 | 0.000928496 | 0.012225197 | 0.060423 | 0.019817 | 0.002321 | 0.002611205 |
| 10620-21 | SL000548 | PSP-94 | MSMB | P08118 | -0.10771 | 0.03244 | 0.000933676 | 0.012267984 | -0.09343 | 0.020541 | 5.67e-06 | 7.31673267326733e-06 |
| 24490-16 | SL023768 | THAP4 | THAP4 | Q8WY91 | -0.10743 | 0.032357 | 0.000935121 | 0.012267984 | -0.07908 | 0.019307 | 4.34e-05 | 5.28641744548287e-05 |
| 6448-36 | SL003688 | Sema E | SEMA3C | Q99985 | 0.118346 | 0.035676 | 0.00094412 | 0.012363725 | 0.097621 | 0.018769 | 2.14e-07 | 2.9937030411449e-07 |
| 8606-39 | SL025877 | GPNMB:CD | GPNMB | Q14956 | -0.12148 | 0.036645 | 0.000951734 | 0.012441015 | -0.15558 | 0.020908 | 1.38e-13 | 2.73205063291139e-13 |
| 19341-36 | SL019254 | ACBD6 | ACBD6 | Q9BR61 | 0.118763 | 0.03591 | 0.000978087 | 0.01274686 | 0.169568 | 0.021821 | 1.14e-14 | 2.37095744680851e-14 |
| 13712-104 | SL004298 | granzyme A | GZMA | P12544 | 0.122859 | 0.03715 | 0.000978639 | 0.01274686 | 0.153912 | 0.02055 | 9.6e-14 | 1.91510204081633e-13 |
| 6060-2 | SL000060 | PIP | PIP | P12273 | -0.11105 | 0.033596 | 0.000984784 | 0.012799019 | -0.07054 | 0.020916 | 0.000756 | 0.000865581 |
| 5586-66 | SL007807 | MINP1 | MINPP1 | Q9UNW1 | -0.1224 | 0.037038 | 0.000986588 | 0.012799019 | -0.03705 | 0.02053 | 0.071276 | 0.076249172 |
| 18332-17 | SL020907 | CPLX1 | CPLX1 | O14810 | -0.11474 | 0.034723 | 0.000987927 | 0.012799019 | -0.11859 | 0.017839 | 3.65e-11 | 6.205e-11 |
| 11440-58 | SL004268 | SOCS-3 | SOCS3 | O14543 | 0.114832 | 0.034758 | 0.000990393 | 0.012807121 | 0.114317 | 0.017749 | 1.43e-10 | 2.33945606694561e-10 |
| 14032-2 | SL003310 | VEGF121 | VEGFA | P15692 | 0.123974 | 0.037536 | 0.00099372 | 0.012807121 | 0.018734 | 0.019629 | 0.339978 | 0.351205601 |
| 6470-19 | SL004461 | fibulin 1 | FBLN1 | P23142 | -0.12192 | 0.036915 | 0.000993838 | 0.012807121 | -0.18251 | 0.020782 | 2.96e-18 | 7.76751677852349e-18 |
| 11573-3 | SL017909 | SRSF6 | SRSF6 | Q13247 | 0.121031 | 0.03667 | 0.001001671 | 0.012885211 | 0.181954 | 0.018847 | 1.15e-21 | 3.71611570247934e-21 |
| 7767-1 | SL012589 | CF126 | CLPSL2 | Q6UWE3 | 0.120734 | 0.036854 | 0.001091572 | 0.014016868 | 0.111985 | 0.019795 | 1.72e-08 | 2.55225806451613e-08 |
| 7154-92 | SL018080 | CF058 | LEG1 | Q6P5S2 | -0.10939 | 0.033424 | 0.001104094 | 0.014152652 | -0.09022 | 0.020605 | 1.25e-05 | 1.58685064935065e-05 |
| 8841-65 | SL008847 | CILP2 | CILP2 | Q8IUL8 | -0.10819 | 0.033065 | 0.001106683 | 0.014160865 | -0.24768 | 0.020468 | 8.89e-33 | 5.474e-32 |
| 23414-12 | SL023681 | ZN483 | ZNF483 | Q8TF39 | -0.12163 | 0.037197 | 0.001115742 | 0.014250408 | -0.1892 | 0.019461 | 6.06e-22 | 2.00801694915254e-21 |
| 11851-21 | SL017099 | TREM2 | TREM2 | Q9NZC2 | 0.096911 | 0.029643 | 0.001117602 | 0.014250408 | 0.086221 | 0.02094 | 3.96e-05 | 4.84619718309859e-05 |
| 12678-66 | SL013011 | SNRPA | SNRPA | P09012 | 0.12269 | 0.037549 | 0.001124697 | 0.01431576 | 0.12393 | 0.020377 | 1.38e-09 | 2.16264529058116e-09 |
| 2523-31 | SL000563 | RANTES | CCL5 | P13501 | 0.121537 | 0.037203 | 0.001127071 | 0.014320893 | -0.01208 | 0.022033 | 0.583418 | 0.592510432 |
| 4563-61 | SL012108 | PLCG1 | PLCG1 | P19174 | 0.119538 | 0.036598 | 0.001129777 | 0.014330225 | 0.069854 | 0.019908 | 0.000458 | 0.000530949 |
| 11200-52 | SL007696 | C1QR1 | CD93 | Q9NPY3 | -0.11414 | 0.03499 | 0.001146029 | 0.014511045 | -0.1068 | 0.019923 | 9.07e-08 | 1.29429562043796e-07 |
| 2986-49 | SL003175 | Gro-g | CXCL3 | P19876 | 0.122136 | 0.037459 | 0.001152449 | 0.014566958 | -0.00992 | 0.021961 | 0.651576 | 0.660016408 |
| 13123-3 | SL025865 | FLRT3:ECD | FLRT3 | Q9NZU0 | -0.11375 | 0.0349 | 0.00115772 | 0.014596557 | -0.09668 | 0.021202 | 5.37e-06 | 6.94105785123967e-06 |
| 2973-15 | SL000668 | CD36 ANTIGEN | CD36 | P16671 | 0.119393 | 0.036636 | 0.001158808 | 0.014596557 | 0.040607 | 0.020135 | 0.043831 | 0.047473465 |
| 8007-19 | SL000343 | Cathepsin B | CTSB | P07858 | -0.10748 | 0.033016 | 0.001173068 | 0.014750623 | -0.09616 | 0.02058 | 3.14e-06 | 4.09246666666667e-06 |
| 22402-12 | SL022926 | H2A1A | H2AC1 | Q96QV6 | 0.115853 | 0.035654 | 0.001198061 | 0.015038879 | 0.145199 | 0.02074 | 3.27e-12 | 6.00267605633803e-12 |
| 12572-236 | SL019783 | EFS | EFS | O43281 | 0.120879 | 0.03722 | 0.001204894 | 0.015098564 | 0.110824 | 0.018282 | 1.56e-09 | 2.43984e-09 |
| 5124-62 | SL005169 | sICAM-5 | ICAM5 | Q9UMF0 | -0.1124 | 0.034632 | 0.001214068 | 0.015187341 | -0.14168 | 0.019233 | 2.39e-13 | 4.68416040100251e-13 |
| 21533-51 | SL014775 | LSP1 | LSP1 | P33241 | 0.115641 | 0.035666 | 0.001227638 | 0.015310138 | 0.132532 | 0.017518 | 5.44e-14 | 1.09078974358974e-13 |
| 16613-3 | SL008898 | CAD17 | CDH17 | Q12864 | -0.11507 | 0.035499 | 0.001231813 | 0.015310138 | -0.06789 | 0.02078 | 0.001102 | 0.001257935 |
| 21771-47 | SL022690 | MGAT3 | MGAT3 | Q09327 | -0.11409 | 0.035197 | 0.001232037 | 0.015310138 | -0.04672 | 0.020498 | 0.022731 | 0.024966119 |
| 15385-116 | SL004848 | FABP2 | FABP2 | P12104 | -0.12181 | 0.037581 | 0.00123231 | 0.015310138 | -0.01812 | 0.022078 | 0.412014 | 0.423383535 |
| 11416-23 | SL025857 | FBXL4:LRR4 and LRR5 | FBXL4 | Q9UKA2 | -0.1216 | 0.037534 | 0.001238264 | 0.015357859 | -0.06835 | 0.020052 | 0.000664 | 0.000762364 |
| 12630-8 | SL019787 | ARFP2 | ARFIP2 | P53365 | -0.1185 | 0.036676 | 0.001277137 | 0.015812996 | -0.0712 | 0.020425 | 0.000499 | 0.000576046 |
| 6107-3 | SL012871 | ELA1 | CELA1 | Q9UNI1 | -0.11807 | 0.036623 | 0.00130783 | 0.016165485 | -0.08974 | 0.021588 | 3.34e-05 | 4.12619273301738e-05 |
| 13724-27 | SL004337 | FGF-19 | FGF19 | O95750 | -0.11903 | 0.037015 | 0.001345894 | 0.016607742 | -0.08865 | 0.021427 | 3.63e-05 | 4.46330188679245e-05 |
| 8059-1 | SL000598 | Tpo | THPO | P40225 | -0.11996 | 0.03735 | 0.001363961 | 0.016798819 | -0.07727 | 0.017179 | 7.18e-06 | 9.21963875205255e-06 |
| 22047-46 | SL007149 | CO5A1 | COL5A1 | P20908 | 0.10686 | 0.033277 | 0.001366938 | 0.016798819 | 0.129797 | 0.020927 | 6.51e-10 | 1.03053036437247e-09 |
| 8653-132 | SL025839 | DNJC4:C-term | DNAJC4 | Q9NNZ3 | 0.118006 | 0.036752 | 0.001368313 | 0.016798819 | 0.087653 | 0.021254 | 3.85e-05 | 4.71896551724138e-05 |
| 4989-7 | SL003341 | Fibrinogen g-chain dimer | FGG | P02679 | 0.112288 | 0.035029 | 0.001393795 | 0.017082801 | 0.166264 | 0.020492 | 7.71e-16 | 1.73752737752161e-15 |
| 21498-3 | SL008882 | AL7A1 | ALDH7A1 | P49419 | 0.118653 | 0.037056 | 0.001410953 | 0.017263987 | 0.185675 | 0.019895 | 2.24e-20 | 6.68580152671756e-20 |
| 18397-5 | SL014729 | AK1C4 | AKR1C4 | P17516 | 0.110167 | 0.034428 | 0.001421007 | 0.017357783 | 0.191621 | 0.020508 | 2.03e-20 | 6.10561538461538e-20 |
| 15525-294 | SL014682 | ADH1G | ADH1C | P00326 | 0.115613 | 0.036152 | 0.001430338 | 0.017442447 | 0.165222 | 0.021117 | 7.56e-15 | 1.59350943396226e-14 |
| 11381-56 | SL010934 | PRPS1 | PRPS1 | P60891 | -0.12016 | 0.037595 | 0.001439096 | 0.017519853 | -0.02331 | 0.018666 | 0.211877 | 0.222399226 |
| 3891-56 | SL009629 | MBD4 | MBD4 | O95243 | 0.119961 | 0.037595 | 0.001465953 | 0.017795386 | -0.0203 | 0.017831 | 0.255029 | 0.266977836 |
| 15487-164 | SL006149 | carboxylesterase, liver | CES1 | P23141 | 0.117675 | 0.036884 | 0.001468168 | 0.017795386 | 0.135897 | 0.020623 | 5.39e-11 | 9.00636752136752e-11 |
| 7947-19 | SL018328 | AP4AT | TEPSIN | Q96N21 | -0.11957 | 0.037484 | 0.00147102 | 0.017795386 | -0.03306 | 0.017792 | 0.063254 | 0.068017028 |
| 17346-61 | SL012716 | FDSCP | FDCSP | Q8NFU4 | -0.11897 | 0.037297 | 0.001471523 | 0.017795386 | -0.11441 | 0.019971 | 1.14e-08 | 1.71109404990403e-08 |
| 3366-51 | SL006550 | ECM1 | ECM1 | Q16610 | -0.11435 | 0.035874 | 0.001482216 | 0.01789493 | -0.09013 | 0.021607 | 3.13e-05 | 3.87901743264659e-05 |
| 13510-7 | SL011088 | AT2A3 | ATP2A3 | Q93084 | -0.11934 | 0.037449 | 0.001486153 | 0.017908688 | -0.08641 | 0.016992 | 3.95e-07 | 5.43820422535211e-07 |
| 8231-122 | SL003321 | VEGF sR1 | FLT1 | P17948 | -0.1189 | 0.037315 | 0.001488284 | 0.017908688 | -0.12275 | 0.019891 | 7.92e-10 | 1.24867741935484e-09 |
| 18304-19 | SL021083 | BCCIP | BCCIP | Q9P287 | 0.11855 | 0.037256 | 0.001510806 | 0.018149644 | 0.01946 | 0.016397 | 0.235409 | 0.246768696 |
| 9350-3 | SL012577 | FSTL4 | FSTL4 | Q6MZW2 | 0.118562 | 0.037323 | 0.001538879 | 0.018456386 | 0.017523 | 0.01744 | 0.31513 | 0.325967494 |
| 19575-4 | SL005159 | EPO-R | EPOR | P19235 | 0.11116 | 0.034998 | 0.001541431 | 0.018456546 | 0.086681 | 0.018249 | 2.15e-06 | 2.84483925549915e-06 |
| 21811-20 | SL022296 | NO40 | ZCCHC17 | Q9NP64 | 0.116622 | 0.036746 | 0.001554014 | 0.0185766 | 0.096927 | 0.019573 | 7.84e-07 | 1.06070588235294e-06 |
| 7140-1 | SL012410 | ELA2A | CELA2A | P08217 | -0.11946 | 0.03767 | 0.001567305 | 0.018676179 | -0.0895 | 0.021508 | 3.28e-05 | 4.05848101265823e-05 |
| 2654-19 | SL001992 | TNF sR-I | TNFRSF1A | P19438 | 0.096854 | 0.030545 | 0.001569611 | 0.018676179 | 0.197063 | 0.018838 | 4.39e-25 | 1.82605319148936e-24 |
| 4775-34 | SL005572 | Gelsolin | GSN | P06396 | -0.11908 | 0.037555 | 0.001570053 | 0.018676179 | -0.16203 | 0.019877 | 5.66e-16 | 1.28666279069767e-15 |
| 9183-7 | SL004475 | IFN-a/b R1 | IFNAR1 | P17181 | -0.11841 | 0.037374 | 0.001582993 | 0.018799335 | -0.11181 | 0.018757 | 2.86e-09 | 4.40259842519685e-09 |
| 23581-131 | SL023012 | Obestatin | GHRL | Q9UBU3 | -0.10957 | 0.034687 | 0.001634787 | 0.019382763 | -0.19803 | 0.019129 | 1.31e-24 | 5.3634554973822e-24 |
| 24422-36 | SL023761 | SPA24 | SPATA24 | Q86W54 | -0.11773 | 0.037294 | 0.001646511 | 0.019489969 | -0.1183 | 0.020779 | 1.4e-08 | 2.08533333333333e-08 |
| 3009-3 | SL005059 | TGF-b R III | TGFBR3 | Q03167 | -0.11414 | 0.036248 | 0.001690876 | 0.019982584 | -0.09089 | 0.020441 | 9.13e-06 | 1.16470799347471e-05 |
| 8245-27 | SL005169 | sICAM-5 | ICAM5 | Q9UMF0 | -0.10853 | 0.034496 | 0.001706397 | 0.020133266 | -0.14132 | 0.019544 | 6.38e-13 | 1.21390754257908e-12 |
| 11142-11 | SL025792 | ANGL1:C-term | ANGPTL1 | O95841 | -0.1159 | 0.03687 | 0.001722369 | 0.020288786 | -0.19529 | 0.020899 | 2.02e-20 | 6.09899613899614e-20 |
| 10978-39 | SL009327 | SOM2 | GH2 | P01242 | -0.10625 | 0.033822 | 0.001733632 | 0.020374205 | -0.21945 | 0.020248 | 9.13e-27 | 4.24979761904762e-26 |
| 20941-7 | SL022284 | UBL3 | UBL3 | O95164 | -0.11786 | 0.03752 | 0.001735227 | 0.020374205 | -0.08028 | 0.021327 | 0.000171 | 0.000203154 |
| 16858-384 | SL017265 | RCN3 | RCN3 | Q96D15 | -0.11829 | 0.037678 | 0.001744902 | 0.020442258 | -0.15021 | 0.019502 | 1.93e-14 | 3.96131233595801e-14 |
| 10637-50 | SL025903 | K1324:ECD | ELAPOR1 | Q6UXG2 | -0.11723 | 0.037344 | 0.001747612 | 0.020442258 | -0.13483 | 0.018735 | 8.16e-13 | 1.53761927710843e-12 |
| 3538-26 | SL009216 | dopa decarboxylase | DDC | P20711 | -0.11704 | 0.037288 | 0.001749461 | 0.020442258 | -0.11861 | 0.017582 | 1.89e-11 | 3.26986725663717e-11 |
| 21891-31 | SL022635 | FBLN7 | FBLN7 | Q53RD9 | -0.11736 | 0.037402 | 0.001756387 | 0.020490238 | -0.15444 | 0.021054 | 2.99e-13 | 5.81636815920398e-13 |
| 8587-21 | SL012570 | ISK52 | SPINK14 | Q6IE38 | 0.111973 | 0.035705 | 0.001766069 | 0.020570173 | 0.042223 | 0.019299 | 0.028777 | 0.031429082 |
| 4968-50 | SL008099 | CAPG | CAPG | P40121 | 0.104978 | 0.03349 | 0.001774722 | 0.020637882 | 0.111366 | 0.019046 | 5.67e-09 | 8.64315789473684e-09 |
| 3340-53 | SL007207 | TSP4 | THBS4 | P35443 | 0.110142 | 0.035148 | 0.00178044 | 0.020671304 | 0.161352 | 0.020426 | 4.21e-15 | 8.99513661202186e-15 |
| 11353-143 | SL004101 | SMAD2 | SMAD2 | Q15796 | 0.11611 | 0.037074 | 0.001791482 | 0.020741476 | 0.072085 | 0.019284 | 0.00019 | 0.000224426 |
| 4314-12 | SL011809 | XTP3A | DCTPP1 | Q9H773 | -0.1097 | 0.03503 | 0.001792191 | 0.020741476 | -0.09205 | 0.021485 | 1.9e-05 | 2.3887459807074e-05 |
| 2797-56 | SL000020 | Apo B | APOB | P04114 | -0.11561 | 0.036993 | 0.001832418 | 0.021164411 | -0.13387 | 0.021308 | 3.93e-10 | 6.29766393442623e-10 |
| 9876-20 | SL004727 | aldolase C | ALDOC | P09972 | -0.10767 | 0.034456 | 0.00183456 | 0.021164411 | -0.01261 | 0.021122 | 0.550676 | 0.560714772 |
| 8942-2 | SL018771 | RM21 | MRPL21 | Q7Z2W9 | 0.11484 | 0.036766 | 0.001842067 | 0.021217342 | 0.120142 | 0.021169 | 1.55e-08 | 2.30437262357414e-08 |
| 24907-3 | SL023621 | MPPA | PMPCA | Q10713 | 0.115041 | 0.036887 | 0.001872074 | 0.021511189 | 0.19148 | 0.020346 | 1.1e-20 | 3.4e-20 |
| 12867-40 | SL019922 | DYLT3 | DYNLT3 | P51808 | 0.117413 | 0.037651 | 0.001873498 | 0.021511189 | -0.01318 | 0.0176 | 0.454111 | 0.466029616 |
| 22468-54 | SL005850 | Histone H2A type 1 | H2AC11 | P0C0S8 | 0.111264 | 0.035701 | 0.001885844 | 0.021618794 | 0.113428 | 0.021612 | 1.67e-07 | 2.35304504504504e-07 |
| 21526-88 | SL009242 | sperm-egg fusion protein 1 | IZUMO1 | Q8IYV9 | 0.115276 | 0.037 | 0.001891816 | 0.0216531 | 0.210007 | 0.021276 | 1.47e-22 | 5.22518181818182e-22 |
| 12475-48 | SL019722 | CLIC5 | CLIC5 | Q9NZA1 | -0.10251 | 0.032965 | 0.001929047 | 0.022044514 | -0.10921 | 0.0195 | 2.38e-08 | 3.51162264150943e-08 |
| 9402-18 | SL017500 | DCBD1 | DCBLD1 | Q8N8Z6 | 0.117132 | 0.037674 | 0.001933338 | 0.022058875 | -0.02972 | 0.017642 | 0.092192 | 0.098087164 |
| 23696-256 | SL023309 | TRM6 | TRMT6 | Q9UJA5 | 0.116248 | 0.037405 | 0.001941678 | 0.022119308 | 0.003157 | 0.018384 | 0.863654 | 0.866979809 |
| 6896-3 | SL017990 | CA162 | C1orf162 | Q8NEQ5 | 0.113748 | 0.036625 | 0.001955064 | 0.022211615 | 0.207909 | 0.020093 | 1.38e-24 | 5.620625e-24 |
| 21708-149 | SL014852 | FCGRN | FCGRT | P55899 | -0.108 | 0.034776 | 0.001955893 | 0.022211615 | 0.00559 | 0.020871 | 0.788864 | 0.794963262 |
| 17691-1 | SL007957 | TPP1 | TPP1 | O14773 | 0.114993 | 0.037048 | 0.001967654 | 0.022310308 | 0.072593 | 0.018403 | 8.21e-05 | 9.862089093702e-05 |
| 11157-35 | SL000451 | HSP 70 | HSPA1A | P0DMV8 | 0.116095 | 0.037432 | 0.001983351 | 0.022453262 | 0.198665 | 0.019742 | 2.28e-23 | 8.53090909090909e-23 |
| 17755-5 | SL008706 | UGDH | UGDH | O60701 | 0.114935 | 0.037083 | 0.001997435 | 0.022577543 | 0.164808 | 0.021069 | 7.66e-15 | 1.61024731182796e-14 |
| 5006-71 | SL006993 | MK13 | MAPK13 | O15264 | 0.116601 | 0.037632 | 0.002003515 | 0.022611094 | 0.021547 | 0.019219 | 0.262347 | 0.27390568 |
| 8091-16 | SL025929 | MASP3:Heavy | MASP1 | P48740 | -0.11129 | 0.035943 | 0.002018042 | 0.022739738 | -0.11992 | 0.020597 | 6.58e-09 | 9.9913786407767e-09 |
| 12503-5 | SL007451 | SPR1 | GPR68 | Q15743 | 0.116041 | 0.03749 | 0.002024978 | 0.022758605 | 0.066613 | 0.020288 | 0.00104 | 0.001189463 |
| 10603-1 | SL012419 | HIS3 | HTN3 | P15516 | -0.11101 | 0.035865 | 0.002025979 | 0.022758605 | -0.15738 | 0.020216 | 1.02e-14 | 2.12704e-14 |
| 3303-23 | SL004438 | Cystatin M | CST6 | Q15828 | -0.10125 | 0.03274 | 0.002043951 | 0.022918196 | -0.15934 | 0.019367 | 3.08e-16 | 7.12591715976331e-16 |
| 2760-2 | SL004752 | DRR1 | FAM107A | O95990 | 0.116022 | 0.037527 | 0.002049552 | 0.022918196 | 0.016617 | 0.017585 | 0.344771 | 0.355686923 |
| 20175-17 | SL012498 | CO9A3 | COL9A3 | Q14050 | -0.10632 | 0.034388 | 0.002049646 | 0.022918196 | -0.15064 | 0.018596 | 8.56e-16 | 1.92354022988506e-15 |
| 11696-7 | SL008838 | RABP2 | CRABP2 | P29373 | 0.111778 | 0.036195 | 0.002073166 | 0.023125957 | 0.221207 | 0.020251 | 3.75e-27 | 1.81018518518519e-26 |
| 9266-1 | SL004968 | sTREM-1 | TREM1 | Q9NP99 | 0.093273 | 0.030205 | 0.002074591 | 0.023125957 | 0.068444 | 0.019275 | 0.000391 | 0.000455119 |
| 17823-40 | SL013062 | SMD2 | SNRPD2 | P62316 | 0.115726 | 0.037502 | 0.002089711 | 0.023242937 | 0.030171 | 0.018975 | 0.111959 | 0.118794501 |
| 25066-32 | SL023321 | GLYG2 | GYG2 | O15488 | 0.116138 | 0.037639 | 0.002091481 | 0.023242937 | 0.006686 | 0.018037 | 0.710909 | 0.71918628 |
| 16916-19 | SL018357 | SLIK6 | SLITRK6 | Q9H5Y7 | -0.1144 | 0.037098 | 0.002104215 | 0.023348758 | -0.08283 | 0.020456 | 5.3e-05 | 6.43571428571429e-05 |
| 2683-1 | SL000456 | iC3b | C3 | P01024 | 0.114801 | 0.037265 | 0.00212603 | 0.02355486 | 0.185741 | 0.020798 | 8.15e-19 | 2.21295138888889e-18 |
| 23394-125 | SL023440 | GAS2 | GAS2 | O43903 | 0.115157 | 0.037453 | 0.002168179 | 0.023985277 | 0.143602 | 0.019903 | 7.17e-13 | 1.35761259079903e-12 |
| 2946-52 | SL003327 | Factor D | CFD | P00746 | 0.108065 | 0.035181 | 0.002190609 | 0.024196578 | 0.206938 | 0.021279 | 5.87e-22 | 1.96168376068376e-21 |
| 21696-80 | SL022657 | LRFN4 | LRFN4 | Q6PJG9 | -0.11295 | 0.036834 | 0.00222925 | 0.024586021 | -0.03119 | 0.020874 | 0.135312 | 0.143379488 |
| 3296-92 | SL008623 | CNTN2 | CNTN2 | Q02246 | -0.11149 | 0.036371 | 0.002237531 | 0.024639964 | -0.10581 | 0.021077 | 5.53e-07 | 7.56024475524476e-07 |
| 12478-15 | SL008862 | RL30 | RPL30 | P62888 | 0.113647 | 0.037126 | 0.002268582 | 0.024944105 | 0.146418 | 0.020299 | 7.26e-13 | 1.37133333333333e-12 |
| 5002-76 | SL002646 | MMP-14 | MMP14 | P50281 | 0.115088 | 0.037688 | 0.002324262 | 0.025517726 | -0.02176 | 0.020909 | 0.298209 | 0.309283489 |
| 6923-1 | SL012351 | PLOD2 | PLOD2 | O00469 | 0.112863 | 0.037037 | 0.002374182 | 0.026026483 | 0.137152 | 0.020671 | 3.99e-11 | 6.75363636363636e-11 |
| 24659-6 | SL009432 | DJB13 | DNAJB13 | P59910 | 0.113556 | 0.037272 | 0.002378849 | 0.026038368 | 0.059003 | 0.017159 | 0.000594 | 0.000684567 |
| 24638-3 | SL023635 | LRA25 | FAM89B | Q8N5H3 | 0.109786 | 0.03605 | 0.002388948 | 0.026109584 | 0.113703 | 0.019662 | 8.28e-09 | 1.25483720930233e-08 |
| 7009-6 | SL017967 | CD72 | CD72 | P21854 | 0.114051 | 0.037483 | 0.002410252 | 0.026302875 | 0.002608 | 0.017502 | 0.881573 | 0.883833845 |
| 23411-108 | SL023483 | PNMA2 | PNMA2 | Q9UL42 | -0.11427 | 0.037607 | 0.002444857 | 0.026640513 | -0.05707 | 0.017658 | 0.001245 | 0.001418657 |
| 19743-12 | SL018234 | VSTM1 | VSTM1 | Q6UX27 | 0.113441 | 0.037354 | 0.002456935 | 0.026732044 | 0.067085 | 0.019555 | 0.000612 | 0.000704145 |
| 25242-12 | SL023499 | CATIP | CATIP | Q7Z7H3 | 0.111373 | 0.036736 | 0.002498944 | 0.027148464 | 0.083414 | 0.021133 | 8.13e-05 | 9.78101538461538e-05 |
| 15653-9 | SL006511 | COAA1 | COL10A1 | Q03692 | -0.10728 | 0.035401 | 0.002509783 | 0.027225524 | -0.10002 | 0.019426 | 2.83e-07 | 3.9169203539823e-07 |
| 18398-1 | SL021346 | AK1D1 | AKR1D1 | P51857 | 0.10833 | 0.035755 | 0.002514545 | 0.027236532 | 0.209121 | 0.020456 | 4.74e-24 | 1.86265326633166e-23 |
| 25080-16 | SL023140 | UDB15 | UGT2B15 | P54855 | 0.111812 | 0.036951 | 0.00254675 | 0.027544317 | 0.03915 | 0.021073 | 0.06332 | 0.068017028 |
| 11104-13 | SL003340 | YKL-40 | CHI3L1 | P36222 | 0.107444 | 0.035517 | 0.002553343 | 0.027574584 | 0.097237 | 0.021013 | 3.9e-06 | 5.06611295681063e-06 |
| 13543-7 | SL005407 | LTB4DH | PTGR1 | Q14914 | 0.111549 | 0.036924 | 0.002587466 | 0.027901641 | 0.139788 | 0.020657 | 1.64e-11 | 2.84995555555556e-11 |
| 21815-7 | SL022111 | MOB3B | MOB3B | Q86TA1 | 0.111767 | 0.037007 | 0.002595285 | 0.027944492 | -0.01385 | 0.019006 | 0.466212 | 0.477195848 |
| 8407-84 | SL007356 | NOTC2 | NOTCH2 | Q04721 | -0.10601 | 0.03513 | 0.002617734 | 0.028144517 | -0.14246 | 0.019994 | 1.37e-12 | 2.56301435406699e-12 |
| 5675-6 | SL003198 | Tenascin | TNC | P24821 | -0.11293 | 0.037439 | 0.002627932 | 0.028212421 | -0.06507 | 0.021853 | 0.002934 | 0.003287425 |
| 4534-10 | SL012822 | BSSP4 | PRSS22 | Q9GZN4 | -0.10661 | 0.035366 | 0.002644054 | 0.028343637 | -0.12377 | 0.021226 | 6.24e-09 | 9.49354085603113e-09 |
| 19127-1 | SL007954 | HSPB6 | HSPB6 | O14558 | 0.08536 | 0.028322 | 0.002648968 | 0.028354487 | 0.108184 | 0.019674 | 4.22e-08 | 6.14532588454376e-08 |
| 9359-9 | SL008677 | EGFL9 | DLK2 | Q6UY11 | -0.09922 | 0.032934 | 0.002659002 | 0.028420036 | -0.13858 | 0.019429 | 1.29e-12 | 2.41913669064748e-12 |
| 22821-50 | SL004997 | UNG | UNG | P13051 | 0.113425 | 0.037663 | 0.002669323 | 0.028488458 | 0.113533 | 0.021881 | 2.29e-07 | 3.19212121212121e-07 |
| 17342-13 | SL012732 | AGR3 | AGR3 | Q8TD06 | 0.106741 | 0.035573 | 0.002765783 | 0.029439765 | 0.080724 | 0.020323 | 7.33e-05 | 8.85944358578053e-05 |
| 8973-23 | SL025858 | FCRL4:ECD | FCRL4 | Q96PJ5 | -0.11034 | 0.036772 | 0.00276656 | 0.029439765 | -0.07819 | 0.02184 | 0.00035 | 0.000410199 |
| 7628-40 | SL012774 | CREL1 | CRELD1 | Q96HD1 | 0.090389 | 0.030134 | 0.002775417 | 0.029490839 | 0.107342 | 0.01796 | 2.61e-09 | 4.03363636363636e-09 |
| 12848-9 | SL019951 | ARHG2 | ARHGEF2 | Q92974 | 0.112039 | 0.037452 | 0.002848499 | 0.030223197 | 0.082823 | 0.016958 | 1.11e-06 | 1.48888507718696e-06 |
| 8310-6 | SL008737 | U773 | ZG16B | Q96DA0 | -0.1123 | 0.03758 | 0.002879644 | 0.030509118 | -0.05445 | 0.017753 | 0.002186 | 0.0024633 |
| 14636-25 | SL005350 | Ribonuclease UK114 | RIDA | P52758 | 0.109573 | 0.036711 | 0.002912452 | 0.03081179 | 0.197127 | 0.020393 | 1.02e-21 | 3.3235e-21 |
| 9265-10 | SL017117 | GLIP1 | GLIPR1 | P48060 | -0.11179 | 0.037507 | 0.002952124 | 0.031186101 | -0.04292 | 0.018954 | 0.023628 | 0.025878286 |
| 8589-13 | SL017488 | CDCP1 | CDCP1 | Q9H5V8 | 0.111799 | 0.037536 | 0.00297183 | 0.031348711 | 0.032951 | 0.018036 | 0.067832 | 0.072664297 |
| 16773-29 | SL012672 | SCUB3 | SCUBE3 | Q8IX30 | -0.1109 | 0.037243 | 0.002979124 | 0.031380101 | -0.04797 | 0.019102 | 0.012096 | 0.013340986 |
| 2558-51 | SL000300 | b-Endorphin | POMC | P01189 | -0.1119 | 0.037617 | 0.003006727 | 0.031625023 | -0.04671 | 0.017343 | 0.007128 | 0.007895431 |
| 21949-4 | SL022524 | AB1IP | APBB1IP | Q7Z5R6 | 0.10972 | 0.036981 | 0.003084456 | 0.032395706 | 0.19689 | 0.020168 | 4.12e-22 | 1.4193127753304e-21 |
| 3364-76 | SL006910 | Cathepsin V | CTSV | O60911 | -0.0985 | 0.033223 | 0.003105103 | 0.032523004 | -0.14019 | 0.020481 | 9.63e-12 | 1.70376923076923e-11 |
| 25901-3 | SL024259 | CRTC3 | CRTC3 | Q6UUV7 | 0.111682 | 0.037669 | 0.003105526 | 0.032523004 | -0.00087 | 0.018329 | 0.962186 | 0.9621864 |
| 15522-2 | SL008305 | GAPR1 | GLIPR2 | Q9H4G4 | -0.11044 | 0.037362 | 0.003195614 | 0.033418302 | -0.13811 | 0.021016 | 6.06e-11 | 1.00828085106383e-10 |
| 15347-12 | SL000440 | Hemopexin | HPX | P02790 | -0.10947 | 0.037091 | 0.003243752 | 0.033872978 | -0.14958 | 0.021401 | 3.55e-12 | 6.48621495327103e-12 |
| 15589-1 | SL000431 | Gc-Globulin, Mixed Type | GC | P02774 | -0.10459 | 0.035493 | 0.003292158 | 0.034283506 | -0.1394 | 0.020478 | 1.24e-11 | 2.16930648769575e-11 |
| 16035-8 | SL003322 | VEGF sR3 | FLT4 | P35916 | -0.10875 | 0.036911 | 0.003295785 | 0.034283506 | -0.11328 | 0.021059 | 8.19e-08 | 1.17515229357798e-07 |
| 19239-5 | SL003998 | ARF1 | ARF1 | P84077 | -0.11076 | 0.0376 | 0.003301688 | 0.034283506 | -0.03972 | 0.018212 | 0.029303 | 0.031959697 |
| 13652-2 | SL020197 | TM1L1 | TOM1L1 | O75674 | -0.10716 | 0.036377 | 0.003301934 | 0.034283506 | -0.09078 | 0.018869 | 1.59e-06 | 2.11819420783646e-06 |
| 7157-22 | SL018095 | PCDA4 | PCDHA4 | Q9UN74 | -0.11079 | 0.037616 | 0.003307901 | 0.034296471 | -0.16748 | 0.020985 | 2.21e-15 | 4.84095238095238e-15 |
| 5316-54 | SL000558 | Prothrombin | F2 | P00734 | -0.10915 | 0.0371 | 0.003340362 | 0.034583689 | -0.09075 | 0.021347 | 2.2e-05 | 2.74385964912281e-05 |
| 4284-18 | SL011535 | RBM39 | RBM39 | Q14498 | 0.109803 | 0.037359 | 0.003372414 | 0.034865868 | 0.04204 | 0.018507 | 0.023198 | 0.025442968 |
| 16558-2 | SL008560 | MYOC | MYOC | Q99972 | -0.10237 | 0.034854 | 0.003395593 | 0.03505564 | -0.07897 | 0.01974 | 6.51e-05 | 7.88052631578947e-05 |
| 5481-16 | SL013754 | RASA1 | RASA1 | P20936 | 0.108776 | 0.037044 | 0.003401658 | 0.035068436 | 0.021633 | 0.019231 | 0.26073 | 0.272581468 |
| 19590-46 | SL004484 | SP-D | SFTPD | P35247 | -0.09793 | 0.033401 | 0.003452463 | 0.035541788 | -0.1439 | 0.019826 | 5.26e-13 | 1.00570171149144e-12 |
| 17816-58 | SL020927 | NCALD | NCALD | P61601 | -0.10682 | 0.036458 | 0.003471806 | 0.035690365 | -0.02769 | 0.019522 | 0.156191 | 0.165056212 |
| 24958-3 | SL023563 | RFX5 | RFX5 | P48382 | -0.10851 | 0.037053 | 0.003488719 | 0.035813573 | -0.14036 | 0.020248 | 5.29e-12 | 9.55376443418014e-12 |
| 18342-2 | SL008364 | SERC | PSAT1 | Q9Y617 | 0.102967 | 0.035172 | 0.003499533 | 0.035873914 | 0.151487 | 0.020601 | 2.62e-13 | 5.10932668329177e-13 |
| 8106-15 | SL015513 | SSRA | SSR1 | P43307 | 0.108677 | 0.037192 | 0.00356111 | 0.03645373 | 0.171376 | 0.019988 | 1.75e-17 | 4.41451612903226e-17 |
| 12830-4 | SL019934 | SAP18 | SAP18 | O00422 | 0.109353 | 0.037435 | 0.003571922 | 0.036470475 | 0.052999 | 0.019407 | 0.00636 | 0.007075042 |
| 5646-20 | SL012758 | RNAS6 | RNASE6 | Q93091 | 0.098581 | 0.033749 | 0.003572782 | 0.036470475 | 0.188919 | 0.019411 | 5.46e-22 | 1.86450655021834e-21 |
| 3152-57 | SL001800 | TNF sR-II | TNFRSF1B | P20333 | 0.097749 | 0.033472 | 0.003580681 | 0.036499848 | 0.127257 | 0.016051 | 3.35e-15 | 7.2168044077135e-15 |
| 15636-49 | SL014090 | SORC1 | SORCS1 | Q8WY21 | -0.10582 | 0.036267 | 0.003609667 | 0.036743779 | -0.09943 | 0.018364 | 6.74e-08 | 9.72450184501845e-08 |
| 3343-1 | SL005574 | Aminoacylase-1 | ACY1 | Q03154 | 0.101673 | 0.034879 | 0.003641128 | 0.037012191 | 0.149653 | 0.020121 | 1.41e-13 | 2.78439393939394e-13 |
| 13388-57 | SL012430 | NEC1 | PCSK1 | P29120 | -0.10797 | 0.037059 | 0.003658946 | 0.037141372 | -0.11656 | 0.020219 | 9.22e-09 | 1.39189961389961e-08 |
| 21395-23 | SL012360 | PCD17 | PCDH17 | O14917 | -0.1079 | 0.037041 | 0.003664948 | 0.037150411 | -0.04732 | 0.021023 | 0.024473 | 0.026766165 |
| 2597-8 | SL000002 | VEGF | VEGFA | P15692 | 0.100088 | 0.03446 | 0.003765948 | 0.038121046 | 0.114186 | 0.019422 | 4.69e-09 | 7.20546168958743e-09 |
| 5708-1 | SL007589 | LEAP2 | LEAP2 | Q969E1 | 0.108116 | 0.037239 | 0.00377905 | 0.038176965 | 0.268571 | 0.020889 | 1.18e-36 | 9.13623762376238e-36 |
| 11204-80 | SL009892 | CXAR | CXADR | P78310 | 0.107258 | 0.036946 | 0.003781978 | 0.038176965 | 0.094462 | 0.020244 | 3.23e-06 | 4.20276206322795e-06 |
| 14157-21 | SL004984 | 14-3-3E | YWHAE | P62258 | -0.10769 | 0.037119 | 0.003804367 | 0.03834971 | -0.12978 | 0.019968 | 9.77e-11 | 1.60845052631579e-10 |
| 10781-19 | SL008886 | CLC4G | CLEC4G | Q6UXB4 | 0.107569 | 0.037106 | 0.003831618 | 0.03851386 | 0.024883 | 0.018338 | 0.174947 | 0.184377789 |
| 9017-58 | SL018796 | LPH | LCT | P09848 | 0.097413 | 0.033603 | 0.003831872 | 0.03851386 | 0.020171 | 0.018376 | 0.27246 | 0.283329311 |
| 7918-114 | SL005114 | Amylase, alpha 1A | AMY1A | P04745 | -0.10207 | 0.035214 | 0.003836548 | 0.03851386 | -0.16541 | 0.019834 | 1.23e-16 | 2.9325e-16 |
| 22974-25 | SL014983 | H2B2E | H2BC21 | Q16778 | 0.103169 | 0.035614 | 0.003857698 | 0.038654967 | 0.113298 | 0.020711 | 4.95e-08 | 7.18163265306122e-08 |
| 22118-7 | SL022866 | CSAG2 | CSAG2 | Q9Y5P2 | -0.10883 | 0.037572 | 0.003861242 | 0.038654967 | 0.012083 | 0.019402 | 0.533498 | 0.544641357 |
| 4160-49 | SL000124 | MMP-2 | MMP2 | P08253 | -0.10449 | 0.036097 | 0.00388523 | 0.038788315 | -0.17671 | 0.021263 | 1.56e-16 | 3.68555891238671e-16 |
| 6064-4 | SL005292 | TXNDC4 | ERP44 | Q9BS26 | 0.107123 | 0.037008 | 0.003885236 | 0.038788315 | 0.059391 | 0.022044 | 0.007104 | 0.007879565 |
| 20428-5 | SL022166 | PHOP1 | PHOSPHO1 | Q8TCT1 | -0.10835 | 0.03745 | 0.003903022 | 0.038912432 | -0.11237 | 0.020046 | 2.31e-08 | 3.41478260869565e-08 |
| 12438-127 | SL014754 | 3MG | MPG | P29372 | 0.108276 | 0.037467 | 0.0039427 | 0.039234583 | 0.099171 | 0.017906 | 3.38e-08 | 4.9590243902439e-08 |
| 21001-393 | SL021967 | BRK1 | BRK1 | Q8WUW1 | 0.106267 | 0.036775 | 0.003946131 | 0.039234583 | 0.057322 | 0.020092 | 0.004367 | 0.004871157 |
| 8229-1 | SL018440 | GXLT1 | GXYLT1 | Q4G148 | -0.107 | 0.037035 | 0.003952286 | 0.039242096 | -0.2471 | 0.020727 | 6.88e-32 | 4.10699236641221e-31 |
| 14318-1 | SL013288 | VPS29 | VPS29 | Q9UBQ0 | 0.108739 | 0.037648 | 0.003963181 | 0.039296592 | 0.039663 | 0.021423 | 0.06423 | 0.068899885 |
| 5688-65 | SL012852 | CBLN4 | CBLN4 | Q9NTU7 | -0.10205 | 0.03536 | 0.003991438 | 0.039522849 | -0.07202 | 0.021224 | 0.000702 | 0.000804474 |
| 20913-27 | SL022028 | EIF1 | EIF1 | P41567 | 0.107634 | 0.037358 | 0.004052726 | 0.040075117 | 0.106444 | 0.02121 | 5.58e-07 | 7.61528795811518e-07 |
| 9749-190 | SL004609 | L-plastin | LCP1 | P13796 | 0.107003 | 0.037152 | 0.004065497 | 0.040146779 | 0.081141 | 0.02051 | 7.83e-05 | 9.44916666666667e-05 |
| 13990-1 | SL003652 | pyruvate carboxylase | PC | P11498 | -0.10853 | 0.037718 | 0.004101784 | 0.040450155 | -0.00363 | 0.018105 | 0.841075 | 0.845399695 |
| 17784-23 | SL013389 | GMFB | GMFB | P60983 | 0.108212 | 0.037623 | 0.004116162 | 0.040536949 | 0.076211 | 0.021357 | 0.000366 | 0.000427704 |
| 5763-67 | SL004535 | HBD-4 | DEFB104A | Q8WTQ1 | -0.07452 | 0.025938 | 0.0041595 | 0.040908315 | -0.01787 | 0.016037 | 0.265215 | 0.276530319 |
| 22973-8 | SL003175 | Gro-g | CXCL3 | P19876 | 0.10704 | 0.037277 | 0.004178088 | 0.041035602 | -0.00689 | 0.022116 | 0.755374 | 0.763181079 |
| 7933-75 | SL018350 | ADA22 | ADAM22 | Q9P0K1 | -0.10714 | 0.037331 | 0.004198331 | 0.041178774 | -0.15659 | 0.019385 | 1.03e-15 | 2.30790830945559e-15 |
| 23293-15 | SL023174 | CHAC1 | CHAC1 | Q9BUX1 | -0.10814 | 0.037706 | 0.00422341 | 0.041368924 | -0.07668 | 0.018693 | 4.23e-05 | 5.16853125e-05 |
| 12338-27 | SL019671 | EGFLA | EGFLAM | Q63HQ2 | -0.10456 | 0.036478 | 0.004245988 | 0.041534104 | -0.18985 | 0.021104 | 4.58e-19 | 1.25668771929825e-18 |
| 9713-67 | SL012503 | PGFRL | PDGFRL | Q15198 | -0.10498 | 0.036633 | 0.004251905 | 0.041536082 | -0.07566 | 0.021308 | 0.000392 | 0.00045514 |
| 21734-36 | SL022703 | C1GLT/C1GLC Complex | C1GALT1\|C1GALT1C1 | Q9NS00\|Q96EU7 | -0.1078 | 0.037654 | 0.004292766 | 0.041854643 | -0.01215 | 0.019549 | 0.534396 | 0.544847132 |
| 6947-4 | SL009530 | SIA10 | ST3GAL6 | Q9Y274 | -0.10573 | 0.036935 | 0.004297643 | 0.041854643 | -0.10026 | 0.019687 | 3.8e-07 | 5.24091710758377e-07 |
| 21643-8 | SL008141 | RS20 | RPS20 | P60866 | 0.10635 | 0.037157 | 0.004301791 | 0.041854643 | 0.133938 | 0.019431 | 6.93e-12 | 1.24010526315789e-11 |
| 16605-2 | SL017275 | C1T9A | C1QTNF9 | P0C862 | -0.10764 | 0.037716 | 0.004411653 | 0.042866167 | -0.04193 | 0.018268 | 0.021814 | 0.023992772 |
| 20087-3 | SL008697 | selenoprotein15 | SELENOF | O60613 | -0.10382 | 0.036412 | 0.004452077 | 0.043201191 | -0.17877 | 0.021363 | 9.7e-17 | 2.33396923076923e-16 |
| 19372-7 | SL021669 | MDGA2 | MDGA2 | Q7Z553 | -0.10507 | 0.036872 | 0.004473773 | 0.043353839 | -0.09955 | 0.021008 | 2.27e-06 | 2.99349072512648e-06 |
| 5680-54 | SL012845 | OBP2B | OBP2B | Q9NPH6 | -0.094 | 0.032992 | 0.004480902 | 0.043365107 | -0.1054 | 0.020806 | 4.37e-07 | 6.00586994727592e-07 |
| 9187-2 | SL013165 | HMGN1 | HMGN1 | P05114 | 0.106369 | 0.037342 | 0.00448875 | 0.043383287 | 0.115764 | 0.020911 | 3.42e-08 | 5.00831460674157e-08 |
| 16900-29 | SL012713 | MDGA1 | MDGA1 | Q8NFP4 | -0.10371 | 0.036453 | 0.004537379 | 0.043795048 | -0.1162 | 0.020883 | 2.92e-08 | 4.30026365348399e-08 |
| 18882-7 | SL021519 | CSTN2 | CLSTN2 | Q9H4D0 | -0.10571 | 0.037189 | 0.004573017 | 0.044080489 | -0.08724 | 0.02074 | 2.69e-05 | 3.34964968152866e-05 |
| 7015-8 | SL018005 | LIRB5 | LILRB5 | O75023 | -0.1043 | 0.036706 | 0.004586697 | 0.044153794 | 0.031929 | 0.021643 | 0.140273 | 0.148434923 |
| 8464-31 | SL012517 | RSPO4 | RSPO4 | Q2I0M5 | -0.08717 | 0.030684 | 0.004597741 | 0.044201559 | -0.08909 | 0.019474 | 5.01e-06 | 6.48645695364239e-06 |
| 9182-3 | SL025988 | sLRP1:ECD | LRP1 | Q07954 | -0.10533 | 0.037108 | 0.004630741 | 0.044460006 | -0.07756 | 0.018521 | 2.92e-05 | 3.63027027027027e-05 |
| 3148-49 | SL003174 | Gro-b | CXCL2 | P19875 | 0.106337 | 0.037489 | 0.00465931 | 0.04467528 | -0.01036 | 0.022087 | 0.63917 | 0.648289157 |
| 9772-153 | SL025945 | NLGN2:ECD | NLGN2 | Q8NFZ4 | -0.10668 | 0.037616 | 0.004667139 | 0.044691392 | -0.05665 | 0.020023 | 0.004702 | 0.005237283 |
| 19335-2 | SL005988 | HN1 | JPT1 | Q9UK76 | 0.105845 | 0.03738 | 0.004731561 | 0.045248668 | 0.082751 | 0.021352 | 0.000109 | 0.000130174 |
| 11955-1 | SL025968 | RHG01:Rho-GAP | ARHGAP1 | Q07960 | 0.104124 | 0.036809 | 0.00477266 | 0.045581726 | 0.101627 | 0.018788 | 6.95e-08 | 1.00090239410681e-07 |
| 20912-10 | SL022194 | HDHD1 | PUDP | Q08623 | 0.101058 | 0.035737 | 0.004787349 | 0.045662007 | 0.087371 | 0.020509 | 2.12e-05 | 2.652544e-05 |
| 21286-29 | SL014873 | RL11 | RPL11 | P62913 | 0.104473 | 0.036978 | 0.004824169 | 0.04594795 | 0.112528 | 0.018794 | 2.44e-09 | 3.77837623762376e-09 |
| 11218-84 | SL019351 | TPMT | TPMT | P51580 | 0.104273 | 0.036912 | 0.004829972 | 0.04594795 | 0.031943 | 0.019951 | 0.10948 | 0.116322075 |
| 5364-7 | SL007336 | SET | SET | Q01105 | -0.09382 | 0.033274 | 0.004908377 | 0.046632793 | -0.04159 | 0.021263 | 0.05058 | 0.054631449 |
| 14143-8 | SL014983 | H2B2E | H2BC21 | Q16778 | 0.100256 | 0.035575 | 0.004931578 | 0.04679205 | 0.138576 | 0.021257 | 8.57e-11 | 1.41986016949153e-10 |
| 22579-93 | SL009054 | NRX1B | NRXN1 | P58400 | -0.10497 | 0.037295 | 0.004986335 | 0.047082558 | -0.0211 | 0.019593 | 0.281663 | 0.292510164 |
| 4469-78 | SL007502 | ST4S6 | CHST15 | Q7LFX5 | -0.10323 | 0.036677 | 0.004987492 | 0.047082558 | -0.07712 | 0.021132 | 0.000269 | 0.000314793 |
| 17495-141 | SL018569 | SIR3 | SIRT3 | Q9NTG7 | 0.101727 | 0.036143 | 0.004987585 | 0.047082558 | 0.029576 | 0.017077 | 0.083419 | 0.088873661 |
| 5312-49 | SL000277 | Apo E2 | APOE | P02649 | -0.10492 | 0.037279 | 0.004988108 | 0.047082558 | -0.12012 | 0.021822 | 4.08e-08 | 5.95253731343284e-08 |
| 8759-29 | SL002622 | a1,4-Galactosyltransferase | A4GALT | Q9NPC4 | -0.1051 | 0.037348 | 0.004996566 | 0.047101217 | -0.14944 | 0.019131 | 8.32e-15 | 1.73963636363636e-14 |
| 9524-46 | SL019067 | S31D4 | SPATA31D4 | Q6ZUB0 | 0.10612 | 0.037729 | 0.005016536 | 0.04722822 | 0.024576 | 0.017587 | 0.162424 | 0.171410812 |
| 14158-17 | SL003848 | Annexin V | ANXA5 | P08758 | -0.10574 | 0.037612 | 0.005038856 | 0.047376983 | -0.0145 | 0.01969 | 0.461625 | 0.473119839 |
| 3795-6 | SL004642 | ADAM 9 | ADAM9 | Q13443 | -0.10574 | 0.037665 | 0.005101171 | 0.047900919 | -0.05903 | 0.018536 | 0.001469 | 0.001671795 |
| 9248-36 | SL008272 | CS010 | MYDGF | Q969H8 | 0.1053 | 0.037548 | 0.005145743 | 0.048221682 | 0.020594 | 0.018686 | 0.270522 | 0.281689104 |
| 4155-3 | SL003198 | Tenascin | TNC | P24821 | -0.10345 | 0.036893 | 0.0051486 | 0.048221682 | -0.08433 | 0.021862 | 0.000118 | 0.000139974 |
| 12528-40 | SL019761 | WRIP1 | WRNIP1 | Q96S55 | 0.10394 | 0.037079 | 0.005164094 | 0.048304545 | 0.04185 | 0.018072 | 0.020659 | 0.022753446 |
| 7195-119 | SL008537 | EPHA7 | EPHA7 | Q15375 | -0.10059 | 0.035935 | 0.005228245 | 0.048798053 | -0.03647 | 0.01843 | 0.047918 | 0.051828105 |
| 23258-56 | SL023396 | COA7 | COA7 | Q96BR5 | -0.10045 | 0.035886 | 0.005230281 | 0.048798053 | -0.02333 | 0.017681 | 0.187034 | 0.196851292 |
| 13943-38 | SL020276 | DPY30 | DPY30 | Q9C005 | 0.104409 | 0.037308 | 0.005238552 | 0.04881256 | 0.12149 | 0.019015 | 1.99e-10 | 3.24204166666667e-10 |
| 10626-116 | SL018611 | SIA7C | ST6GALNAC3 | Q8NDV1 | -0.10491 | 0.037511 | 0.005267877 | 0.049022959 | -0.08843 | 0.017029 | 2.24e-07 | 3.128e-07 |
| 9287-6 | SL008541 | SPRE | SPR | P35270 | -0.10514 | 0.037615 | 0.00529339 | 0.049197393 | -0.05927 | 0.016048 | 0.000226 | 0.00026588 |

**Supplemental Table 2.**

|  | | | | | BLSA | | | | | | | CARDIA | | | | | | |
| --- | --- | --- | --- | --- | --- | --- | --- | --- | --- | --- | --- | --- | --- | --- | --- | --- | --- | --- |
|  |  |  |  |  | Association with DSST | | | Indirect Effect | | | | Association with DSST | | | Indirect Effect | | | |
| SeqId | SomaId | Target | EntrezGeneSymbol | UniProt | **beta** | **SE** | **p** | **beta** | **SE** | **p** | % Mediation | **beta** | **SE** | **p** | **beta** | **SE** | **p** | % Mediation |
| 10565-19 | SL017951 | SLIK3 | SLITRK3 | O94933 | 0.1869 | 0.0430 | 1.50e-05 | -0.00035 | 0.00011 | 0.00128 | 20.91 | 0.08172 | 0.01722 | 2.19e-06 | -0.03747 | 0.01546 | 0.01537 | 4.67 |
| 6478-2 | SL017460 | IGLO5 | IGLON5 | A6NGN9 | 0.1547 | 0.0414 | 0.00020 | -0.00034 | 0.00011 | 0.00241 | 20.04 | 0.08900 | 0.01857 | 1.74e-06 | -0.04919 | 0.01668 | 0.00319 | 3.63 |
| 21707-15 | SL018449 | C1QL3 | C1QL3 | Q5VWW1 | 0.1871 | 0.0444 | 2.72e-05 | -0.00027 | 8.32e-05 | 0.00128 | 15.85 | 0.11772 | 0.01953 | 1.92e-09 | -0.05274 | 0.01475 | 0.00035 | 3.48 |
| 9199-6 | SL018946 | UB2G2 | UBE2G2 | P60604 | -0.1539 | 0.0448 | 0.00061 | -0.00027 | 0.00011 | 0.01523 | 15.74 | -0.08075 | 0.01796 | 7.24e-06 | -0.03702 | 0.01518 | 0.01474 | 4.72 |
| 15594-47 | SL008268 | HTRA1 | HTRA1 | Q92743 | -0.1543 | 0.0426 | 0.00031 | -0.00026 | 0.00011 | 0.02053 | 15.53 | -0.11854 | 0.01714 | 5.96e-12 | -0.06959 | 0.01983 | 0.00045 | 2.82 |
| 11214-40 | SL019363 | DNJB9 | DNAJB9 | Q9UBS3 | -0.1740 | 0.0439 | 7.89e-05 | -0.00026 | 0.00009 | 0.00387 | 15.35 | -0.06830 | 0.01708 | 6.58e-05 | -0.02818 | 0.01427 | 0.04823 | 6.29 |
| 25245-22 | SL023554 | RIC8A | RIC8A | Q9NPQ8 | -0.1993 | 0.0452 | 1.14e-05 | -0.00023 | 0.00008 | 0.00314 | 13.43 | -0.12273 | 0.01977 | 6.23e-10 | -0.03569 | 0.01190 | 0.00270 | 5.08 |
| 7210-25 | SL004470 | Amyloid-like protein 1 | APLP1 | P51693 | 0.1694 | 0.0436 | 0.00011 | -0.00023 | 0.00009 | 0.01234 | 13.41 | 0.07819 | 0.01745 | 7.81e-06 | -0.03840 | 0.01476 | 0.00928 | 4.55 |
| 6315-58 | SL017411 | PLBL1 | PLBD1 | Q6P4A8 | -0.1683 | 0.0442 | 0.00015 | -0.00022 | 0.00008 | 0.00340 | 13.11 | -0.06421 | 0.02160 | 0.00299 | -0.02084 | 0.01045 | 0.04622 | 7.90 |
| 3344-60 | SL000272 | Antithrombin III | SERPINC1 | P01008 | 0.1504 | 0.0444 | 0.00073 | -0.00021 | 0.00008 | 0.00942 | 12.57 | 0.06703 | 0.01699 | 8.18e-05 | -0.03786 | 0.01171 | 0.00122 | 4.22 |
| 8469-41 | SL000466 | IGFBP-2 | IGFBP2 | P18065 | 0.0986 | 0.0354 | 0.00546 | -0.00021 | 0.00009 | 0.02059 | 12.43 | -0.02203 | 0.01806 | 0.22248 | 0.02846 | 0.01340 | 0.03367 | -6.46 |
| 14337-1 | SL019699 | TPPC3 | TRAPPC3 | O43617 | -0.1428 | 0.0438 | 0.00115 | -0.00021 | 0.00009 | 0.02172 | 12.30 | -0.09955 | 0.02034 | 1.05e-06 | -0.03043 | 0.01361 | 0.02542 | 5.94 |
| 3175-51 | SL006610 | ATS13 | ADAMTS13 | Q76LX8 | 0.1451 | 0.0417 | 0.00053 | -0.00021 | 0.00008 | 0.00595 | 12.23 | 0.08099 | 0.01784 | 5.93e-06 | -0.02124 | 0.00782 | 0.00660 | 8.09 |
| 10754-113 | SL005237 | Prokineticin-2 | PROK2 | Q9HC23 | -0.1669 | 0.0445 | 0.00018 | -0.00020 | 0.00007 | 0.00443 | 12.09 | -0.07010 | 0.01755 | 6.7e-05 | -0.01991 | 0.00770 | 0.00975 | 8.58 |
| 5107-7 | SL005703 | Notch 1 | NOTCH1 | P46531 | 0.1446 | 0.0450 | 0.00135 | -0.00019 | 0.00008 | 0.01241 | 11.29 | 0.07282 | 0.01726 | 2.54e-05 | -0.02379 | 0.01071 | 0.02633 | 7.38 |
| 22985-160 | SL000466 | IGFBP-2 | IGFBP2 | P18065 | 0.0868 | 0.0341 | 0.01102 | -0.00019 | 0.00009 | 0.03270 | 11.25 | -0.02231 | 0.01813 | 0.21854 | 0.02876 | 0.01279 | 0.02457 | -6.42 |
| 8819-3 | SL000466 | IGFBP-2 | IGFBP2 | P18065 | 0.0895 | 0.0352 | 0.01114 | -0.00019 | 0.00009 | 0.03377 | 11.15 | -0.02449 | 0.01802 | 0.17416 | 0.02816 | 0.01262 | 0.02559 | -6.50 |
| 19768-13 | SL000395 | Cystatin B | CSTB | P04080 | -0.1174 | 0.0420 | 0.00532 | -0.00017 | 0.00008 | 0.02767 | 10.22 | -0.12070 | 0.01979 | 1.23e-09 | -0.04508 | 0.01389 | 0.00117 | 3.92 |
| 6605-17 | SL005608 | IGFALS | IGFALS | P35858 | 0.1012 | 0.0369 | 0.00616 | -0.00017 | 0.00007 | 0.02152 | 9.86 | 0.10418 | 0.01836 | 1.54e-08 | -0.02237 | 0.00817 | 0.00618 | 8.51 |
| 5939-42 | SL004365 | TWEAK | TNFSF12 | O43508 | 0.1246 | 0.0443 | 0.00496 | -0.00016 | 0.00007 | 0.02140 | 9.54 | 0.08536 | 0.02152 | 7.48e-05 | -0.02325 | 0.01068 | 0.02951 | 7.40 |
| 25960-15 | SL006111 | T-plastin | PLS3 | P13797 | -0.1639 | 0.0443 | 0.00023 | -0.00016 | 0.00006 | 0.01369 | 9.47 | -0.04776 | 0.01716 | 0.00542 | -0.01952 | 0.00839 | 0.02002 | 8.49 |
| 18380-78 | SL000254 | Albumin | ALB | P02768 | 0.0860 | 0.0415 | 0.03847 | -0.00016 | 0.00007 | 0.03516 | 9.33 | 0.08961 | 0.01792 | 6.17e-07 | -0.04459 | 0.01547 | 0.00395 | 3.79 |
| 7853-19 | SL018291 | SCO1 | SCO1 | O75880 | 0.1315 | 0.0447 | 0.00332 | -0.00016 | 0.00008 | 0.04444 | 9.17 | 0.08996 | 0.02097 | 1.86e-05 | -0.03980 | 0.01117 | 0.00036 | 3.95 |
| 16908-5 | SL010377 | OMGP | OMG | P23515 | 0.1084 | 0.0440 | 0.01397 | -0.00015 | 0.00008 | 0.04646 | 8.92 | 0.10031 | 0.01749 | 1.1e-08 | -0.04642 | 0.01410 | 0.00099 | 3.83 |
| 19555-1 | SL013060 | SUMO2 | SUMO2 | P61956 | -0.1233 | 0.0430 | 0.00423 | -0.00014 | 0.00007 | 0.02924 | 8.55 | -0.08869 | 0.01852 | 1.78e-06 | -0.02040 | 0.01012 | 0.04391 | 8.88 |
| 16288-17 | SL013797 | EPHA4 | EPHA4 | P54764 | 0.1134 | 0.0449 | 0.01181 | -0.00012 | 0.00006 | 0.04838 | 7.24 | 0.07110 | 0.01721 | 3.74e-05 | -0.02307 | 0.00929 | 0.01298 | 7.50 |
